# Landscaping of Urine Proteome: Unlocking Diagnostic Potential and Overcoming Unique Challenges

**DOI:** 10.1101/2023.10.27.23297705

**Authors:** Bogdan Budnik, Hossein Amirkhani, Klaus Weinberger, Karine Sargsyan, Mohammad H. Forouzanfar, Ashkan Afshin

## Abstract

This study explores the application of deep proteomic profiling to extract disease-specific features from urine. Early detection of cancer and other chronic disorders is crucial for better outcomes, but traditional diagnostics as well as emerging genomic-based diagnostics are expensive and invasive. Our research reveals that a select group of urinary proteins can accurately detect early-stage diseases with high sensitivity, surpassing current tests. While urine-based protein panels could offer cost-effective and accurate alternatives to current screening methods, kidney factors and blood urine barrier pathologies could pose significant challenges. New diagnostic technologies may emerge because of these findings, ushering in an era of early detection for cancer and chronic diseases.

**One-Sentence Summary:** Urine-based protein panels show distinct patterns in early disease detection, promising opportunities for advancing diagnostic tests

## Main Text

Urine-based diagnostics are becoming increasingly popular as a non-invasive and cost-effective method of early detection of various types of diseases.(*1-8*) In recent years, urine-based cancer diagnostics have been developed for a variety of different types of cancer, including bladder, prostate, and colorectal cancer. For example, a urine-based test called UroSEEK has been developed for the early detection of bladder cancer. The test detects mutations in 11 cancer-associated genes in urine samples and has shown a sensitivity of 96% and specificity of 88% in early detection of urothelial cancer.(*9*) EpiCheck is another example of urine-based test detecting 15 DNA methylation targets in urine samples and has demonstrated a sensitivity of 68% at the specificity of 88% for bladder cancer.(*9*) Additionally, a 19-protein urinary biomarker model was recently developed and exhibited an 87% sensitivity and 65% specificity, outperforming traditional markers.(*10*)

While urine is seen as an extract and proxy of the serum, reliable measurement of proteins in urine to develop effective and widely available diagnostic tests is challenging.(*11*) The low abundance of biomarkers in urine, the physiology of kidney and urinary system, and the high variability in sample collection and processing can make it difficult to obtain accurate and consistent results and interpret the variation across different diseases and healthy populations. Additionally, identifying biomarkers that are specific and sensitive enough to detect diseases in low levels has been a major challenge for developing effective urine-based screening tests.(*12*)

Recent advancements in protein measurements, particularly with innovations like the Proximity Extension Assay, coupled with advanced AI algorithms, hold the transformative potential to reshape the landscape of urine-based diagnostics. These innovations not only enable the detection of low-abundance biomarkers in urine with remarkable sensitivity and specificity (*13-19*) but also open the door to a more comprehensive diagnostic approach. Rather than concentrating solely on individual biomarkers, we are now poised to identify the unique patterns associated with each disease within urine samples. This paradigm shift in diagnostics offers the promise of developing more robust and promising biomarker panels for the early detection of various diseases.

Additionally, the establishment of standardized protocols for urine collection, processing, and storage represents a critical step in our pursuit of reliable and accurate urine-based diagnostic tests. These protocols not only reduce variability but also significantly enhance the overall precision and reliability of the diagnostic process.

In this study, leveraging these advancements, we have embarked on landscaping of the urine proteome to develop novel diagnostics for a range of diseases.

### Urine proteome landscape in healthy individuals

We utilized the PEA technology to detect a total of 3,072 proteins in urine samples, and successfully identified 2,850 of these proteins. This subset of 2,850 proteins formed the basis for the exploration of novel biomarkers in urine. Notably, all proteins were directly assessed from urine samples with a fourfold dilution, primarily aimed at minimizing the presence of salt in the samples. It is noteworthy that no further dilution was necessary for the PEA technology analysis, suggesting that the concentrations of the detected proteins generally remained at lower levels across the entire spectrum of proteins examined.

Our analysis indicated that the concentrations of certain proteins did not display significant differences in relation to the age of the individuals. As illustrated in Figure 1.A, specific proteins, such as VEGFB and IL11, exhibited relatively higher urinary concentrations in older patients, while proteins like BOLA1, ARHGEF, and NFKB2 showed lower urinary concentrations in older individuals. It is pertinent to mention that only a limited number of proteins demonstrated a p-value of <0.01, and only one protein exhibited a p-value of 0.0016, indicating the overall subtle age-related differences in protein concentration in urine.

**Figure 1.**
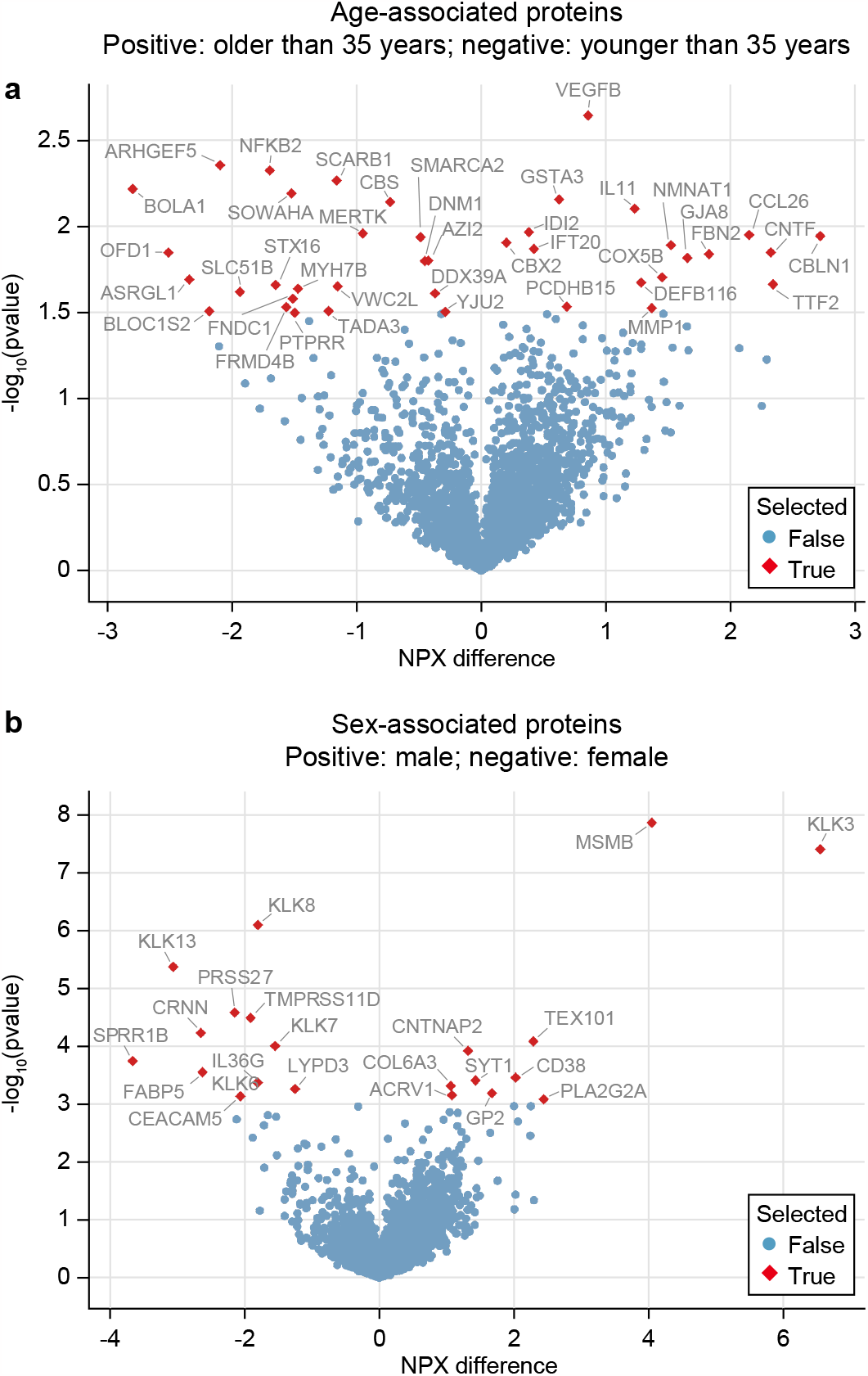
Variations in Urine Proteome by Age and Sex. Volcano plots depicting differential protein abundances in healthy adults aged above and below 35 years (a), and between adult males and females (b). Red points indicate proteins with the lowest p-values for statistical differences.

Furthermore, protein concentrations in urine exhibited variations based on the biological sex of the individuals. Figure 1.B depicts these differences in protein abundances between healthy males and females. Notably, Kallikreins, in particular, proved to be highly sensitive to the sex of the samples. Among these proteins, KLK3 and MSMB displayed significantly higher concentrations in male urine samples, while proteins KLK8 and KLK13 exhibited significantly higher concentrations in female samples. In total, our analysis identified more than 20 proteins with a significant p-value of <0.001, indicating a more pronounced contrast among patients when considering biological sex, in comparison to age.

### Urine proteome landscape across diseases

The volcano plots, as illustrated in Figure 2, provide an insightful perspective on the differential expression of proteins in various medical conditions under examination in this study. Notably, it is apparent that most of these conditions exhibited an asymmetric pattern, characterized by overrepresentation of proteins in patients compared to normal samples, except for the case of melanoma. The observations collectively reveal three distinct patterns. Firstly, in the context of melanoma and to lesser extent endometrial cancer, the volcano plot exhibits a symmetrical pattern without a statistically significant changes in the great majority of the proteins. Secondly, in cancers of the cervix, ovary, and prostate as well as in MS, an asymmetrical shape is observed while majority of the protein changes were non-significant. Thirdly, the third pattern was related to cancers of kidney, bladder, and NASH where we observed an asymmetrical shape on the volcano plots and significant increase in the great majority of proteins. These findings underscore the substantial role of the kidneys in protein excretion in urine, which holds important implications for urine proteomics studies. Overall, our results consistently point to a distinctive pattern with statistically significant over-presentation of proteins in patients, offering potential utility for disease detection across a broad spectrum of conditions.

**Figure 2.**
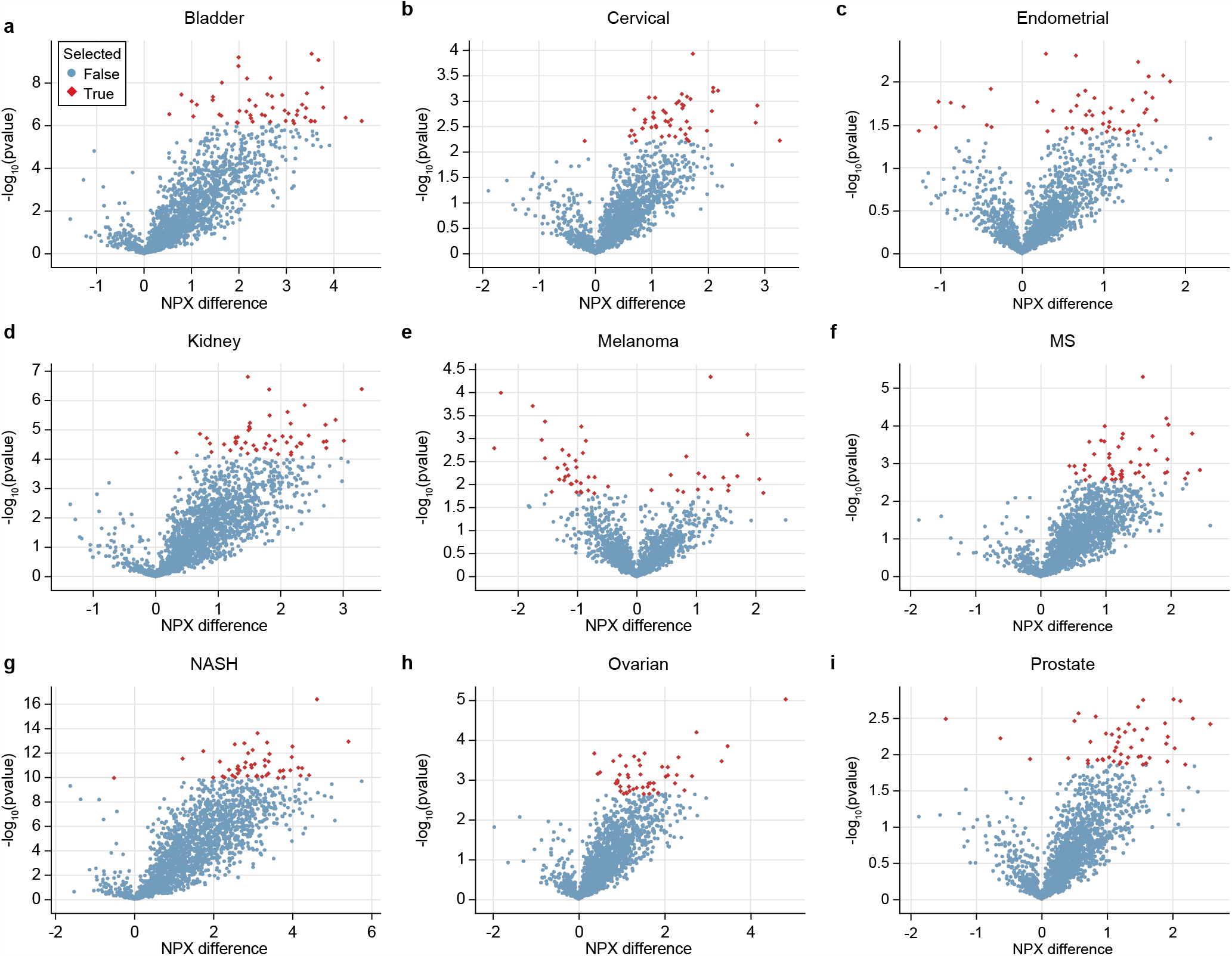
Variations in Urine Proteome across Different Diseases. Volcano plots showing differential protein abundances in healthy adults and patients with various conditions: bladder cancer (a), cervical cancer (b), endometrial cancer (c), kidney cancer (d), melanoma (e), multiple sclerosis (MS) (f), nonalcoholic steatohepatitis (NASH) (g), ovarian cancer (h), and prostate cancer (i). Red points represent proteins with the lowest p-values for statistical differences.

The analysis of correlations between different proteins has revealed intriguing patterns (as shown in Figure S1). To simplify the interpretation of these correlation matrices while ensuring their clarity, we chose to focus on the top 100 proteins with the lowest p-values in disease-normal comparisons. This selective approach allowed us to assess changes in the proteomic landscape across different diseases. For example, when examining the top 100 proteins that distinguish bladder, kidney, and NASH, we observe notably high correlations in normal samples. This suggests a potential non-differential over-leakage of these proteins into urine. In contrast, the correlation between proteins that vary between melanoma and endometrial cancer patients and their respective normal samples is considerably lower, indicating distinct urinary presentations. Furthermore, when comparing the protein correlations in healthy controls and patients, we observe significant differences in correlation patterns for the top 100 proteins in the cases of cervical cancer and prostate cancer. In summary, the volcano plots and correlation matrices together emphasize the unique urinary patterns associated with various diseases, underscoring their pivotal role in developing diagnostics for each disease category.

Figure S2 illustrates the distribution of over-represented and under-represented proteins across various diseases, similar to Figure S1, focusing only on the top 100 proteins with the lowest p-values in disease-normal comparisons. The data underscores the specificity of proteins associated with each disease. As corroborated by the volcano plots, the majority of proteins were over-represented. Notably, among the top 100 proteins with the lowest p-values, only melanoma, prostate cancer, endometrial cancer, and NASH exhibited under-represented proteins in urine. In diagram S2, the over-represented proteins were uniquely linked to individual diseases, with no proteins common to more than two conditions, suggesting a disease-specific protein signature. Figure S2B displays the under-represented proteins identified in the study, found in fewer diseases. These cases are highly disease-specific, emphasizing that urine proteomes exhibit distinctive protein changes in response to diseases, thereby underscoring the specificity and diagnostic potential of urine proteomics in the context of various diseases.

### Optimal disease-specific protein signature

In Figure 3, we present the AUC scores achieved for each of the diseases under examination, with consideration to the number of proteins utilized. Generally, we observed that an optimal disease detection performance, characterized by AUC scores exceeding 0.9, required the incorporation of at least 7 proteins. Notably, ovarian cancer was the sole exception, where the maximum AUC score was attained with a minimum set of 15 proteins. On the other hand, our analysis reveals a distinctive case in our study—namely, the detection of MS, where the highest AUC score achieved was 0.88, despite the utilization of 17 proteins. This unique instance suggests that this neurological disease might exhibit minimal alterations in its protein signature within urine, offering a plausible explanation for the relatively lower AUC score.

**Figure 3.**
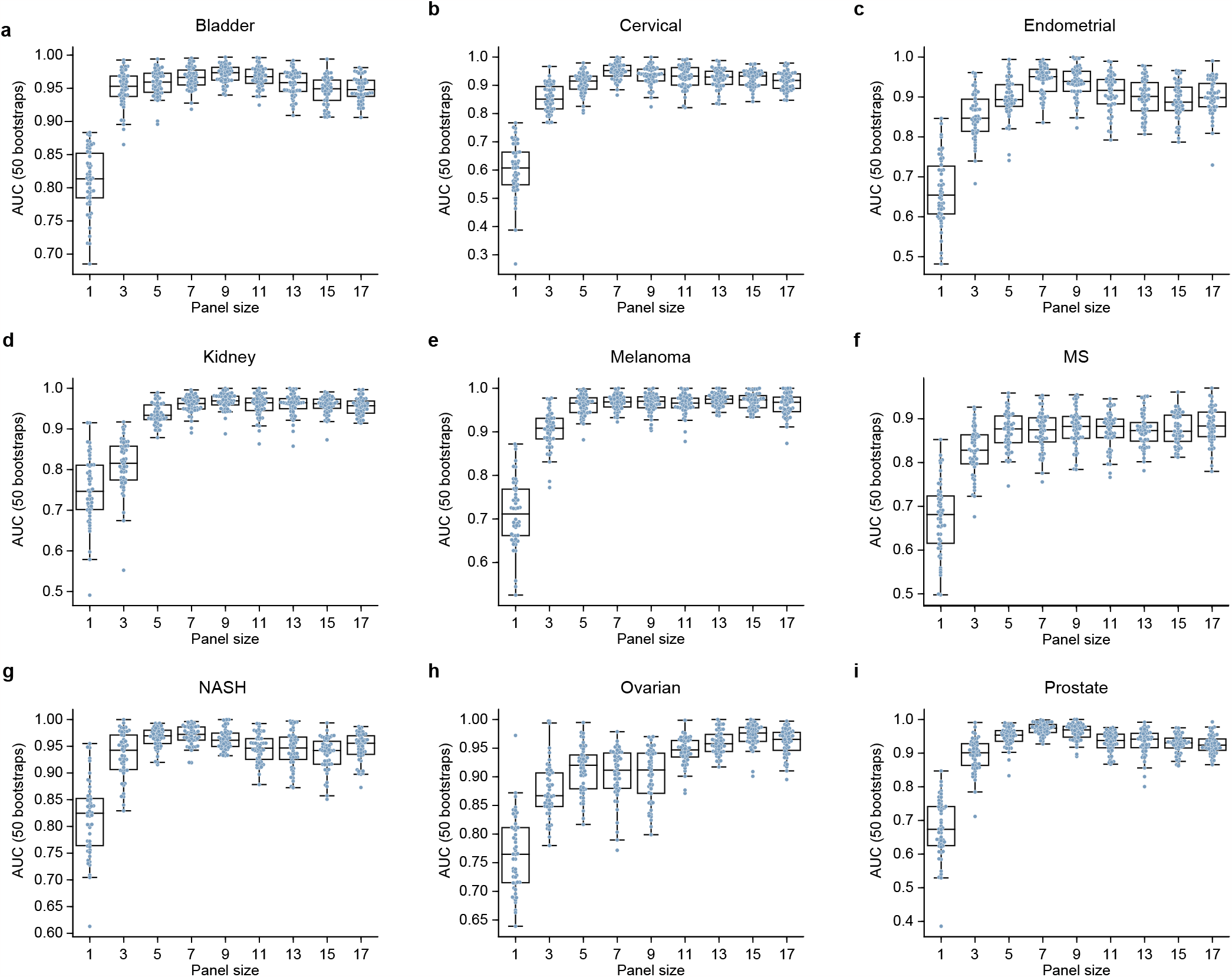
The relationship between the size of the protein panel and the performance of panel. The X-axis displays the number of proteins in the panel, while the Y-axis measures panel performance using the AUC for various conditions: bladder cancer (a), cervical cancer (b), endometrial cancer (c), kidney cancer (d), melanoma (e), multiple sclerosis (f), nonalcoholic steatohepatitis (NASH) (g), ovarian cancer (h), and prostate cancer (i).

Figure 4 illustrates the performance of both individual proteins within the panel and the overall performance of the panel. As depicted in the figure, the overall performance of the panel surpassed that of any individual protein in most instances. This underscores the distinct contribution of each protein within the panel in characterizing the unique proteomic signature of the disease.

**Figure 4.**
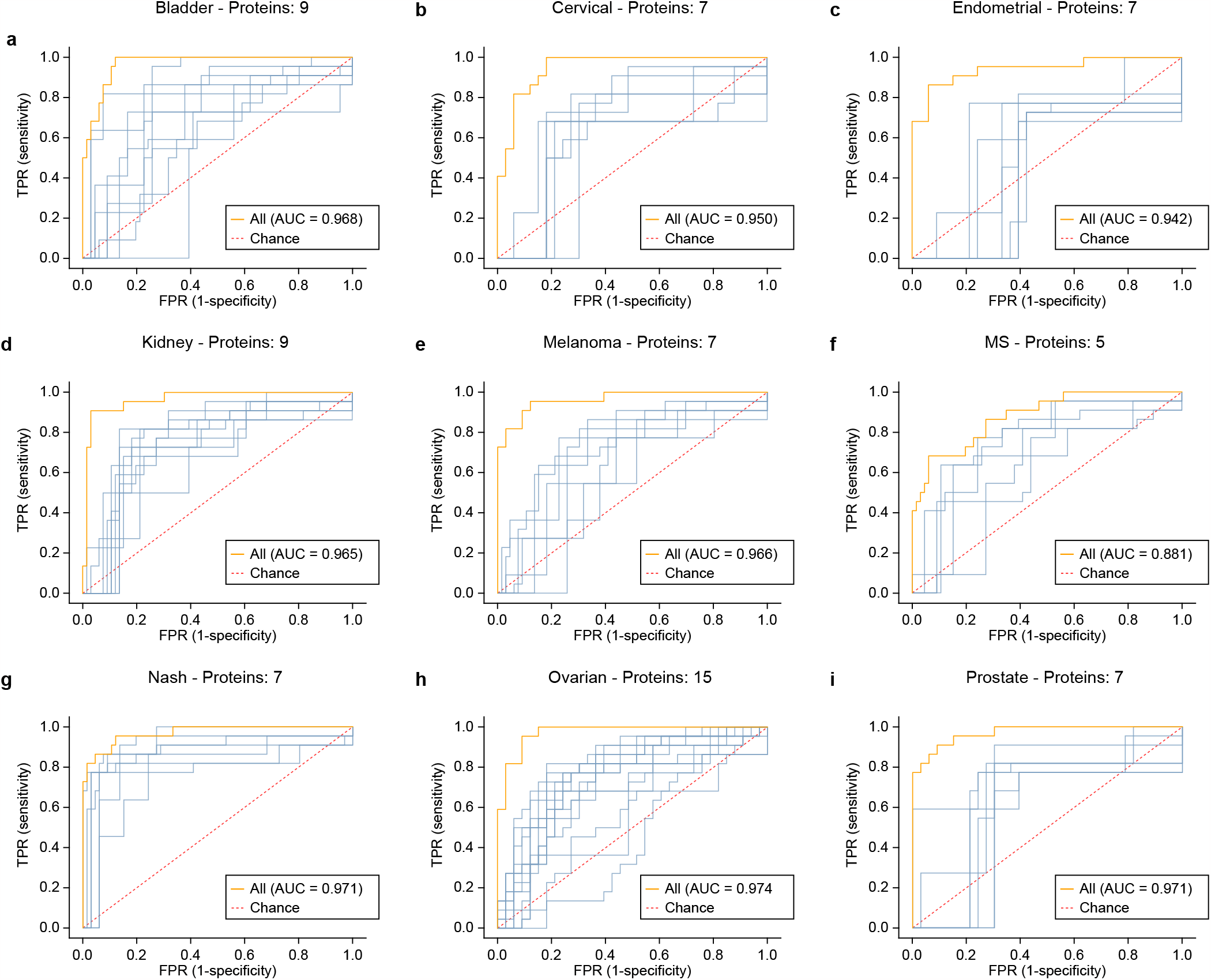
The performance of the selected protein panel. ROC curves depict the performance of selected protein panels for each condition: bladder cancer (a), cervical cancer (b), endometrial cancer (c), kidney cancer (d), melanoma (e), multiple sclerosis (f), nonalcoholic steatohepatitis (NASH) (g), ovarian cancer (h), and prostate cancer (i). Blue lines represent individual protein performance, while the orange line represents the overall panel performance.

While the urinary panels exhibited high performance (AUC>0.95) for most diseases in the study, there was a noticeable trade-off between sensitivity and specificity across diseases. For instance, in the cases of prostate cancer, NASH, and melanoma, relatively high sensitivity could be achieved at 99% specificity. However, achieving high sensitivity for panels related to cervical and endometrial cancers proved more challenging and necessitated a significant reduction in panel specificity.

Figure 5 illustrates the quantified significance of individual proteins for various diseases. To determine the importance of each protein, we employed a random forest classifier trained on each disease using a feature set composed of concatenated proteins from all panels. The feature importance scores generated reflect the normalized total reduction in Gini impurities resulting from the utilization of a specific protein as a feature. Gini impurity serves as a metric to gauge how frequently a randomly selected element from the dataset would be incorrectly classified if it were randomly labeled according to the distribution of labels within the subset. The Gini impurity reaches its minimum value when all cases within the node exclusively belong to a single class. In this context, the scores indicate the influence of each protein in reducing the mixture or impurity of the samples.

**Figure 5.**
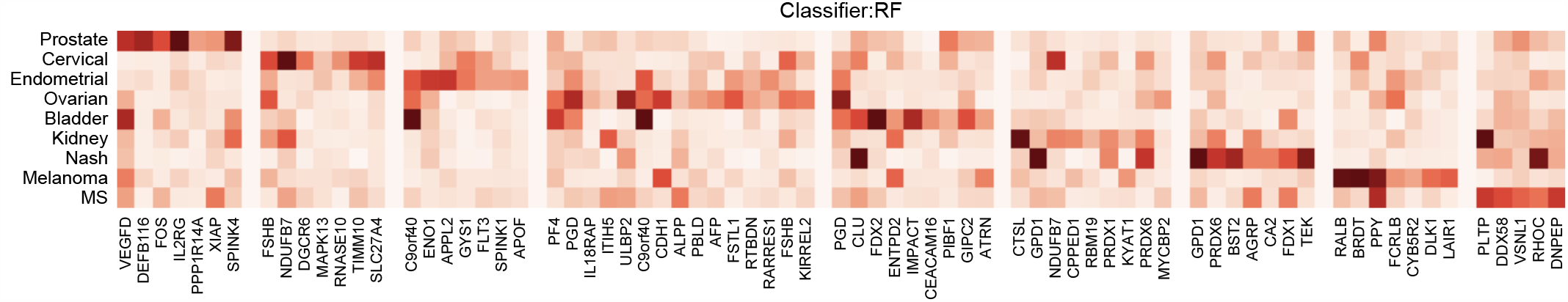
Distinctive Significance of Protein Sets for Disease Detection. This heatmap illustrates the distinct importance of each protein of the panel in detecting other diseases.

The heatmap in Figure 5 effectively portrays the unique significance of protein sets for detecting specific diseases. The majority of disease detection sets typically comprise seven proteins, with the exception of the ovarian cancer protein set, which comprises 15 proteins to achieve an acceptable level of specificity. It is noteworthy that proteins exhibiting the highest predictive specificity for their target diseases are the most prevalent. Moreover, certain proteins play a crucial role in the detection of multiple diseases, in addition to their primary target. For instance, the protein VEGFD, included in the prostate cancer protein set, also proves to be highly significant for bladder cancer. In the case of bladder cancer, protein C9orf40 exhibited substantial importance, even though it wasn’t selected for the final set. Another example is protein PPY, which demonstrated equivalent importance for both melanoma and multiple sclerosis detection.

### Interpretation

Our landscaping of the urine proteome revealed the potential of utilizing low-abundance proteins in urine for the early detection of cancer, metabolic disorders, and neurological conditions. This finding forms the basis for the development of a range of non-invasive urine-based screening tests capable of identifying a variety of diseases. The early diagnosis of conditions such as cancer and metabolic disorders is essential for the development of effective treatments. Our study has demonstrated that distinct biological signals can be detected in urine even during the initial stages of diseases.

Furthermore, our findings indicate that proteins characterizing the urinary pattern of each disease is relatively unique and are not affected by the other conditions. This offers a promising avenue for the development of non-invasive urine-based tests designed to detect disease-specific proteins. These tests, if implemented, could enable early disease diagnosis, thereby preventing disease progression and facilitating the development of more effective treatments. In essence, our research has the potential to significantly impact healthcare by improving early disease detection and advancing public health.

Alterations in the urine proteome can be attributed to structural or physiological damage within the kidneys, potentially affecting the integrity of the blood-urine barrier. It is worth noting that distinguishing certain diseases that share similarities in protein classes with kidney damage can be challenging. Therefore, in the development of protein-based tests designed to detect a range of disease types, careful consideration of these complexities is imperative. Recent insights gleaned from the CKD273 biomarker panel, specifically tailored for the identification of impaired kidney function, have shed light on the predominant biomarkers, primarily comprising collagen fragments originating from modified extracellular matrix turnover.(*9*) This knowledge offers a practical opportunity to either accommodate or differentiate kidney-related changes when formulating diagnostic tests for other diseases.

Our new generation of protein-based urine test has exhibited remarkable sensitivity in the early detection of a variety of tumors in asymptomatic individuals, positioning it as a strong candidate for widespread screening—a role currently unattainable with existing methods. The non-invasive and cost-effective nature of urine testing makes it a practical option for screening large populations for cancer, especially among individuals with risk-elevating lifestyle factors or family histories. The potential for earlier detection and subsequent treatment holds promise for substantially improving patient outcomes.

In addition to our comprehensive protein measurement coverage and the accuracy of these measurements, even for proteins present in small quantities; our strengths encompass developing a machine learning platform for extracting unique features from a wide range of urinary proteins. Furthermore, our approach encompasses diseases with significant unmet diagnostic needs. There are also limitations to consider, including the small size of the cohort and the presence of comorbidities. Hence, we need to validate our protein panels in a larger population cohort before it can be widely accepted across a variety of populations. Additionally, the test should be evaluated for accuracy and precision in different populations. Finally, the cost and ease-of-use of the test should be determined.

In summary, this study presents several distinct contributions. These include an analysis of the most extensive proteomics dataset derived from urine, the formulation of a cancer-specific protein signature tailored for early-stage cancers, with an emphasis on the baseline carcinogenic state rather than the later-stage tumor behavior and human response.

## Data Availability

All data produced in the present study are available upon reasonable request to the authors.

## Author contributions

*All authors contributed to drafting the overall structure and flow of the manuscript. Subsequently, authors contributed subsections based on their domain of expertise. AA integrated subsections, incorporated comments and performed additional revisions*.

## Data and materials availability

*The datasets generated during and/or analyzed during the current study are available from the corresponding author on reasonable request*.

## References

1. E. Rodriguez-Suarez, J. Siwy, P. Zurbig, H. Mischak, Urine as a source for clinical proteome analysis: from discovery to clinical application. Biochim Biophys Acta 1844, 884–898 (2014).

2. A. Kentsis, Challenges and opportunities for discovery of disease biomarkers using urine proteomics. Pediatr Int 53, 1–6 (2011).

3. C. J. Rosser et al., Urinary protein biomarker panel for the detection of recurrent bladder cancer. Cancer Epidemiol Biomarkers Prev 23, 1340–1345 (2014).

4. N. Davis et al., A Novel Urine-Based Assay for Bladder Cancer Diagnosis: Multi-Institutional Validation Study. Eur Urol Focus 4, 388–394 (2018).

5. M. Frantzi et al., Development and Validation of Urine-based Peptide Biomarker Panels for Detecting Bladder Cancer in a Multi-center Study. Clin Cancer Res 22, 4077–4086 (2016).

6. S. Thomas, L. Hao, W. A. Ricke, L. Li, Biomarker discovery in mass spectrometry-based urinary proteomics. Proteomics Clin Appl 10, 358–370 (2016).

7. P. Kumar et al., Highly sensitive and specific novel biomarkers for the diagnosis of transitional bladder carcinoma. Oncotarget 6, 13539–13549 (2015).

8. W. S. Tan et al., Novel urinary biomarkers for the detection of bladder cancer: A systematic review. Cancer Treat Rev 69, 39–52 (2018).

9. A. Argiles et al., CKD273, a new proteomics classifier assessing CKD and its prognosis. PLoS One 8, e62837 (2013).

10. M. Frantzi et al., Validation of diagnostic nomograms based on CE-MS urinary biomarkers to detect clinically significant prostate cancer. World J Urol 40, 2195–2203 (2022).

11. N. Chebotareva et al., Urinary Protein and Peptide Markers in Chronic Kidney Disease. Int J Mol Sci 22, (2021).

12. Q. U. Ain, S. Muhammad, Y. Hai, L. Peiling, The role of urine and serum biomarkers in the early detection of ovarian epithelial tumours. J Obstet Gynaecol 42, 3441–3449 (2022).

13. J. Daza et al., Urine supernatant reveals a signature that predicts survival in clear-cell renal cell carcinoma. BJU Int 132, 75–83 (2023).

14. J. Suh et al., Next-generation Proteomics-Based Discovery, Verification, and Validation of Urine Biomarkers for Bladder Cancer Diagnosis. Cancer Res Treat 54, 882–893 (2022).

15. T. Sun et al., Diagnostic value of a comprehensive, urothelial carcinoma-specific next-generation sequencing panel in urine cytology and bladder tumor specimens. Cancer Cytopathol 129, 537–547 (2021).

16. L. M. Chen et al., External validation of a multiplex urinary protein panel for the detection of bladder cancer in a multicenter cohort. Cancer Epidemiol Biomarkers Prev 23, 1804–1812 (2014).

17. E. Schiffer et al., Prediction of muscle-invasive bladder cancer using urinary proteomics. Clin Cancer Res 15, 4935–4943 (2009).

18. C. K. Chen, J. Liao, M. S. Li, B. L. Khoo, Urine biopsy technologies: Cancer and beyond. Theranostics 10, 7872–7888 (2020).

19. M. Maas, T. Todenhofer, P. C. Black, Urine biomarkers in bladder cancer - current status and future perspectives. Nat Rev Urol 20, 597–614 (2023).

