## Supplementary Materials for "Landscaping of Urine Proteome: Unlocking Diagnostic Potential and Overcoming Unique Challenges"

### Materials and Methods

#### Study Design

We collected urine samples from 266 healthy individuals and patients diagnosed with various disease types including solid tumors (such as cancers of kidney, bladder, prostate, ovary, cervix, uterine, and melanoma), a neurologic disorder (i.e., multiple sclerosis), and a metabolic disorder (i.e., nonalcoholic steatohepatitis). Our focus was on the early stages of these diseases, where timely detection followed by medical or surgical intervention can significantly improve patient survival rates. Utilizing Proximity Extension Assay technology, we conducted deep proteomic profiling on the urine samples, identifying over 2,850 proteins in each sample. Through a two-step model, significant proteins were isolated and analyzed using Light Gradient Boosting Model (LGBM) to ascertain a robust set of proteins with the highest predictive performance.

#### Samples

The urine samples utilized in this analysis were supplied by the Ukraine Association of Biobank (UAB). These samples were primarily collected from patients undergoing routine medical check-ups, who were identified to have early-stage diseases. All the patients incorporated in this study were treatment-naïve, and for those with cancer, their urine samples were collected prior to tumor removal or any other form of treatment. The control samples were obtained from healthy donors.

#### Protein Measurements

The protein levels in urine were measured using Olink® Explore 3072 technology, as previously described in detail. A comprehensive list of the measured proteins, including their characteristics, is available in Table S1. All protein measurements for the samples were conducted in a single run. In summary, this technology relies on antibody-based detection to quantify the levels of 3,072 target proteins in plasma. Antibodies, each conjugated with two complementary probes, were organized into eight distinct 384-plex panels. Within each panel, three control assays (interleukin-6 (IL6), interleukin-8 (CXCL8), and tumor necrosis factor (TNF)) were included for quality control purposes.
The assay process commenced with an overnight incubation to facilitate the binding of conjugated antibodies to the target proteins in the samples. This was followed by an extension and pre-amplification step, enabling the hybridization and extension of complementary probes. The extended DNA was subsequently amplified via PCR and indexed to prepare libraries, which were then subjected to sequencing on the Illumina NovaSeq platform. The resulting sequencing counts underwent a rigorous quality control and normalization procedure, incorporating internal controls to minimize intra-assay variability. These internal controls encompassed an incubation control with a non-human antigen, an extension control with a unique probe pair, and an amplification control with a double-stranded DNA sequence. Additionally, external controls, including a negative control (buffer sample) and plate controls (a plasma pool), were employed to establish the limit of detection and standardize measurements between plates. Finally, two known samples were used as controls to assess the precision of the measurements. Following quality control and normalization, the data was presented in Normalized Protein eXpression (NPX) units, logarithmically transformed (log2 scale), with higher NPX values indicating higher protein levels.

Olink's panel performance has been extensively validated for various analytical parameters, including sensitivity, dynamic range, specificity, precision, and scalability. The analytical measuring range was determined by establishing the lower limit of quantification (LLOQ) and upper limit of quantification (ULOQ) for each analyte, reported in pg/mL. Additionally, the potential occurrence of a high-dose hook effect, characterized by antigen excess in comparison to the reagent antibodies, leading to erroneously lower values, has been identified for all analytes. All assays have undergone rigorous validation for precision (encompassing repeatability and reproducibility). Intra-assay variation (within a single run) was calculated by determining the mean coefficient of variation (CV) across 6 individual samples in each of 7 separate runs conducted during the validation studies. Likewise, inter-assay variation (between different runs) was computed as the mean CV across the same 6 individual samples, measured across 7 separate runs during the validation studies.

**Statistical Approach**

As with other biomarker studies, our experiment size was relatively small. Therefore, we adopted a two-step approach to identify a minimal set of proteins that could effectively classify samples within a stable model, ensuring general applicability. In the first phase, we identified proteins that exhibited statistically significant and robust differences between the diseased and healthy samples. Subsequently, to determine the specific features comprising this minimal set, we utilized the LGBM on 100 bootstrap samples generated from the original dataset. We then assessed the model's performance using these selected features through a leave-one-out validation process. Our performance evaluation relied on the commonly accepted statistical measure known as the Area Under the Curve (AUC) of the Receiver Operating Characteristic (ROC) curve, which offered a reliable assessment of the biomarker panels' effectiveness.

The pre-processing and modelling were carried out using the Scikit-learn Python library.(*1*) After identifying the biomarkers, LGBM classifiers were trained on the training data utilizing the selected biomarkers. The predicted probability for each sample is compared to a threshold to determine whether it is normal or cancerous. The threshold is set to attain the desired level of specificity.

For data visualization, we used matplotlib (version 3.6.0),(*2*) seaborn (version 0.12.0),(*3*) and plotly (version 5.10.0).(*4*) Pre-processing and modelling were performed in Python (version 3.9.13), using scikit-learn (version 1.1.2),(*1*) pandas (version 1.4.4),(*5*) numpy (version 1.23.3),(*6*) and statsmodels (version 0.13.2).(*7*)

### References


1. F. Pedregosa, G. Varoquaux, A. Gramfort, V. Michel, B. Thirion, Scikit-learn: Machine Learning in Python. *Journal of Machine Learning Research* **12**, 2825-2830 (2011).
2. J. D. Hunter, Matplotlib: A 2D Graphics Environment. *Computing in Science and Engineering* **9**, 90–95 (2007).
3. M. L. Waskom, Seaborn: statistical data visualization. *Journal of Open Source Software* **6**, (2021).
4. Plotly Technologies Inc. (Plotly Technologies Inc., 2015).
5. W. McKinney, in *9th Python in Science Conference*. (2010), vol. 445.
6. C. R. Harris *et al.*, Array programming with NumPy. *Nature* **585**, 357-362 (2020).
7. S. Seabold, J. Perktold, in *9th Python in Science Conference*. (2010), vol. 57.

**Table S1.** List of proteins included in the Olink® Explore 3072 analysis and their characteristics.

| UniProt | Panel | LOD (pg/ml) | LLOQ (pg/ml) | ULOQ (pg/ml) | Hook (pg/mL) | Range (log10) | Intra-CV (%) | Inter-CV (%) |
|---|---|---|---|---|---|---|---|---|
| P31483 | Cardiometabolic | | | | | | 9 | 27 |
| P21964 | Cardiometabolic | 97.7 | 195.3 | 100000 | 800000 | 2.7 | 8 | 10 |
| Q9NRD8 | Cardiometabolic | 97.7 | 195.3 | 100000 | 200000 | 2.7 | 7 | 31 |
| P16860 | Cardiometabolic | | | | | | 10 | 18 |
| O60635 | Cardiometabolic | | | | | | 7 | 9 |
| O96017 | Cardiometabolic | | | | | | 13 | 19 |
| Q9UKL0 | Cardiometabolic | 1.5 | 3.1 | 3125 | 12500 | 3.0 | 7 | 12 |
| Q8NHS0 | Cardiometabolic | | | | | | 9 | 10 |
| P58546 | Cardiometabolic | | | | | | 8 | 8 |
| O43854 | Cardiometabolic | 24.4 | 48.8 | 200000 | 800000 | 3.6 | 8 | 13 |
| P40225 | Cardiometabolic | 97.7 | 781.3 | 200000 | 400000 | 2.4 | 9 | 14 |
| Q99549 | Cardiometabolic | 48.8 | 97.7 | 12500 | 25000 | 2.1 | 8 | 19 |
| P08319 | Cardiometabolic | | | | | | 7 | 8 |
| P25815 | Cardiometabolic | | | | | | 12 | 15 |
| Q8TE57 | Cardiometabolic | 6.1 | 24.4 | 25000 | 200000 | 3.0 | 9 | 6 |
| Q04760 | Cardiometabolic | | | | | | 7 | 9 |
| Q9BYF1 | Cardiometabolic | 48.8 | 97.7 | 100000 | 400000 | 3.0 | 9 | 10 |
| O14793 | Cardiometabolic | 390.6 | 781.3 | 50000 | 400000 | 1.8 | 10 | 14 |
| Q9NWQ8 | Cardiometabolic | 24.4 | 48.8 | 6250 | 200000 | 2.1 | 11 | 11 |
| Q13444 | Cardiometabolic | 24.4 | 48.8 | 25000 | 200000 | 2.7 | 7 | 10 |
| P34913 | Cardiometabolic | 390.6 | 781.3 | 100000 | 200000 | 2.1 | 7 | 21 |
| P09496 | Cardiometabolic | 3125.0 | 6250.0 | 6400000 | 12800000 | 3.0 | 7 | 18 |
| P34947 | Cardiometabolic | 1562.5 | 3125.0 | 400000 | 800000 | 2.1 | 8 | 14 |
| P55259 | Cardiometabolic | 3.1 | 6.1 | 12500 | 200000 | 3.3 | 6 | 9 |
| P01375 | Cardiometabolic | 6.1 | 12.2 | 6250 | 200000 | 2.7 | 8 | 16 |
| P52789 | Cardiometabolic | 97.7 | 195.3 | 50000 | 200000 | 2.4 | 9 | 39 |
| P09668 | Cardiometabolic | | | | | | 11 | 10 |
| O75354 | Cardiometabolic | 195.3 | 781.3 | 50000 | 800000 | 1.8 | 8 | 8 |
| Q9BWV1 | Cardiometabolic | | | | | | 8 | 10 |

Figure S1. Correlation Matrices of the Top 100 Differentially Represented Proteins between Patients and Healthy Controls. Matrices are displayed separately for healthy controls (A) and patients (B).

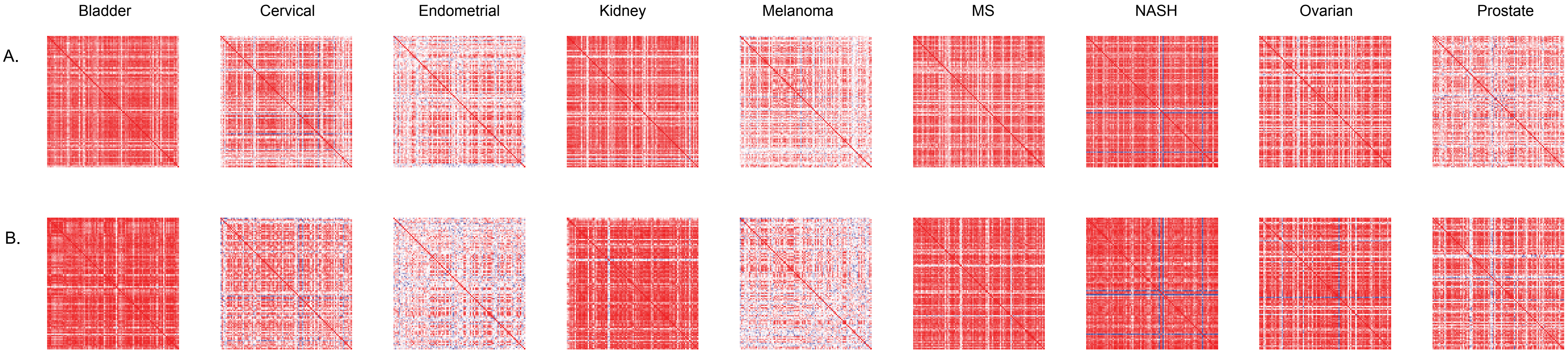

Figure S2. Venn Diagrams Illustrating Significantly Altered Proteins Across Various Diseases.
The shared proteins are presented separately for overrepresented proteins (A, B) and underrepresented proteins in urine (C).

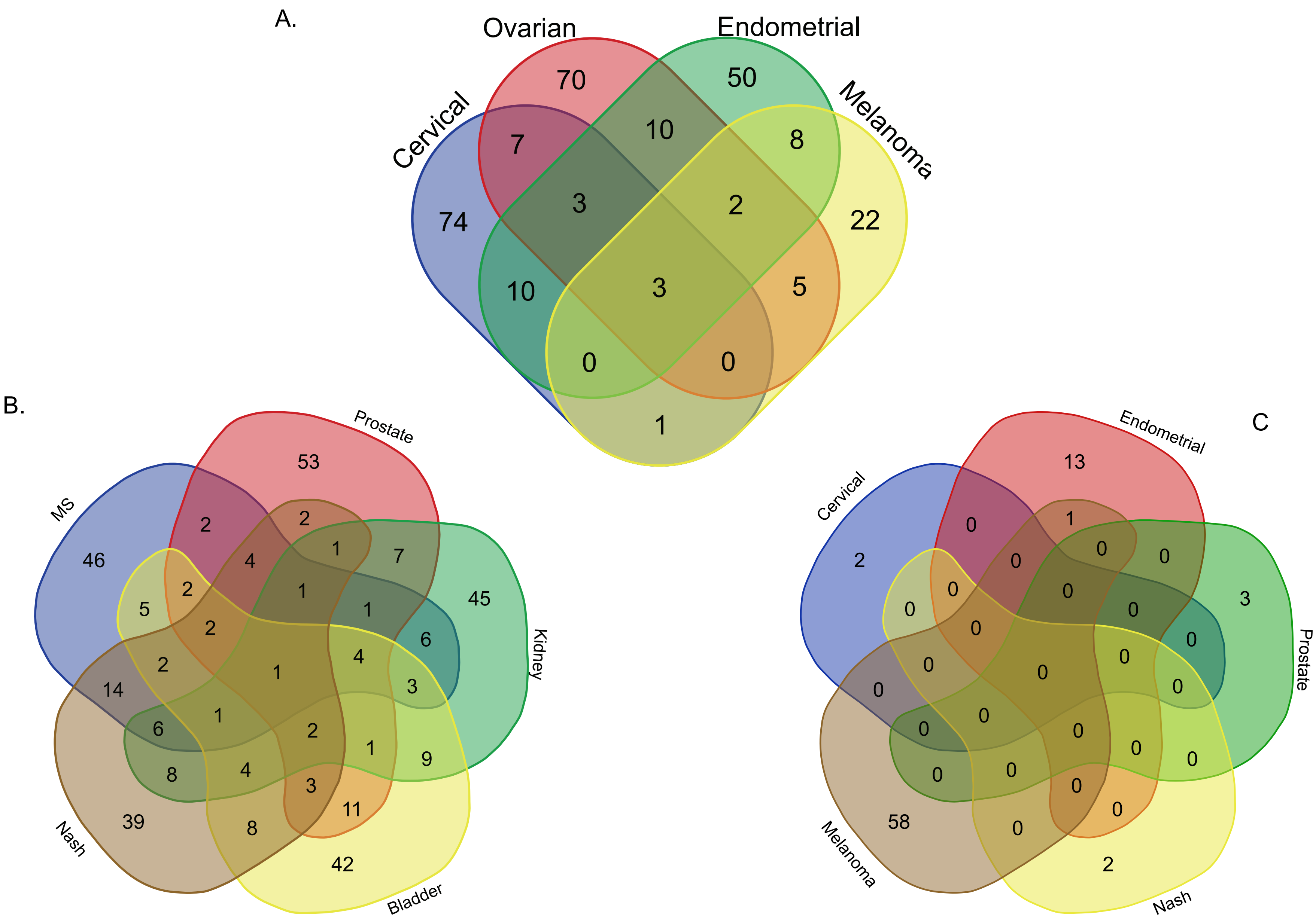

| | | | | | | | | |
|---|---|---|---|---|---|---|---|---|
| P22004 | Cardiometabolic | 0.001 | 97.7 | 12500 | 200000 | 2.1 | 13 | 16 |
| P05231 | Cardiometabolic | 0.4 | 0.8 | 3125 | 12500 | 3.6 | 6 | 12 |
| P46379 | Cardiometabolic | 390.6 | 781.3 | 200000 | 400000 | 2.4 | 8 | 10 |
| P40818 | Cardiometabolic | 3125.0 | 6250.0 | 800000 | 800000 | 2.1 | 8 | 13 |
| P62736 | Cardiometabolic | | | | | | 7 | 8 |
| P51161 | Cardiometabolic | | | | | | 10 | 24 |
| P09237 | Cardiometabolic | 97.7 | 97.7 | 6250 | 50000 | 1.8 | 6 | 8 |
| Q15165 | Cardiometabolic | | | | | | 8 | 8 |
| Q92558 | Cardiometabolic | 6250.0 | 12500.0 | 800000 | 1600000 | 1.8 | 10 | 19 |
| O43186 | Cardiometabolic | | | | | | 8 | 13 |
| P08670 | Cardiometabolic | 12500.0 | 12500.0 | 1600000 | 3200000 | 2.1 | 6 | 60 |
| P07585 | Cardiometabolic | 195.3 | 390.6 | 100000 | 200000 | 2.4 | 7 | 11 |
| Q15831 | Cardiometabolic | 390.6 | 781.3 | 400000 | 800000 | 2.7 | 10 | 21 |
| P19429 | Cardiometabolic | 97.7 | 195.3 | 25000 | 200000 | 2.1 | 15 | 14 |
| Q9UKP3 | Cardiometabolic | 195.3 | 781.3 | 100000 | 200000 | 2.1 | 9 | 18 |
| O95988 | Cardiometabolic | 97.7 | 195.3 | 200000 | 800000 | 3.0 | 8 | 24 |
| P36952 | Cardiometabolic | 781.3 | 1562.5 | 100000 | 200000 | 1.8 | 11 | 21 |
| Q16619 | Cardiometabolic | 195.3 | 781.3 | 100000 | 400000 | 2.1 | 11 | 22 |
| P61978 | Cardiometabolic | 97.7 | 97.7 | 50000 | 200000 | 2.7 | 8 | 12 |
| P17676 | Cardiometabolic | | | | | | 11 | 41 |
| Q96N03 | Cardiometabolic | | | | | | 8 | 11 |
| Q13105 | Cardiometabolic | 6.1 | 12.2 | 6250 | 200000 | 2.7 | 8 | 8 |
| O95684 | Cardiometabolic | 781.3 | 1562.5 | 100000 | 3200000 | 1.8 | 10 | 24 |
| P21246 | Cardiometabolic | | | | | | 20 | 17 |
| P34998 | Cardiometabolic | 24.4 | 48.8 | 6250 | 25000 | 2.1 | 7 | 24 |
| Q6UWL2 | Cardiometabolic | 48.8 | 97.7 | 12500 | 200000 | 2.1 | 8 | 14 |
| Q969D9 | Cardiometabolic | 6.1 | 48.8 | 12500 | 200000 | 2.4 | 7 | 14 |
| P35218 | Cardiometabolic | 1.5 | 6.1 | 6250 | 25000 | 3.0 | 7 | 9 |
| P55082 | Cardiometabolic | 24.4 | 48.8 | 100000 | 800000 | 3.3 | 9 | 26 |
| P17516 | Cardiometabolic | 390.6 | 781.3 | 100000 | 400000 | 2.1 | 12 | 17 |
| O15354 | Cardiometabolic | 781.3 | 1562.5 | 100000 | 400000 | 1.8 | 9 | 11 |
| Q12912 | Cardiometabolic | 48.8 | 97.7 | 12500 | 50000 | 2.1 | 8 | 12 |
| P31997 | Cardiometabolic | | | | | | 9 | 12 |

| | | | | | | | | |
|---|---|---|---|---|---|---|---|---|
| Q9NRV9 | Cardiometabolic | | | | | | 8 | 12 |
| Q9Y2B0 | Cardiometabolic | | | | | | 7 | 10 |
| O95183 | Cardiometabolic | 390.6 | 781.3 | 200000 | 400000 | 2.4 | 8 | 30 |
| P13807 | Cardiometabolic | 12.2 | 48.8 | 25000 | 200000 | 2.7 | 8 | 16 |
| P20718 | Cardiometabolic | | | | | | 8 | 12 |
| Q9H5Y7 | Cardiometabolic | 24.4 | 48.8 | 100000 | 400000 | 3.3 | 7 | 14 |
| Q8NC01 | Cardiometabolic | 97.7 | 195.3 | 25000 | 400000 | 2.1 | 7 | 10 |
| O75356 | Cardiometabolic | 48.8 | 97.7 | 100000 | 800000 | 3.0 | 8 | 8 |
| Q96A56 | Cardiometabolic | 97.7 | 195.3 | 25000 | 200000 | 2.1 | 8 | 8 |
| Q9GZM7 | Cardiometabolic | 1.5 | 3.1 | 6250 | 50000 | 3.3 | 10 | 6 |
| P27352 | Cardiometabolic | 0.2 | 0.4 | 6250 | 50000 | 4.2 | 9 | 6 |
| P12104 | Cardiometabolic | 0.4 | 1.5 | 781 | 6250 | 2.7 | 9 | 8 |
| Q9NQX5 | Cardiometabolic | 6.1 | 12.2 | 3125 | 50000 | 2.4 | 10 | 6 |
| P12724 | Cardiometabolic | | | | | | 12 | 18 |
| Q9UBU3 | Cardiometabolic | 24.4 | 48.8 | 50000 | 200000 | 3.0 | 8 | 13 |
| P35754 | Cardiometabolic | | | | | | 9 | 11 |
| P41159 | Cardiometabolic | 12.2 | 48.8 | 12500 | 200000 | 2.4 | 7 | 11 |
| P09382 | Cardiometabolic | 24.4 | 48.8 | 50000 | 200000 | 3.0 | 12 | 15 |
| P40189 | Cardiometabolic | 390.6 | 781.3 | 200000 | 800000 | 2.4 | 7 | 7 |
| Q92692 | Cardiometabolic | 0.1 | 0.1 | 195 | 781 | 3.3 | 9 | 6 |
| Q15067 | Cardiometabolic | | | | | | 9 | 15 |
| Q16620 | Cardiometabolic | 48.8 | 48.8 | 6250 | 50000 | 2.1 | 10 | 10 |
| P21583 | Cardiometabolic | 3.1 | 6.1 | 12500 | 50000 | 3.3 | 10 | 8 |
| P31431 | Cardiometabolic | 1.5 | 3.1 | 3125 | 200000 | 3.0 | 9 | 6 |
| P09417 | Cardiometabolic | 12.2 | 24.4 | 50000 | 800000 | 3.3 | 7 | 7 |
| Q8WVQ1 | Cardiometabolic | 6.1 | 12.2 | 6250 | 50000 | 2.7 | 8 | 5 |
| Q15846 | Cardiometabolic | 6.1 | 12.2 | 6250 | 50000 | 2.7 | 9 | 12 |
| Q9UKJ0 | Cardiometabolic | 0.8 | 1.5 | 3125 | 12500 | 3.3 | 7 | 6 |
| O00161 | Cardiometabolic | 6.1 | 12.2 | 12500 | 50000 | 3.0 | 12 | 13 |
| Q6WN34 | Cardiometabolic | 195.3 | 390.6 | 50000 | 200000 | 2.1 | 10 | 7 |
| Q92823 | Cardiometabolic | 6.1 | 12.2 | 6250 | 12500 | 2.7 | 15 | 16 |
| P00568 | Cardiometabolic | | | | | | 9 | 10 |
| Q13043 | Cardiometabolic | 195.3 | 390.6 | 6250 | 50000 | 1.2 | 6 | 28 |

| | | | | | | | | |
|---|---|---|---|---|---|---|---|---|
| P09525 | Cardiometabolic | 97.7 | 195.3 | 25000 | 800000 | 2.1 | 7 | 14 |
| Q05315 | Cardiometabolic | 390.6 | 781.3 | 50000 | 800000 | 1.8 | 10 | 8 |
| Q9UHL4 | Cardiometabolic | 97.7 | 195.3 | 50000 | 400000 | 2.4 | 11 | 15 |
| Q03154 | Cardiometabolic | 390.6 | 781.3 | 400000 | 800000 | 2.7 | 10 | 11 |
| P10644 | Cardiometabolic | 24.4 | 48.8 | 100000 | 200000 | 3.3 | 12 | 12 |
| O94903 | Cardiometabolic | 12.2 | 24.4 | 12500 | 50000 | 2.7 | 9 | 12 |
| P16234 | Cardiometabolic | 12.2 | 48.8 | 6250 | 50000 | 2.1 | 10 | 6 |
| Q9H773 | Cardiometabolic | 12.2 | 24.4 | 12500 | 50000 | 2.7 | 9 | 9 |
| O14917 | Cardiometabolic | 24.4 | 48.8 | 50000 | 400000 | 3.0 | 13 | 10 |
| Q9H7M9 | Cardiometabolic | 1.5 | 3.1 | 1563 | 6250 | 2.7 | 10 | 14 |
| NT-proBNP | Cardiometabolic | 97.7 | 195.3 | 50000 | 200000 | 2.4 | 11 | 9 |
| P31949 | Cardiometabolic | 6.1 | 6.1 | 3125 | 12500 | 2.7 | 14 | 10 |
| Q9Y4X3 | Cardiometabolic | | | | | | 17 | 14 |
| P01222 | Cardiometabolic | 1.5 | 3.1 | 1563 | 3125 | 2.7 | 11 | 7 |
| P21980 | Cardiometabolic | 6.1 | 24.4 | 50000 | 200000 | 3.3 | 15 | 11 |
| P21549 | Cardiometabolic | 390.6 | 781.3 | 1600000 | 12800000 | 3.3 | 9 | 10 |
| Q9UMF0 | Cardiometabolic | 12.2 | 24.4 | 6250 | 50000 | 2.4 | 9 | 9 |
| Q6GTS8 | Cardiometabolic | 48.8 | 97.7 | 100000 | 800000 | 3.0 | 11 | 13 |
| Q9NY25 | Cardiometabolic | 0.1 | 0.4 | 3125 | 6250 | 3.9 | 9 | 8 |
| Q9HBB8 | Cardiometabolic | 12.2 | 48.8 | 25000 | 200000 | 2.7 | 10 | 7 |
| P16112 | Cardiometabolic | 12.2 | 24.4 | 50000 | 200000 | 3.3 | 12 | 9 |
| P55285 | Cardiometabolic | 97.7 | 195.3 | 25000 | 200000 | 2.1 | 12 | 10 |
| O60664 | Cardiometabolic | 195.3 | 390.6 | 50000 | 800000 | 2.1 | 12 | 13 |
| P08263 | Cardiometabolic | 3.1 | 6.1 | 12500 | 400000 | 3.3 | 9 | 7 |
| P52888 | Cardiometabolic | 6.1 | 12.2 | 6250 | 200000 | 2.7 | 9 | 5 |
| Q969P0 | Cardiometabolic | 12.2 | 12.2 | 25000 | 200000 | 3.3 | 9 | 9 |
| O75340 | Cardiometabolic | 1562.5 | 1562.5 | 100000 | 800000 | 1.8 | 8 | 10 |
| Q9ULL4 | Cardiometabolic | 24.4 | 48.8 | 50000 | 200000 | 3.0 | 10 | 13 |
| P41218 | Cardiometabolic | 390.6 | 781.3 | 800000 | 800000 | 3.0 | 9 | 10 |
| P48357 | Cardiometabolic | | | | | | 8 | 12 |
| Q9Y286 | Cardiometabolic | 3.1 | 3.1 | 6250 | 50000 | 3.3 | 8 | 8 |
| P51693 | Cardiometabolic | 195.3 | 195.3 | 50000 | 400000 | 2.4 | 12 | 15 |
| O95502 | Cardiometabolic | 6.1 | 12.2 | 25000 | 50000 | 3.3 | 10 | 7 |

| | | | | | | | | |
|---|---|---|---|---|---|---|---|---|
| O75791 | Cardiometabolic | 1562.5 | 3125.0 | 400000 | 800000 | 2.1 | 8 | 10 |
| Q06418 | Cardiometabolic | 3.1 | 6.1 | 1563 | 6250 | 2.4 | 9 | 10 |
| Q12864 | Cardiometabolic | 195.3 | 390.6 | 50000 | 400000 | 2.1 | 14 | 13 |
| Q9Y5X1 | Cardiometabolic | 390.6 | 781.3 | 400000 | 800000 | 2.7 | 9 | 11 |
| Q13541 | Cardiometabolic | 24.4 | 24.4 | 12500 | 50000 | 2.7 | 8 | 8 |
| Q9UHD0 | Cardiometabolic | 3.1 | 12.2 | 100000 | 400000 | 3.9 | 9 | 7 |
| Q8WX77 | Cardiometabolic | 0.001 | 1.5 | 3125 | 6250 | 3.3 | 9 | 9 |
| Q8WTU2 | Cardiometabolic | 6.1 | 6.1 | 6250 | 200000 | 3.0 | 7 | 8 |
| P78380 | Cardiometabolic | 3.1 | 3.1 | 3125 | 12500 | 3.0 | 9 | 8 |
| Q99674 | Cardiometabolic | 24.4 | 48.8 | 6250 | 50000 | 2.1 | 8 | 7 |
| Q8NI22 | Cardiometabolic | 48.8 | 97.7 | 12500 | 400000 | 2.1 | 9 | 7 |
| P23526 | Cardiometabolic | 195.3 | 390.6 | 100000 | 400000 | 2.4 | 8 | 13 |
| Q8IW75 | Cardiometabolic | 24.4 | 97.7 | 6250 | 50000 | 1.8 | 9 | 14 |
| P09601 | Cardiometabolic | 3.1 | 3.1 | 6250 | 25000 | 3.3 | 9 | 13 |
| Q9BQR3 | Cardiometabolic | 12.2 | 24.4 | 25000 | 50000 | 3.0 | 8 | 6 |
| Q6PJW8 | Cardiometabolic | 12.2 | 48.8 | 6250 | 50000 | 2.1 | 10 | 21 |
| Q8IZP9 | Cardiometabolic | 12.2 | 48.8 | 12500 | 50000 | 2.4 | 10 | 8 |
| P06858 | Cardiometabolic | 195.3 | 195.3 | 400000 | 800000 | 3.3 | 8 | 11 |
| Q13158 | Cardiometabolic | 781.3 | 3125.0 | 800000 | 800000 | 2.4 | 9 | 10 |
| Q9NR28 | Cardiometabolic | 3.1 | 6.1 | 12500 | 50000 | 3.3 | 12 | 28 |
| Q86VZ4 | Cardiometabolic | 12.2 | 48.8 | 6250 | 200000 | 2.1 | 11 | 7 |
| P35247 | Cardiometabolic | 6.1 | 12.2 | 6250 | 50000 | 2.7 | 8 | 5 |
| O95544 | Cardiometabolic | 62.5 | 125.0 | 256000 | 512000 | 3.3 | 10 | 6 |
| Q14956 | Cardiometabolic | 24.4 | 48.8 | 12500 | 200000 | 2.4 | 12 | 10 |
| P18827 | Cardiometabolic | 6.1 | 12.2 | 6250 | 200000 | 2.7 | 14 | 12 |
| P10145 | Cardiometabolic | 0.1 | 0.2 | 781 | 6250 | 3.6 | 10 | 15 |
| Q53H82 | Cardiometabolic | 48.8 | 97.7 | 25000 | 200000 | 2.4 | 8 | 9 |
| Q9BUD6 | Cardiometabolic | 24.4 | 48.8 | 25000 | 400000 | 2.7 | 9 | 8 |
| Q16820 | Cardiometabolic | 97.7 | 195.3 | 25000 | 50000 | 2.1 | 9 | 9 |
| Q9Y5K6 | Cardiometabolic | 6.1 | 12.2 | 12500 | 50000 | 3.0 | 10 | 9 |
| P41236 | Cardiometabolic | 12.2 | 24.4 | 12500 | 50000 | 2.7 | 9 | 10 |
| Q13275 | Cardiometabolic | 148.4 | 296.9 | 38000 | 304000 | 2.1 | 9 | 8 |
| Q96LA6 | Cardiometabolic | 97.7 | 195.3 | 400000 | 800000 | 3.3 | 10 | 7 |

| | | | | | | | | |
|---|---|---|---|---|---|---|---|---|
| P19022 | Cardiometabolic | 390.6 | 781.3 | 200000 | 800000 | 2.4 | 11 | 9 |
| P00797 | Cardiometabolic | 6.1 | 12.2 | 12500 | 50000 | 3.0 | 9 | 9 |
| Q8N1Q1 | Cardiometabolic | 6.1 | 24.4 | 12500 | 50000 | 2.7 | 8 | 15 |
| Q07108 | Cardiometabolic | 0.8 | 3.1 | 12500 | 50000 | 3.6 | 12 | 14 |
| Q9UK05 | Cardiometabolic | 1.5 | 6.1 | 12500 | 50000 | 3.3 | 10 | 8 |
| O95841 | Cardiometabolic | 97.7 | 195.3 | 100000 | 400000 | 2.7 | 9 | 7 |
| Q9UEW3 | Cardiometabolic | 24.4 | 24.4 | 50000 | 800000 | 3.3 | 7 | 6 |
| P02462 | Cardiometabolic | 0.8 | 3.1 | 50000 | 200000 | 4.2 | 10 | 6 |
| P07204 | Cardiometabolic | 3.1 | 6.1 | 6250 | 50000 | 3.0 | 9 | 8 |
| P01241 | Cardiometabolic | | | | | | 7 | 15 |
| A6NI73 | Cardiometabolic | 0.8 | 1.5 | 3125 | 6250 | 3.3 | 8 | 7 |
| Q01973 | Cardiometabolic | 24.4 | 48.8 | 3125 | 12500 | 1.8 | 10 | 7 |
| Q16773 | Cardiometabolic | 0.8 | 1.5 | 3125 | 12500 | 3.3 | 12 | 8 |
| P09467 | Cardiometabolic | 97.7 | 195.3 | 50000 | 200000 | 2.4 | 8 | 40 |
| P42830 | Cardiometabolic | 3.1 | 6.1 | 3125 | 6250 | 2.7 | 11 | 10 |
| Q9BQB4 | Cardiometabolic | 6.1 | 6.1 | 25000 | 50000 | 3.6 | 8 | 11 |
| Q76M96 | Cardiometabolic | 24.4 | 48.8 | 50000 | 800000 | 3.0 | 10 | 8 |
| P19971 | Cardiometabolic | 390.6 | 781.3 | 200000 | 800000 | 2.4 | 9 | 17 |
| Q92520 | Cardiometabolic | 3.1 | 6.1 | 6250 | 50000 | 3.0 | 8 | 7 |
| P07711 | Cardiometabolic | 24.4 | 97.7 | 50000 | 200000 | 2.7 | 10 | 9 |
| P04792 | Cardiometabolic | | | | | | 15 | 28 |
| Q99523 | Cardiometabolic | 24.4 | 24.4 | 50000 | 400000 | 3.3 | 9 | 8 |
| P20711 | Cardiometabolic | 3.1 | 6.1 | 25000 | 50000 | 3.6 | 8 | 7 |
| O60496 | Cardiometabolic | 6.1 | 12.2 | 25000 | 200000 | 3.3 | 10 | 15 |
| P07911 | Cardiometabolic | 1.5 | 3.1 | 12500 | 100000 | 3.6 | 4 | 3 |
| Q13361 | Cardiometabolic | 12.2 | 24.4 | 12500 | 100000 | 2.7 | 6 | 9 |
| P00750 | Cardiometabolic | 0.8 | 3.1 | 25000 | 100000 | 3.9 | 8 | 8 |
| O75326 | Cardiometabolic | 0.8 | 1.5 | 12500 | 50000 | 3.9 | 5 | 5 |
| P23141 | Cardiometabolic | 24.4 | 48.8 | 400000 | 800000 | 3.9 | 5 | 7 |
| P22748 | Cardiometabolic | 0.8 | 1.5 | 12500 | 100000 | 3.9 | 5 | 6 |
| P55058 | Cardiometabolic | 6.1 | 12.2 | 25000 | 400000 | 3.3 | 5 | 11 |
| P01130 | Cardiometabolic | 0.8 | 1.5 | 25000 | 100000 | 4.2 | 4 | 14 |
| P13598 | Cardiometabolic | 97.7 | 195.3 | 200000 | 400000 | 3.0 | 6 | 9 |

| | | | | | | | | |
|---|---|---|---|---|---|---|---|---|
| Q8NBP7 | Cardiometabolic | 97.7 | 195.3 | 400000 | 800000 | 3.3 | 5 | 28 |
| P15090 | Cardiometabolic | 48.8 | 195.3 | 50000 | 200000 | 2.4 | 5 | 5 |
| Q76LX8 | Cardiometabolic | 6.1 | 12.2 | 6250 | 100000 | 2.7 | 6 | 9 |
| P08833 | Cardiometabolic | 1.5 | 3.1 | 12500 | 100000 | 3.6 | 3 | 3 |
| P33151 | Cardiometabolic | 195.3 | 390.6 | 200000 | 400000 | 2.7 | 5 | 8 |
| Q16270 | Cardiometabolic | 12.2 | 24.4 | 12500 | 200000 | 2.7 | 7 | 8 |
| P54760 | Cardiometabolic | 1.5 | 3.1 | 12500 | 100000 | 3.6 | 4 | 5 |
| Q96AP7 | Cardiometabolic | 3.1 | 6.1 | 3125 | 12500 | 2.7 | 4 | 5 |
| P32942 | Cardiometabolic | 1.5 | 1.5 | 3125 | 12500 | 3.3 | 3 | 4 |
| P08118 | Cardiometabolic | 0.001 | 6.1 | 6250 | 12500 | 3.0 | 3 | 5 |
| Q06141 | Cardiometabolic | 0.4 | 0.8 | 1563 | 12500 | 3.3 | 4 | 5 |
| P01589 | Cardiometabolic | 0.02 | 0.0 | 391 | 3125 | 3.9 | 5 | 5 |
| P07858 | Cardiometabolic | 12.2 | 24.4 | 12500 | 25000 | 2.7 | 4 | 4 |
| Q9UBP4 | Cardiometabolic | 6.1 | 12.2 | 25000 | 100000 | 3.3 | 5 | 5 |
| Q86U17 | Cardiometabolic | 3.1 | 6.1 | 12500 | 100000 | 3.3 | 4 | 14 |
| P04066 | Cardiometabolic | 6.1 | 12.2 | 50000 | 200000 | 3.6 | 6 | 13 |
| Q14767 | Cardiometabolic | 6.1 | 12.2 | 25000 | 200000 | 3.3 | 4 | 5 |
| Q9NQ79 | Cardiometabolic | 6.1 | 24.4 | 100000 | 800000 | 3.6 | 6 | 8 |
| O14798 | Cardiometabolic | 0.4 | 0.8 | 6250 | 12500 | 3.9 | 5 | 5 |
| Q5VY43 | Cardiometabolic | 12.2 | 24.4 | 12500 | 100000 | 2.7 | 5 | 4 |
| P48304 | Cardiometabolic | 0.8 | 1.5 | 1563 | 12500 | 3.0 | 4 | 4 |
| P15085 | Cardiometabolic | 0.4 | 0.8 | 3125 | 12500 | 3.6 | 6 | 5 |
| Q07507 | Cardiometabolic | 3.1 | 6.1 | 6250 | 400000 | 3.0 | 6 | 7 |
| P17931 | Cardiometabolic | 12.2 | 48.8 | 12500 | 50000 | 2.4 | 5 | 5 |
| P04275 | Cardiometabolic | 1.5 | 3.1 | 12500 | 100000 | 3.6 | 9 | 32 |
| P55808 | Cardiometabolic | 1.5 | 3.1 | 6250 | 12500 | 3.3 | 6 | 10 |
| Q03167 | Cardiometabolic | 12.2 | 24.4 | 100000 | 200000 | 3.6 | 5 | 8 |
| P14555 | Cardiometabolic | | | | | | 6 | 9 |
| Q9Y275 | Cardiometabolic | 0.2 | 0.8 | 12500 | 50000 | 4.2 | 4 | 4 |
| P08581 | Cardiometabolic | 6.1 | 12.2 | 12500 | 50000 | 3.0 | 4 | 6 |
| Q9H2A7 | Cardiometabolic | 0.8 | 1.5 | 12500 | 50000 | 3.9 | 5 | 6 |
| Q9UM47 | Cardiometabolic | 1.5 | 3.1 | 6250 | 50000 | 3.3 | 6 | 8 |
| P07451 | Cardiometabolic | 1562.5 | 12500.0 | 12800000 | 12800000 | 3.0 | 4 | 5 |

| | | | | | | | | |
|---|---|---|---|---|---|---|---|---|
| P09619 | Cardiometabolic | 3.1 | 6.1 | 3125 | 12500 | 2.7 | 4 | 4 |
| P80370 | Cardiometabolic | 0.8 | 3.1 | 6250 | 25000 | 3.3 | 4 | 5 |
| Q14162 | Cardiometabolic | 0.8 | 1.5 | 3125 | 12500 | 3.3 | 5 | 6 |
| Q99988 | Cardiometabolic | 1.5 | 3.1 | 6250 | 25000 | 3.3 | 5 | 4 |
| P04080 | Cardiometabolic | 3.1 | 6.1 | 3125 | 12500 | 2.7 | 4 | 3 |
| P02144 | Cardiometabolic | 0.8 | 0.8 | 195 | 3125 | 2.4 | 4 | 5 |
| Q13822 | Cardiometabolic | 390.6 | 781.3 | 100000 | 800000 | 2.1 | 4 | 5 |
| P08236 | Cardiometabolic | 3.1 | 6.1 | 6250 | 50000 | 3.0 | 4 | 6 |
| Q01638 | Cardiometabolic | 0.8 | 3.1 | 3125 | 12500 | 3.0 | 4 | 4 |
| Q13740 | Cardiometabolic | 3.1 | 6.1 | 3125 | 12500 | 2.7 | 4 | 5 |
| P48960 | Cardiometabolic | 3.1 | 6.1 | 25000 | 200000 | 3.6 | 5 | 7 |
| P17813 | Cardiometabolic | 1.5 | 3.1 | 3125 | 12500 | 3.0 | 4 | 4 |
| P31146 | Cardiometabolic | 6.1 | 24.4 | 100000 | 400000 | 3.6 | 6 | 7 |
| P12111 | Cardiometabolic | 3.1 | 6.1 | 3125 | 100000 | 2.7 | 4 | 6 |
| P16581 | Cardiometabolic | 0.8 | 1.5 | 1563 | 3125 | 3.0 | 5 | 5 |
| P15086 | Cardiometabolic | 1.5 | 3.1 | 6250 | 25000 | 3.3 | 4 | 5 |
| Q15828 | Cardiometabolic | 12.2 | 24.4 | 3125 | 12500 | 2.1 | 5 | 5 |
| Q9NNX6 | Cardiometabolic | 1.5 | 3.1 | 12500 | 100000 | 3.6 | 4 | 6 |
| P04054 | Cardiometabolic | 0.4 | 0.8 | 6250 | 12500 | 3.9 | 4 | 6 |
| Q9H1U4 | Cardiometabolic | 0.8 | 1.5 | 6250 | 25000 | 3.6 | 5 | 6 |
| P19021 | Cardiometabolic | 97.7 | 195.3 | 200000 | 800000 | 3.0 | 4 | 6 |
| P48745 | Cardiometabolic | 0.001 | 1.5 | 6250 | 25000 | 3.6 | 3 | 5 |
| P20062 | Cardiometabolic | 0.8 | 1.5 | 6250 | 12500 | 3.6 | 5 | 9 |
| O75023 | Cardiometabolic | 0.4 | 0.8 | 6250 | 25000 | 3.9 | 4 | 4 |
| P18065 | Cardiometabolic | 12.2 | 24.4 | 50000 | 200000 | 3.3 | 5 | 7 |
| O00584 | Cardiometabolic | 0.4 | 0.8 | 3125 | 12500 | 3.6 | 4 | 4 |
| P19961 | Cardiometabolic | 12.2 | 24.4 | 6250 | 25000 | 2.4 | 8 | 10 |
| Q12860 | Cardiometabolic | 3.1 | 6.1 | 12500 | 100000 | 3.3 | 5 | 6 |
| Q13231 | Cardiometabolic | 0.4 | 0.8 | 6250 | 25000 | 3.9 | 5 | 5 |
| P39060 | Cardiometabolic | 3.1 | 6.1 | 3125 | 12500 | 2.7 | 4 | 8 |
| P25445 | Cardiometabolic | 0.1 | 0.2 | 6250 | 12500 | 4.5 | 5 | 6 |
| P23284 | Cardiometabolic | 24.4 | 48.8 | 12500 | 200000 | 2.4 | 8 | 10 |
| O15467 | Cardiometabolic | 3.1 | 3.1 | 6250 | 12500 | 3.3 | 4 | 8 |

| | | | | | | | | |
|---|---|---|---|---|---|---|---|---|
| Q13867 | Cardiometabolic | 24.4 | 48.8 | 12500 | 100000 | 2.4 | 4 | 5 |
| Q13332 | Cardiometabolic | 3.1 | 6.1 | 6250 | 12500 | 3.0 | 5 | 5 |
| P19957 | Cardiometabolic | 12.2 | 48.8 | 6250 | 12500 | 2.1 | 5 | 5 |
| P35590 | Cardiometabolic | | | | | | 4 | 6 |
| P09093 | Cardiometabolic | 1.5 | 6.1 | 6250 | 25000 | 3.0 | 4 | 7 |
| P46531 | Cardiometabolic | 0.2 | 0.4 | 6250 | 12500 | 4.2 | 3 | 4 |
| P04746 | Cardiometabolic | 24.4 | 24.4 | 6250 | 12500 | 2.4 | 8 | 8 |
| P78324 | Cardiometabolic | 0.4 | 0.8 | 6250 | 12500 | 3.9 | 5 | 7 |
| P04085 | Cardiometabolic | 0.4 | 0.8 | 3125 | 12500 | 3.6 | 4 | 6 |
| Q9HD89 | Cardiometabolic | 0.2 | 0.4 | 3125 | 12500 | 3.9 | 6 | 6 |
| O15031 | Cardiometabolic | 0.8 | 1.5 | 6250 | 50000 | 3.6 | 4 | 5 |
| P24158 | Cardiometabolic | 12.2 | 48.8 | 6250 | 100000 | 2.1 | 5 | 4 |
| P05107 | Cardiometabolic | 1.5 | 3.1 | 6250 | 25000 | 3.3 | 6 | 8 |
| P13987 | Cardiometabolic | 0.001 | 6.1 | 1563 | 3125 | 2.4 | 4 | 6 |
| O75594 | Cardiometabolic | 0.8 | 1.5 | 3125 | 12500 | 3.3 | 5 | 6 |
| Q12884 | Cardiometabolic | 12.2 | 24.4 | 12500 | 25000 | 2.7 | 5 | 6 |
| P05121 | Cardiometabolic | 0.8 | 1.5 | 6250 | 25000 | 3.6 | 6 | 5 |
| P00533 | Cardiometabolic | 3.1 | 6.1 | 6250 | 12500 | 3.0 | 4 | 6 |
| P13686 | Cardiometabolic | 0.8 | 1.5 | 6250 | 12500 | 3.6 | 4 | 7 |
| P02452 | Cardiometabolic | | | | | | 3 | 4 |
| P20160 | Cardiometabolic | 6.1 | 12.2 | 6250 | 12500 | 2.7 | 7 | 9 |
| P42574 | Cardiometabolic | 1.5 | 3.1 | 3125 | 12500 | 3.0 | 5 | 9 |
| P10451 | Cardiometabolic | 1.5 | 3.1 | 6250 | 12500 | 3.3 | 3 | 8 |
| Q16769 | Cardiometabolic | 24.4 | 48.8 | 6250 | 50000 | 2.1 | 5 | 7 |
| Q14393 | Cardiometabolic | 48.8 | 97.7 | 400000 | 800000 | 3.6 | 5 | 7 |
| P42785 | Cardiometabolic | 24.4 | 24.4 | 6250 | 25000 | 2.4 | 5 | 6 |
| Q8TDL5 | Cardiometabolic | 97.7 | 390.6 | 200000 | 800000 | 2.7 | 5 | 8 |
| Q16663 | Cardiometabolic | 3.1 | 6.1 | 6250 | 12500 | 3.0 | 8 | 5 |
| Q8N423 | Cardiometabolic | 0.8 | 1.5 | 6250 | 12500 | 3.6 | 4 | 6 |
| P10586 | Cardiometabolic | 3.1 | 6.1 | 25000 | 200000 | 3.6 | 4 | 7 |
| Q9Y4L1 | Cardiometabolic | 24.4 | 24.4 | 12500 | 400000 | 2.7 | 5 | 6 |
| P15907 | Cardiometabolic | 24.4 | 48.8 | 400000 | 800000 | 3.9 | 6 | 9 |
| Q8NHL6 | Cardiometabolic | 0.8 | 1.5 | 3125 | 12500 | 3.3 | 4 | 5 |

| | | | | | | | | |
|---|---|---|---|---|---|---|---|---|
| P43121 | Cardiometabolic | 24.4 | 48.8 | 25000 | 800000 | 2.7 | 8 | 7 |
| P00740 | Cardiometabolic | 48.8 | 97.7 | 50000 | 800000 | 2.7 | 7 | 9 |
| P12830 | Cardiometabolic | 6.1 | 6.1 | 12500 | 50000 | 3.3 | 9 | 12 |
| P15529 | Cardiometabolic | 390.6 | 3125.0 | 6400000 | 12800000 | 3.3 | 11 | 13 |
| P13591 | Cardiometabolic | 6.1 | 12.2 | 25000 | 400000 | 3.3 | 7 | 9 |
| P12318 | Cardiometabolic | 0.2 | 0.4 | 1563 | 6250 | 3.6 | 10 | 10 |
| Q9UBR2 | Cardiometabolic | 0.8 | 1.5 | 3125 | 6250 | 3.3 | 7 | 9 |
| P18428 | Cardiometabolic | | | | | | 11 | 24 |
| Q12794 | Cardiometabolic | 24.4 | 24.4 | 25000 | 100000 | 3.0 | 7 | 6 |
| P07478 | Cardiometabolic | 0.4 | 0.8 | 391 | 12500 | 2.7 | 7 | 6 |
| P07359 | Cardiometabolic | 1.5 | 3.1 | 12500 | 25000 | 3.6 | 7 | 11 |
| P98160 | Cardiometabolic | 0.4 | 1.5 | 6250 | 25000 | 3.6 | 5 | 9 |
| P08887 | Cardiometabolic | 0.1 | 0.2 | 3125 | 6250 | 4.2 | 7 | 10 |
| P59665 | Cardiometabolic | | | | | | 12 | 22 |
| P24821 | Cardiometabolic | 0.4 | 1.5 | 3125 | 25000 | 3.3 | 9 | 10 |
| P16109 | Cardiometabolic | 0.2 | 0.4 | 3125 | 12500 | 3.9 | 9 | 11 |
| Q14515 | Cardiometabolic | 6.1 | 12.2 | 12500 | 50000 | 3.0 | 9 | 9 |
| Q86VB7 | Cardiometabolic | 6.1 | 12.2 | 25000 | 100000 | 3.3 | 8 | 7 |
| O95998 | Cardiometabolic | 0.8 | 1.5 | 12500 | 25000 | 3.9 | 7 | 10 |
| P20023 | Cardiometabolic | 0.4 | 0.8 | 12500 | 25000 | 4.2 | 6 | 8 |
| Q9NZK5 | Cardiometabolic | 0.4 | 0.8 | 6250 | 50000 | 3.9 | 6 | 7 |
| Q13508 | Cardiometabolic | 1.5 | 3.1 | 3125 | 25000 | 3.0 | 7 | 7 |
| Q15485 | Cardiometabolic | 12.2 | 12.2 | 25000 | 800000 | 3.3 | 8 | 11 |
| P80188 | Cardiometabolic | 0.8 | 1.5 | 1563 | 25000 | 3.0 | 8 | 9 |
| P30530 | Cardiometabolic | 0.4 | 0.8 | 6250 | 25000 | 3.9 | 8 | 7 |
| Q99650 | Cardiometabolic | 6.1 | 12.2 | 12500 | 25000 | 3.0 | 7 | 9 |
| Q15113 | Cardiometabolic | 24.4 | 48.8 | 12500 | 25000 | 2.4 | 9 | 13 |
| O14786 | Cardiometabolic | 6.1 | 12.2 | 12500 | 25000 | 3.0 | 8 | 8 |
| Q96KN2 | Cardiometabolic | 390.6 | 781.3 | 400000 | 800000 | 2.7 | 8 | 15 |
| Q6EMK4 | Cardiometabolic | 6.1 | 12.2 | 12500 | 50000 | 3.0 | 9 | 9 |
| P19320 | Cardiometabolic | 1.5 | 3.1 | 12500 | 50000 | 3.6 | 6 | 8 |
| P00441 | Cardiometabolic | 24.4 | 97.7 | 6250 | 25000 | 1.8 | 5 | 19 |
| O75015 | Cardiometabolic | 3.1 | 6.1 | 6250 | 25000 | 3.0 | 7 | 7 |

| | | | | | | | | |
|---|---|---|---|---|---|---|---|---|
| P07339 | Cardiometabolic | 48.8 | 195.3 | 25000 | 50000 | 2.1 | | |
| Q16853 | Cardiometabolic | 6.1 | 12.2 | 6250 | 25000 | 2.7 | 8 | 8 |
| P15144 | Cardiometabolic | 24.4 | 48.8 | 100000 | 800000 | 3.3 | 7 | 8 |
| P08174 | Cardiometabolic | 0.2 | 0.4 | 3125 | 6250 | 3.9 | 8 | 9 |
| O00533 | Cardiometabolic | 24.4 | 48.8 | 12500 | 400000 | 2.4 | 7 | 8 |
| P02786 | Cardiometabolic | 24.4 | 48.8 | 200000 | 800000 | 3.6 | 7 | 6 |
| P05556 | Cardiometabolic | 0.05 | 0.4 | 12500 | 25000 | 4.5 | 11 | 13 |
| P10646 | Cardiometabolic | 0.8 | 1.5 | 6250 | 50000 | 3.6 | 10 | 13 |
| Q9BXJ1 | Cardiometabolic | 0.8 | 1.5 | 6250 | 50000 | 3.6 | 17 | 19 |
| Q9NPY3 | Cardiometabolic | 0.2 | 0.4 | 6250 | 25000 | 4.2 | 7 | 8 |
| P10721 | Cardiometabolic | 0.4 | 0.8 | 1563 | 6250 | 3.3 | 8 | 10 |
| P14543 | Cardiometabolic | 3.1 | 6.1 | 50000 | 200000 | 3.9 | 7 | 8 |
| O95445 | Cardiometabolic | 6.1 | 6.1 | 12500 | 50000 | 3.3 | 11 | 12 |
| Q96H15 | Cardiometabolic | 0.2 | 0.4 | 1563 | 6250 | 3.6 | 8 | 8 |
| P08571 | Cardiometabolic | 12.2 | 24.4 | 200000 | 400000 | 3.9 | 10 | 14 |
| Q99969 | Cardiometabolic | 6.1 | 24.4 | 25000 | 800000 | 3.0 | 8 | 12 |
| A1L4H1 | Cardiometabolic | 48.8 | 97.7 | 50000 | 400000 | 2.7 | 9 | 10 |
| Q07654 | Cardiometabolic | 0.2 | 0.4 | 781 | 3125 | 3.3 | 10 | 10 |
| P35443 | Cardiometabolic | 6.1 | 24.4 | 50000 | 200000 | 3.3 | 7 | 9 |
| P55774 | Cardiometabolic | 0.4 | 0.8 | 3125 | 6250 | 3.6 | 10 | 23 |
| Q9Y5C1 | Cardiometabolic | 1.5 | 3.1 | 12500 | 100000 | 3.6 | 10 | 9 |
| Q16627 | Cardiometabolic | 0.1 | 0.4 | 1563 | 6250 | 3.6 | 7 | 11 |
| P08709 | Cardiometabolic | 0.8 | 3.1 | 6250 | 25000 | 3.3 | 6 | 10 |
| P41222 | Cardiometabolic | 3.1 | 6.1 | 12500 | 50000 | 3.3 | 7 | 8 |
| P06681 | Cardiometabolic | 48.8 | 97.7 | 200000 | 800000 | 3.3 | 7 | 11 |
| P24592 | Cardiometabolic | 0.8 | 1.5 | 12500 | 25000 | 3.9 | 6 | 8 |
| Q15582 | Cardiometabolic | 97.7 | 195.3 | 100000 | 400000 | 2.7 | 8 | 12 |
| P36222 | Cardiometabolic | 0.4 | 0.8 | 3125 | 6250 | 3.6 | 6 | 10 |
| Q06033 | Cardiometabolic | 48.8 | 97.7 | 50000 | 100000 | 2.7 | 7 | 10 |
| Q9UGM5 | Cardiometabolic | 12.2 | 48.8 | 50000 | 200000 | 3.0 | 7 | 7 |
| P49747 | Cardiometabolic | 1.5 | 3.1 | 12500 | 50000 | 3.6 | 8 | 9 |
| Q92820 | Cardiometabolic | 195.3 | 390.6 | 50000 | 400000 | 2.1 | 6 | 7 |
| P00915 | Cardiometabolic | 0.8 | 1.5 | 25000 | 100000 | 4.2 | 6 | 7 |

| | | | | | | | | |
|---|---|---|---|---|---|---|---|---|
| P13501 | Cardiometabolic | 0.4 | 1.5 | 1563 | 12500 | 3.0 | 10 | 14 |
| P05451 | Cardiometabolic | 0.05 | 0.1 | 3125 | 6250 | 4.5 | 6 | 7 |
| Q12805 | Cardiometabolic | 24.4 | 48.8 | 50000 | 100000 | 3.0 | 10 | 14 |
| P03950 | Cardiometabolic | 0.8 | 1.5 | 781 | 3125 | 2.7 | 7 | 11 |
| P27487 | Cardiometabolic | 12.2 | 48.8 | 12500 | 100000 | 2.4 | 6 | 8 |
| P04070 | Cardiometabolic | 97.7 | 195.3 | 100000 | 400000 | 2.7 | 7 | 8 |
| P05362 | Cardiometabolic | 0.4 | 0.8 | 6250 | 25000 | 3.9 | 8 | 8 |
| P01034 | Cardiometabolic | 12.2 | 48.8 | 6250 | 12500 | 2.1 | 6 | 13 |
| P17936 | Cardiometabolic | 12.2 | 24.4 | 12500 | 25000 | 2.7 | 8 | 9 |
| P01033 | Cardiometabolic | 0.8 | 1.5 | 3125 | 12500 | 3.3 | 9 | 10 |
| P14902 | Cardiometabolic | 390.6 | 1562.5 | 200000 | 400000 | 2.1 | 10 | 13 |
| Q14160 | Cardiometabolic | 195.3 | 390.6 | 50000 | 200000 | 2.1 | 8 | 22 |
| P12829 | Cardiometabolic | 3125.0 | 3125.0 | 100000 | 800000 | 1.5 | 11 | 20 |
| Q9BY49 | Cardiometabolic II | | | | | | 6 | 7 |
| Q9NZN3 | Cardiometabolic | 6250.0 | 12500.0 | 800000 | 800000 | 1.8 | 6 | 27 |
| Q96C92 | Cardiometabolic | 781.3 | 781.3 | 400000 | 800000 | 2.7 | 9 | 15 |
| Q5SW79 | Cardiometabolic | 48.8 | 48.8 | 3125 | 6250 | 1.8 | 6 | 11 |
| O75506 | Cardiometabolic | 12.2 | 12.2 | 3125 | 25000 | 2.4 | 6 | 29 |
| Q15477 | Cardiometabolic II | | | | | | 7 | 25 |
| P04141 | Cardiometabolic | 12.2 | 24.4 | 12500 | 50000 | 2.7 | 16 | 18 |
| P21817 | Cardiometabolic | 390.6 | 781.3 | 100000 | 200000 | 2.1 | 13 | 12 |
| A6BM72 | Cardiometabolic | 97.7 | 97.7 | 25000 | 200000 | 2.4 | 10 | 10 |
| O00291 | Cardiometabolic | 1562.5 | 3125.0 | 200000 | 400000 | 1.8 | 8 | |
| Q8IZC4 | Cardiometabolic | 3125.0 | 6250.0 | 400000 | 800000 | 1.8 | 5 | |
| O60701 | Cardiometabolic II | | | | | | 9 | 24 |
| O14958 | Cardiometabolic | 6250.0 | 6250.0 | 400000 | 1600000 | 1.8 | 12 | 21 |
| E2RYF7 | Cardiometabolic | 24.4 | 48.8 | 12500 | 50000 | 2.4 | 6 | 16 |
| Q9NVZ3 | Cardiometabolic | 12500.0 | 12500.0 | 400000 | 800000 | 1.5 | 15 | 25 |
| P23634 | Cardiometabolic | 390.6 | 390.6 | 25000 | 400000 | 1.8 | | |
| Q9Y4C8 | Cardiometabolic | 390.6 | 390.6 | 100000 | 400000 | 2.4 | 14 | 18 |
| Q9Y623 | Cardiometabolic | 390.6 | 390.6 | 25000 | 200000 | 1.8 | 14 | |
| P54709 | Cardiometabolic | 781.3 | 781.3 | 100000 | 400000 | 2.1 | | |
| Q07973 | Cardiometabolic | 1562.5 | 3125.0 | 200000 | 800000 | 1.8 | 15 | |

| | | | | | | | | |
|---|---|---|---|---|---|---|---|---|
| P48507 | Cardiometabolic | 1562.5 | 3125.0 | 200000 | 800000 | 1.8 | 11 | 17 |
| P06753 | Cardiometabolic | 1562.5 | 1562.5 | 200000 | 400000 | 2.1 | 11 | 12 |
| Q04695 | Cardiometabolic | 390.6 | 781.3 | 200000 | 800000 | 2.4 | | |
| P25391 | Cardiometabolic II | | | | | | | |
| Q15059 | Cardiometabolic II | | | | | | 12 | 32 |
| O00567 | Cardiometabolic | 3125.0 | 3125.0 | 200000 | 400000 | 1.8 | 13 | 37 |
| Q9NZJ5 | Cardiometabolic | 3125.0 | 6250.0 | 400000 | 800000 | 1.8 | 17 | 15 |
| P35228 | Cardiometabolic | 390.6 | 781.3 | 400000 | 800000 | 2.7 | 15 | 17 |
| Q13503 | Cardiometabolic | 781.3 | 1562.5 | 100000 | 400000 | 1.8 | 9 | 17 |
| P08913 | Cardiometabolic | 3125.0 | 6250.0 | 200000 | 400000 | 1.5 | 9 | 15 |
| P33121 | Cardiometabolic | 3125.0 | 6250.0 | 200000 | 400000 | 1.5 | | |
| Q9BY32 | Cardiometabolic | 781.3 | 1562.5 | 50000 | 400000 | 1.5 | 8 | 21 |
| P30049 | Cardiometabolic | 390.6 | 390.6 | 50000 | 200000 | 2.1 | 10 | 20 |
| P10109 | Cardiometabolic II | | | | | | 9 | |
| P55011 | Cardiometabolic II | | | | | | 8 | |
| Q01780 | Cardiometabolic | 3125.0 | 6250.0 | 400000 | 800000 | 1.8 | 12 | 3 |
| Q6UWF7 | Cardiometabolic | 1562.5 | 1562.5 | 200000 | 400000 | 2.1 | 7 | 14 |
| Q9Y3B8 | Cardiometabolic | 97.7 | 195.3 | 12500 | 400000 | 1.8 | 14 | |
| A6NCE7 | Cardiometabolic | 3125.0 | 3125.0 | 200000 | 800000 | 1.8 | 8 | 15 |
| Q08499 | Cardiometabolic | 25000.0 | 50000.0 | 3200000 | 12800000 | 1.8 | 10 | 50 |
| P46783 | Cardiometabolic II | | | | | | | |
| Q96DA2 | Cardiometabolic | 3125.0 | 3125.0 | 200000 | 800000 | 1.8 | 13 | 16 |
| P49755 | Cardiometabolic | 390.6 | 781.3 | 100000 | 200000 | 2.1 | 7 | 12 |
| Q96HD9 | Cardiometabolic II | | | | | | 10 | 9 |
| B6SEH8 | Cardiometabolic | 195.3 | 781.3 | 100000 | 200000 | 2.1 | 15 | 33 |
| O43734 | Cardiometabolic | 195.3 | 195.3 | 100000 | 200000 | 2.7 | 10 | 11 |
| O95180 | Cardiometabolic | 390.6 | 781.3 | 100000 | 400000 | 2.1 | 12 | 20 |
| Q9H2M3 | Cardiometabolic | 50000.0 | 100000.0 | 6400000 | 12800000 | 1.8 | | |
| P06729 | Cardiometabolic | 781.3 | 1562.5 | 200000 | 400000 | 2.1 | 10 | 14 |
| Q96IW2 | Cardiometabolic | 6250.0 | 6250.0 | 400000 | 800000 | 1.8 | 12 | 8 |
| P55769 | Cardiometabolic II | | | | | | 8 | 12 |
| Q9Y2W1 | Cardiometabolic | 3125.0 | 3125.0 | 200000 | 800000 | 1.8 | 9 | 15 |
| O95858 | Cardiometabolic | 390.6 | 781.3 | 100000 | 200000 | 2.1 | 15 | 14 |

| | | | | | | | | |
|---|---|---|---|---|---|---|---|---|
| Q9H347 | Cardiometabolic | 390.6 | 390.6 | 100000 | 200000 | 2.4 | | |
| P78524 | Cardiometabolic | 3125.0 | 3125.0 | 200000 | 400000 | 1.8 | 11 | 6 |
| Q14353 | Cardiometabolic II | | | | | | 12 | |
| Q15370 | Cardiometabolic | 1562.5 | 1562.5 | 100000 | 400000 | 1.8 | 10 | 32 |
| P20929 | Cardiometabolic | 781.3 | 1562.5 | 200000 | 400000 | 2.1 | 13 | 8 |
| Q9BW61 | Cardiometabolic | 1171.9 | 1171.9 | 300000 | 600000 | 2.4 | 11 | 11 |
| Q5TA50 | Cardiometabolic | 6250.0 | 12500.0 | 400000 | 800000 | 1.5 | 7 | 17 |
| O15305 | Cardiometabolic II | | | | | | 11 | 39 |
| P05026 | Cardiometabolic | 195.3 | 195.3 | 25000 | 200000 | 2.1 | 11 | 11 |
| Q86UW2 | Cardiometabolic | 781.3 | 781.3 | 100000 | 400000 | 2.1 | 10 | 22 |
| P38935 | Cardiometabolic | 390.6 | 781.3 | 50000 | 200000 | 1.8 | | |
| Q14088 | Cardiometabolic | 6250.0 | 12500.0 | 800000 | 800000 | 1.8 | 12 | 21 |
| Q9Y2Y0 | Cardiometabolic | 1562.5 | 3125.0 | 200000 | 400000 | 1.8 | 9 | 17 |
| Q8WZ42 | Cardiometabolic | 195.3 | 390.6 | 12500 | 200000 | 1.5 | 10 | 12 |
| P12270 | Cardiometabolic | 97.7 | 97.7 | 12500 | 100000 | 2.1 | 14 | 16 |
| O75521 | Cardiometabolic | 6250.0 | 12500.0 | 400000 | 800000 | 1.5 | 13 | 35 |
| P05976 | Cardiometabolic | 6250.0 | 6250.0 | 200000 | 400000 | 1.5 | 6 | 14 |
| P14415 | Cardiometabolic | 781.3 | 1562.5 | 100000 | 400000 | 1.8 | | |
| Q9UFP1 | Cardiometabolic II | | | | | | 12 | 15 |
| Q9BZC7 | Cardiometabolic | 781.3 | 1562.5 | 200000 | 400000 | 2.1 | 19 | 26 |
| Q6NZY4 | Cardiometabolic | 48.8 | 97.7 | 50000 | 100000 | 2.7 | 18 | 30 |
| Q9NYX4 | Cardiometabolic | 3125.0 | 3125.0 | 200000 | 800000 | 1.8 | 6 | 15 |
| P16066 | Cardiometabolic | 3125.0 | 6250.0 | 200000 | 800000 | 1.5 | 8 | 9 |
| Q99707 | Cardiometabolic | 781.3 | 781.3 | 50000 | 400000 | 1.8 | 20 | 10 |
| Q8N8E3 | Cardiometabolic | 195.3 | 390.6 | 100000 | 200000 | 2.4 | 8 | 10 |
| P37058 | Cardiometabolic | 3125.0 | 3125.0 | 200000 | 800000 | 1.8 | 12 | 25 |
| Q92935 | Cardiometabolic | 390.6 | 390.6 | 100000 | 200000 | 2.4 | 10 | 14 |
| P21673 | Cardiometabolic | 195.3 | 195.3 | 12500 | 100000 | 1.8 | 2 | |
| O43290 | Cardiometabolic | 390.6 | 1562.5 | 100000 | 400000 | 1.8 | 11 | 15 |
| Q96K76 | Cardiometabolic | 390.6 | 781.3 | 100000 | 400000 | 2.1 | | |
| Q13296 | Cardiometabolic | 1562.5 | 3125.0 | 200000 | 400000 | 1.8 | 23 | |
| Q6P4F2 | Cardiometabolic | 195.3 | 195.3 | 12500 | 200000 | 1.8 | 13 | 18 |
| P05000 | Cardiometabolic | 195.3 | 390.6 | 200000 | 400000 | 2.7 | 4 | 1 |

| | | | | | | | | |
|---|---|---|---|---|---|---|---|---|
| P57078 | Cardiometabolic | 1562.5 | 1562.5 | 200000 | 800000 | 2.1 | 15 | 31 |
| Q9UKX7 | Cardiometabolic II | | | | | | 11 | |
| Q02127 | Cardiometabolic | 12500.0 | 12500.0 | 800000 | 800000 | 1.8 | 10 | |
| Q6ZN66 | Cardiometabolic II | | | | | | | |
| Q9BV94 | Cardiometabolic | 1562.5 | 3125.0 | 200000 | 800000 | 1.8 | 13 | 6 |
| Q07075 | Cardiometabolic | 390.6 | 781.3 | 100000 | 400000 | 2.1 | 13 | 21 |
| P23511 | Cardiometabolic II | | | | | | 11 | 10 |
| Q96LB8 | Cardiometabolic | 390.6 | 390.6 | 50000 | 100000 | 2.1 | 8 | 25 |
| Q8NET8 | Cardiometabolic | 24.4 | 24.4 | 12500 | 50000 | 2.7 | 14 | 23 |
| Q9NV35 | Cardiometabolic | 390.6 | 390.6 | 25000 | 200000 | 1.8 | 14 | 19 |
| Q16774 | Cardiometabolic II | | | | | | 13 | 3 |
| Q16836 | Cardiometabolic II | | | | | | | |
| P54296 | Cardiometabolic | 97.7 | 195.3 | 12500 | 100000 | 1.8 | 18 | 17 |
| Q9BZL6 | Cardiometabolic | 6250.0 | 12500.0 | 800000 | 3200000 | 1.8 | 9 | 10 |
| Q10587 | Cardiometabolic | 195.3 | 781.3 | 200000 | 800000 | 2.4 | 14 | 11 |
| A6NHS7 | Cardiometabolic | 390.6 | 390.6 | 25000 | 200000 | 1.8 | 10 | 33 |
| Q15018 | Cardiometabolic II | | | | | | 9 | 18 |
| O00425 | Cardiometabolic | 12500.0 | 12500.0 | 200000 | 800000 | 1.2 | | |
| Q9UNN8 | Cardiometabolic II | | | | | | 8 | 26 |
| Q14807 | Cardiometabolic II | | | | | | 15 | 37 |
| P35606 | Cardiometabolic | 195.3 | 195.3 | 6250 | 400000 | 1.5 | 16 | 32 |
| P20382 | Cardiometabolic | 3125.0 | 3125.0 | 100000 | 800000 | 1.5 | 8 | 12 |
| Q96PU4 | Cardiometabolic | 781.3 | 1562.5 | 100000 | 200000 | 1.8 | | |
| P00966 | Cardiometabolic | 781.3 | 3125.0 | 200000 | 400000 | 1.8 | 9 | 18 |
| P48668 | Cardiometabolic | 390.6 | 390.6 | 25000 | 200000 | 1.8 | | |
| O00327 | Cardiometabolic | 3125.0 | 6250.0 | 400000 | 800000 | 1.8 | | |
| O95670 | Cardiometabolic II | | | | | | 22 | 17 |
| P50461 | Cardiometabolic | 1562.5 | 3125.0 | 50000 | 200000 | 1.2 | 24 | 22 |
| Q3SXY8 | Cardiometabolic | 195.3 | 195.3 | 25000 | 400000 | 2.1 | 13 | 24 |
| Q03013 | Cardiometabolic | 3125.0 | 3125.0 | 200000 | 800000 | 1.8 | 12 | 12 |
| O43896 | Cardiometabolic | 97.7 | 97.7 | 25000 | 50000 | 2.4 | 9 | 10 |
| P59901 | Cardiometabolic II | | | | | | 12 | |
| Q01484 | Cardiometabolic | 195.3 | 390.6 | 50000 | 100000 | 2.1 | 11 | 11 |

| | | | | | | | | |
|---|---|---|---|---|---|---|---|---|
| P19838 | Cardiometabolic II | | | | | | 13 | 19 |
| P22033 | Cardiometabolic II | | | | | | 12 | 34 |
| Q12986 | Cardiometabolic | 1562.5 | 3125.0 | 100000 | 400000 | 1.5 | 14 | 22 |
| Q01581 | Cardiometabolic | 6250.0 | 6250.0 | 200000 | 400000 | 1.5 | 8 | 6 |
| O94766 | Cardiometabolic II | | | | | | | |
| Q14781 | Cardiometabolic | 3125.0 | 3125.0 | 400000 | 800000 | 2.1 | 10 | |
| Q96A35 | Cardiometabolic | 3125.0 | 6250.0 | 400000 | 800000 | 1.8 | 11 | 16 |
| Q58F21 | Cardiometabolic | 48.8 | 97.7 | 12500 | 50000 | 2.1 | 13 | 20 |
| Q8NFP7 | Cardiometabolic | 781.3 | 781.3 | 50000 | 200000 | 1.8 | | |
| P46926 | Cardiometabolic | 195.3 | 195.3 | 12500 | 100000 | 1.8 | 9 | 12 |
| Q9UBV2 | Cardiometabolic | 390.6 | 781.3 | 100000 | 400000 | 2.1 | 7 | 8 |
| Q5JTV8 | Cardiometabolic | 6250.0 | 12500.0 | 800000 | 3200000 | 1.8 | 10 | 26 |
| Q8ND90 | Cardiometabolic | 6250.0 | 12500.0 | 800000 | 800000 | 1.8 | 9 | 24 |
| P32241 | Cardiometabolic | 195.3 | 390.6 | 100000 | 800000 | 2.4 | | |
| P35609 | Cardiometabolic | 781.3 | 1562.5 | 100000 | 400000 | 1.8 | 18 | 18 |
| O75427 | Cardiometabolic II | | | | | | 17 | 17 |
| Q93052 | Cardiometabolic II | | | | | | 14 | 15 |
| Q86VR7 | Cardiometabolic | 6250.0 | 6250.0 | 200000 | 800000 | 1.5 | 10 | |
| P41227 | Cardiometabolic II | | | | | | 20 | 35 |
| Q5W0V3 | Cardiometabolic II | | | | | | 8 | 7 |
| Q86VP3 | Cardiometabolic | 97.7 | 195.3 | 25000 | 200000 | 2.1 | 13 | 25 |
| Q99598 | Cardiometabolic II | | | | | | 11 | 20 |
| Q13563 | Cardiometabolic | 781.3 | 1562.5 | 100000 | 200000 | 1.8 | 12 | 17 |
| O75534 | Cardiometabolic II | | | | | | 16 | 20 |
| A6NDB9 | Cardiometabolic II | | | | | | 12 | 19 |
| Q5VVQ6 | Cardiometabolic II | | | | | | 14 | 21 |
| Q96EU7 | Cardiometabolic II | | | | | | 16 | 26 |
| P55010 | Cardiometabolic | 781.3 | 1562.5 | 100000 | 400000 | 1.8 | 16 | 27 |
| Q9Y2L6 | Cardiometabolic II | | | | | | 11 | 19 |
| P13224 | Cardiometabolic II | | | | | | 15 | 28 |
| P0C7L1 | Cardiometabolic | 781.3 | 1562.5 | 200000 | 400000 | 2.1 | 10 | |
| O15018 | Cardiometabolic | 390.6 | 390.6 | 200000 | 400000 | 2.7 | 14 | 20 |
| P10082 | Cardiometabolic II | | | | | | 14 | 18 |

| | | | | | | | | |
|---|---|---|---|---|---|---|---|---|
| Q7Z7H5 | Cardiometabolic | 1562.5 | 1562.5 | 100000 | 400000 | 1.8 | 11 | |
| Q16206 | Cardiometabolic | 1562.5 | 3125.0 | 800000 | 3200000 | 2.4 | 8 | 27 |
| P29536 | Cardiometabolic | 195.3 | 390.6 | 200000 | 800000 | 2.7 | 7 | 24 |
| Q14324 | Cardiometabolic | 781.3 | 1562.5 | 100000 | 800000 | 1.8 | 7 | 21 |
| Q96ID5 | Cardiometabolic | 1562.5 | 1562.5 | 100000 | 400000 | 1.8 | 7 | 15 |
| P13929 | Cardiometabolic II | | | | | | | |
| P20645 | Cardiometabolic | 97.7 | 97.7 | 6250 | 25000 | 1.8 | 9 | 12 |
| P23327 | Cardiometabolic | 781.3 | 1562.5 | 200000 | 3200000 | 2.1 | 8 | 16 |
| Q9H173 | Cardiometabolic | 390.6 | 781.3 | 50000 | 200000 | 1.8 | 6 | 9 |
| Q9BTK6 | Cardiometabolic | 195.3 | 195.3 | 50000 | 400000 | 2.4 | 15 | 17 |
| P01225 | Cardiometabolic | 97.7 | 195.3 | 12500 | 100000 | 1.8 | 6 | 9 |
| Q8TER0 | Cardiometabolic | 1562.5 | 3125.0 | 400000 | 800000 | 2.1 | 12 | 21 |
| Q0VD83 | Cardiometabolic II | | | | | | 7 | 12 |
| O95980 | Cardiometabolic | 1562.5 | 1562.5 | 50000 | 200000 | 1.5 | 7 | 5 |
| Q13316 | Cardiometabolic | 3125.0 | 6250.0 | 800000 | 6400000 | 2.1 | 8 | 35 |
| P50053 | Cardiometabolic | 195.3 | 195.3 | 12500 | 100000 | 1.8 | 6 | 20 |
| Q14457 | Cardiometabolic | 12500.0 | 25000.0 | 800000 | 800000 | 1.5 | 10 | 14 |
| Q99942 | Cardiometabolic II | | | | | | 12 | 21 |
| I3L3R5 | Cardiometabolic | 1562.5 | 1562.5 | 100000 | 400000 | 1.8 | 9 | 12 |
| Q99807 | Cardiometabolic | 390.6 | 781.3 | 100000 | 400000 | 2.1 | 12 | 18 |
| Q53T59 | Cardiometabolic | 3125.0 | 3125.0 | 100000 | 800000 | 1.5 | 8 | 25 |
| Q8N668 | Cardiometabolic | 24.4 | 48.8 | 12500 | 50000 | 2.4 | 8 | 16 |
| P55809 | Cardiometabolic | 781.3 | 3125.0 | 200000 | 800000 | 1.8 | 10 | 21 |
| O75348 | Cardiometabolic | 3125.0 | 3125.0 | 200000 | 800000 | 1.8 | 12 | 35 |
| P11532 | Cardiometabolic | 390.6 | 781.3 | 200000 | 800000 | 2.4 | 9 | 17 |
| Q9Y5X3 | Cardiometabolic II | | | | | | 5 | 9 |
| P05305 | Cardiometabolic II | | | | | | 6 | 15 |
| Q8WZ75 | Cardiometabolic II | | | | | | 6 | 9 |
| Q8IVF2 | Cardiometabolic | 97.7 | 195.3 | 12500 | 50000 | 1.8 | 6 | 9 |
| P35914 | Cardiometabolic | 781.3 | 1562.5 | 100000 | 400000 | 1.8 | 8 | 25 |
| Q14643 | Cardiometabolic | 781.3 | 1562.5 | 100000 | 400000 | 1.8 | 13 | 14 |
| Q9BQI0 | Cardiometabolic | 390.6 | 781.3 | 200000 | 800000 | 2.4 | 7 | 19 |
| P36776 | Cardiometabolic | 781.3 | 781.3 | 50000 | 200000 | 1.8 | 10 | 28 |

| | | | | | | | | |
|---|---|---|---|---|---|---|---|---|
| Q9H7C9 | Cardiometabolic II | | | | | | 8 | 13 |
| O14841 | Cardiometabolic II | | | | | | 7 | 24 |
| Q8WXC3 | Cardiometabolic | 6.1 | 12.2 | 1563 | 12500 | 2.1 | 7 | 11 |
| O75061 | Cardiometabolic II | | | | | | 6 | 30 |
| Q8NC42 | Cardiometabolic | 97.7 | 97.7 | 6250 | 50000 | 1.8 | 9 | 11 |
| Q8TAE8 | Cardiometabolic II | | | | | | 10 | 25 |
| Q5GAN6 | Cardiometabolic | 195.3 | 195.3 | 12500 | 100000 | 1.8 | 7 | 13 |
| P35520 | Cardiometabolic | 3125.0 | 3125.0 | 800000 | 800000 | 2.4 | 9 | 25 |
| P30084 | Cardiometabolic II | | | | | | 8 | 27 |
| Q8WUF8 | Cardiometabolic | 390.6 | 781.3 | 100000 | 400000 | 2.1 | 8 | 20 |
| O43423 | Cardiometabolic | 97.7 | 195.3 | 50000 | 400000 | 2.4 | 1 | |
| Q13137 | Cardiometabolic | 195.3 | 195.3 | 50000 | 200000 | 2.4 | 8 | 15 |
| O94979 | Cardiometabolic II | | | | | | 7 | 31 |
| Q16621 | Cardiometabolic II | | | | | | 8 | 12 |
| Q9H3K6 | Cardiometabolic | 195.3 | 195.3 | 25000 | 200000 | 2.1 | 7 | 11 |
| P07098 | Cardiometabolic | 195.3 | 390.6 | 50000 | 200000 | 2.1 | 7 | 22 |
| P21754 | Cardiometabolic | 48.8 | 97.7 | 25000 | 50000 | 2.4 | | |
| P07492 | Cardiometabolic II | | | | | | 6 | 22 |
| P20042 | Cardiometabolic II | | | | | | 11 | 25 |
| O60476 | Cardiometabolic | 781.3 | 781.3 | 100000 | 800000 | 2.1 | 5 | 8 |
| Q9NYZ4 | Cardiometabolic | 12.2 | 12.2 | 6250 | 25000 | 2.7 | 6 | 11 |
| Q09666 | Cardiometabolic | 97.7 | 195.3 | 50000 | 200000 | 2.4 | 7 | 14 |
| Q92835 | Cardiometabolic | 3125.0 | 3125.0 | 200000 | 800000 | 1.8 | 8 | 12 |
| P43487 | Cardiometabolic II | | | | | | 7 | 14 |
| Q6ZRY4 | Cardiometabolic | 48.8 | 97.7 | 6250 | 12500 | 1.8 | 6 | 30 |
| P07355 | Cardiometabolic | 6250.0 | 12500.0 | 800000 | 800000 | 1.8 | 9 | 14 |
| Q6YN16 | Cardiometabolic II | | | | | | 8 | 12 |
| Q9UJ70 | Cardiometabolic | 195.3 | 390.6 | 12500 | 100000 | 1.5 | 7 | 22 |
| O95825 | Cardiometabolic | 48.8 | 195.3 | 25000 | 50000 | 2.1 | 7 | 28 |
| Q24JP5 | Cardiometabolic II | | | | | | | |
| P02458 | Cardiometabolic | 97.7 | 97.7 | 12500 | 50000 | 2.1 | 6 | 15 |
| P09543 | Cardiometabolic | 781.3 | 781.3 | 100000 | 800000 | 2.1 | 6 | 14 |
| P50914 | Cardiometabolic | 50000.0 | 100000.0 | 6400000 | 12800000 | 1.8 | 14 | 18 |

| | | | | | | | | |
|---|---|---|---|---|---|---|---|---|
| Q7L266 | Cardiometabolic II | | | | | | 16 | 31 |
| P01189 | Cardiometabolic II | | | | | | | |
| Q8NFL0 | Cardiometabolic | 781.3 | 781.3 | 200000 | 400000 | 2.4 | 7 | 11 |
| Q96DR5 | Cardiometabolic | 1562.5 | 3125.0 | 100000 | 400000 | 1.5 | 7 | 32 |
| Q9HB40 | Cardiometabolic | 195.3 | 390.6 | 100000 | 400000 | 2.4 | 7 | 9 |
| Q8IWT1 | Cardiometabolic II | | | | | | 9 | 26 |
| Q5FWE3 | Cardiometabolic | 781.3 | 1562.5 | 200000 | 800000 | 2.1 | 6 | 11 |
| Q6UY14 | Cardiometabolic II | | | | | | 8 | 13 |
| Q9BV79 | Cardiometabolic | 6250.0 | 6250.0 | 400000 | 800000 | 1.8 | | |
| Q6UWR7 | Cardiometabolic | 12.2 | 24.4 | 6250 | 25000 | 2.4 | 8 | 22 |
| P07942 | Cardiometabolic | 195.3 | 390.6 | 50000 | 400000 | 2.1 | 6 | 12 |
| Q9NR61 | Cardiometabolic II | | | | | | 17 | 5 |
| P09681 | Cardiometabolic II | | | | | | 13 | 12 |
| P58107 | Cardiometabolic | 390.6 | 781.3 | 100000 | 400000 | 2.1 | 7 | 18 |
| Q12841 | Cardiometabolic II | | | | | | 5 | 9 |
| P0DPI2 | Cardiometabolic II | | | | | | | |
| P08590 | Cardiometabolic | 390.6 | 781.3 | 50000 | 800000 | 1.8 | 7 | 21 |
| Q86X76 | Cardiometabolic | 6250.0 | 6250.0 | 400000 | 800000 | 1.8 | 6 | 13 |
| Q96DC8 | Cardiometabolic | 24.4 | 48.8 | 25000 | 100000 | 2.7 | 4 | 14 |
| Q8TCD5 | Cardiometabolic | 12500.0 | 12500.0 | 800000 | 800000 | 1.8 | 5 | 21 |
| Q7Z7M9 | Cardiometabolic | 6250.0 | 6250.0 | 400000 | 800000 | 1.8 | 10 | 10 |
| Q00872 | Cardiometabolic | 781.3 | 1562.5 | 200000 | 800000 | 2.1 | 7 | 24 |
| Q14914 | Cardiometabolic | 390.6 | 390.6 | 50000 | 200000 | 2.1 | 5 | 17 |
| Q9UBQ7 | Cardiometabolic | 1562.5 | 1562.5 | 200000 | 800000 | 2.1 | 9 | 18 |
| Q9BVM4 | Cardiometabolic | 48.8 | 48.8 | 1563 | 12500 | 1.5 | 12 | 28 |
| Q12982 | Cardiometabolic | 3125.0 | 3125.0 | 100000 | 800000 | 1.5 | 11 | 13 |
| P33681 | Cardiometabolic II | | | | | | 17 | 18 |
| Q6UW49 | Cardiometabolic | 1562.5 | 3125.0 | 200000 | 800000 | 1.8 | 8 | 24 |
| P51511 | Cardiometabolic II | | | | | | 5 | 2 |
| Q9BW04 | Cardiometabolic | 97.7 | 195.3 | 12500 | 100000 | 1.8 | | |
| O14933 | Cardiometabolic | 1562.5 | 1562.5 | 50000 | 200000 | 1.5 | 7 | 11 |
| Q8WWV6 | Cardiometabolic | 6250.0 | 6250.0 | 400000 | 800000 | 1.8 | 6 | 22 |
| P23919 | Cardiometabolic | 3125.0 | 6250.0 | 400000 | 800000 | 1.8 | 5 | 21 |

| | | | | | | | | |
|---|---|---|---|---|---|---|---|---|
| O75711 | Cardiometabolic | 3125.0 | 3125.0 | 100000 | 400000 | 1.5 | 10 | 12 |
| Q6UXI7 | Cardiometabolic | 195.3 | 390.6 | 25000 | 100000 | 1.8 | 6 | 16 |
| P29692 | Cardiometabolic | 3125.0 | 3125.0 | 400000 | 800000 | 2.1 | 4 | 12 |
| P02008 | Cardiometabolic II | | | | | | 7 | 31 |
| Q9NQR4 | Cardiometabolic II | | | | | | 10 | 32 |
| Q9BQS7 | Cardiometabolic | 97.7 | 97.7 | 12500 | 50000 | 2.1 | 5 | 7 |
| Q9Y2E5 | Cardiometabolic | 390.6 | 781.3 | 50000 | 800000 | 1.8 | 4 | 12 |
| Q9H3S4 | Cardiometabolic | 24.4 | 48.8 | 12500 | 100000 | 2.4 | 5 | 8 |
| O43405 | Cardiometabolic | 195.3 | 195.3 | 25000 | 200000 | 2.1 | 5 | 9 |
| Q96C24 | Cardiometabolic II | | | | | | 13 | 26 |
| O60234 | Cardiometabolic | 781.3 | 781.3 | 50000 | 200000 | 1.8 | 6 | 23 |
| Q7Z304 | Cardiometabolic | 195.3 | 195.3 | 50000 | 200000 | 2.4 | 5 | 10 |
| P78539 | Cardiometabolic | 781.3 | 781.3 | 50000 | 200000 | 1.8 | 6 | 16 |
| Q9P2J2 | Cardiometabolic | 97.7 | 97.7 | 25000 | 100000 | 2.4 | 8 | 19 |
| Q8N4F0 | Cardiometabolic | 1562.5 | 3125.0 | 100000 | 200000 | 1.5 | | |
| P53674 | Cardiometabolic | 12.2 | 12.2 | 6250 | 25000 | 2.7 | 3 | 16 |
| P16035 | Cardiometabolic II | | | | | | 11 | 10 |
| Q8N436 | Cardiometabolic | 12500.0 | 12500.0 | 800000 | 800000 | 1.8 | 8 | 18 |
| Q13442 | Cardiometabolic | 3125.0 | 6250.0 | 200000 | 800000 | 1.5 | | |
| P14854 | Cardiometabolic II | | | | | | 10 | 20 |
| P23467 | Cardiometabolic | 781.3 | 3125.0 | 100000 | 200000 | 1.5 | 6 | 10 |
| Q13428 | Cardiometabolic | 390.6 | 781.3 | 50000 | 200000 | 1.8 | 7 | 9 |
| O75223 | Cardiometabolic | 195.3 | 390.6 | 25000 | 200000 | 1.8 | 9 | 29 |
| O75154 | Cardiometabolic | 48.8 | 97.7 | 25000 | 200000 | 2.4 | 6 | 27 |
| Q6NUS6 | Cardiometabolic | 1562.5 | 1562.5 | 100000 | 800000 | 1.8 | 6 | 12 |
| Q96EM0 | Cardiometabolic | 781.3 | 781.3 | 50000 | 200000 | 1.8 | 9 | 30 |
| Q96FZ7 | Cardiometabolic | 781.3 | 781.3 | 100000 | 800000 | 2.1 | 8 | 22 |
| Q969H8 | Cardiometabolic | 97.7 | 195.3 | 12500 | 200000 | 1.8 | 5 | 29 |
| P98161 | Cardiometabolic | 24.4 | 97.7 | 6250 | 25000 | 1.8 | 6 | 10 |
| Q9BXD5 | Cardiometabolic | 781.3 | 1562.5 | 100000 | 400000 | 1.8 | 5 | 18 |
| P54687 | Cardiometabolic | 1562.5 | 1562.5 | 100000 | 800000 | 1.8 | 8 | 10 |
| Q9BXN1 | Cardiometabolic II | | | | | | 8 | 26 |
| P51688 | Cardiometabolic | 97.7 | 97.7 | 12500 | 50000 | 2.1 | 4 | 6 |

| | | | | | | | | |
|---|---|---|---|---|---|---|---|---|
| O14960 | Cardiometabolic | 6250.0 | 6250.0 | 800000 | 800000 | 2.1 | 9 | 35 |
| P23471 | Cardiometabolic | 781.3 | 781.3 | 100000 | 200000 | 2.1 | 7 | 10 |
| P32320 | Cardiometabolic | 1562.5 | 3125.0 | 100000 | 800000 | 1.5 | 9 | 13 |
| P08138 | Cardiometabolic | 390.6 | 390.6 | 25000 | 100000 | 1.8 | 14 | 22 |
| Q6PI73 | Cardiometabolic | 97.7 | 195.3 | 25000 | 200000 | 2.1 | 6 | 21 |
| Q8NDI1 | Cardiometabolic | 195.3 | 390.6 | 25000 | 100000 | 1.8 | 8 | 20 |
| P08582 | Cardiometabolic | 97.7 | 195.3 | 50000 | 200000 | 2.4 | 5 | 8 |
| P52209 | Cardiometabolic | 6250.0 | 6250.0 | 200000 | 800000 | 1.5 | 8 | 19 |
| O43681 | Cardiometabolic | 1562.5 | 3125.0 | 400000 | 800000 | 2.1 | 8 | 26 |
| P15502 | Cardiometabolic II | | | | | | 11 | 16 |
| Q969X0 | Cardiometabolic | 195.3 | 390.6 | 50000 | 200000 | 2.1 | 6 | 35 |
| Q96MK3 | Cardiometabolic | 390.6 | 390.6 | 100000 | 800000 | 2.4 | 5 | 14 |
| Q8IZF2 | Cardiometabolic | 1562.5 | 3125.0 | 200000 | 800000 | 1.8 | 6 | 8 |
| Q96AG4 | Cardiometabolic II | | | | | | 12 | 29 |
| Q7Z7K0 | Cardiometabolic II | | | | | | 6 | 29 |
| P07093 | Cardiometabolic II | | | | | | 5 | 17 |
| P62072 | Cardiometabolic | 781.3 | 781.3 | 50000 | 200000 | 1.8 | 10 | 10 |
| P61026 | Cardiometabolic II | | | | | | 13 | 16 |
| P45954 | Cardiometabolic | 3125.0 | 3125.0 | 400000 | 800000 | 2.1 | 12 | 24 |
| Q6ZMM2 | Cardiometabolic | 390.6 | 390.6 | 50000 | 100000 | 2.1 | 11 | 19 |
| P05413 | Cardiometabolic II | | | | | | 8 | 16 |
| Q15388 | Cardiometabolic | 97.7 | 195.3 | 25000 | 100000 | 2.1 | 12 | 29 |
| Q9UBR1 | Cardiometabolic | 781.3 | 781.3 | 50000 | 200000 | 1.8 | 14 | 13 |
| P49593 | Cardiometabolic | 6250.0 | 6250.0 | 800000 | 800000 | 2.1 | 10 | 26 |
| O00194 | Cardiometabolic | 195.3 | 390.6 | 100000 | 400000 | 2.4 | | |
| P13667 | Cardiometabolic | 390.6 | 390.6 | 50000 | 100000 | 2.1 | 8 | 27 |
| P23560 | Cardiometabolic | 48.8 | 195.3 | 50000 | 100000 | 2.4 | 9 | 28 |
| P30046 | Cardiometabolic II | | | | | | 10 | 19 |
| Q86TH1 | Cardiometabolic | 3125.0 | 3125.0 | 400000 | 800000 | 2.1 | 9 | 12 |
| P02730 | Cardiometabolic II | | | | | | | |
| P13796 | Cardiometabolic | 1562.5 | 1562.5 | 400000 | 800000 | 2.4 | 10 | 19 |
| Q9Y303 | Cardiometabolic | 3125.0 | 25000.0 | 800000 | 800000 | 1.5 | 8 | 19 |
| Q6H9L7 | Cardiometabolic | 195.3 | 195.3 | 25000 | 200000 | 2.1 | 11 | 22 |

| | | | | | | | | |
|---|---|---|---|---|---|---|---|---|
| P07288 | Cardiometabolic | 195.3 | 195.3 | 25000 | 200000 | 2.1 | 5 | 5 |
| P16410 | Cardiometabolic | 97.7 | 97.7 | 25000 | 100000 | 2.4 | 1 | |
| P40199 | Cardiometabolic | 3125.0 | 3125.0 | 400000 | 800000 | 2.1 | 10 | 15 |
| Q8N6C8 | Cardiometabolic II | | | | | | 5 | 13 |
| Q02817 | Cardiometabolic | 6.1 | 12.2 | 6250 | 25000 | 2.7 | 7 | 30 |
| P98095 | Cardiometabolic | 195.3 | 195.3 | 25000 | 100000 | 2.1 | 4 | 9 |
| P02461 | Cardiometabolic | 97.7 | 195.3 | 25000 | 100000 | 2.1 | 6 | 10 |
| Q6UWP8 | Cardiometabolic II | | | | | | 7 | 19 |
| Q6UVK1 | Cardiometabolic | 97.7 | 97.7 | 25000 | 100000 | 2.4 | 5 | 6 |
| P39059 | Cardiometabolic | 195.3 | 195.3 | 12500 | 50000 | 1.8 | 5 | 10 |
| Q9BYJ0 | Cardiometabolic | 195.3 | 195.3 | 25000 | 50000 | 2.1 | 4 | 15 |
| Q9HCU0 | Cardiometabolic | 97.7 | 195.3 | 25000 | 100000 | 2.1 | 5 | 10 |
| Q96CG8 | Cardiometabolic | 781.3 | 1562.5 | 50000 | 200000 | 1.5 | 8 | 13 |
| Q96NZ9 | Cardiometabolic II | | | | | | 4 | 12 |
| P47972 | Cardiometabolic | 48.8 | 97.7 | 25000 | 100000 | 2.4 | 7 | 14 |
| P02818 | Cardiometabolic | 781.3 | 1562.5 | 100000 | 400000 | 1.8 | 6 | 16 |
| Q8N114 | Cardiometabolic | 390.6 | 390.6 | 25000 | 50000 | 1.8 | 6 | 9 |
| Q6IBS0 | Cardiometabolic | 781.3 | 781.3 | 200000 | 800000 | 2.4 | | |
| P30405 | Cardiometabolic | 6250.0 | 6250.0 | 200000 | 800000 | 1.5 | 10 | 28 |
| P32971 | Cardiometabolic | 12.2 | 24.4 | 12500 | 50000 | 2.7 | 11 | 11 |
| Q9Y2Y8 | Cardiometabolic | 97.7 | 195.3 | 25000 | 200000 | 2.1 | 5 | 14 |
| P35579 | Cardiometabolic II | | | | | | | |
| P13727 | Cardiometabolic | 97.7 | 97.7 | 25000 | 50000 | 2.4 | 5 | 15 |
| P08575 | Cardiometabolic | 48.8 | 48.8 | 12500 | 100000 | 2.4 | 7 | 7 |
| O43280 | Cardiometabolic | 48.8 | 97.7 | 25000 | 800000 | 2.4 | 5 | 20 |
| Q9NRR1 | Cardiometabolic | 390.6 | 390.6 | 25000 | 200000 | 1.8 | 6 | 12 |
| O75339 | Cardiometabolic II | | | | | | 9 | 26 |
| Q9H2X3 | Cardiometabolic | 24.4 | 48.8 | 6250 | 25000 | 2.1 | 6 | 14 |
| Q9Y646 | Cardiometabolic | 195.3 | 195.3 | 50000 | 200000 | 2.4 | 4 | 11 |
| P10645 | Cardiometabolic | 1562.5 | 3125.0 | 200000 | 800000 | 1.8 | 9 | 22 |
| Q04721 | Cardiometabolic | 48.8 | 97.7 | 6250 | 50000 | 1.8 | 5 | 6 |
| O95965 | Cardiometabolic | 97.7 | 195.3 | 25000 | 100000 | 2.1 | 6 | 13 |
| Q9Y251 | Cardiometabolic II | | | | | | 6 | 33 |

| | | | | | | | | |
|---|---|---|---|---|---|---|---|---|
| Q8TDY8 | Cardiometabolic | 48.8 | 48.8 | 6250 | 50000 | 2.1 | 6 | 8 |
| Q15063 | Cardiometabolic II | | | | | | 9 | 15 |
| P08217 | Cardiometabolic | 48.8 | 48.8 | 25000 | 100000 | 2.7 | 5 | 9 |
| Q9UQP3 | Cardiometabolic | 50000.0 | 100000.0 | 6400000 | 12800000 | 1.8 | 7 | 8 |
| P17900 | Cardiometabolic | 781.3 | 1562.5 | 50000 | 200000 | 1.5 | 7 | 17 |
| P37837 | Cardiometabolic | 97.7 | 195.3 | 25000 | 200000 | 2.1 | 4 | 8 |
| Q8WWQ8 | Cardiometabolic | 390.6 | 1562.5 | 100000 | 800000 | 1.8 | 5 | 9 |
| P55000 | Cardiometabolic | 1562.5 | 1562.5 | 100000 | 800000 | 1.8 | 7 | 9 |
| P12277 | Cardiometabolic II | | | | | | 9 | 15 |
| Q13510 | Cardiometabolic | 195.3 | 390.6 | 25000 | 200000 | 1.8 | 8 | 18 |
| P11279 | Cardiometabolic | 97.7 | 390.6 | 50000 | 100000 | 2.1 | 7 | 6 |
| P07602 | Cardiometabolic | 1562.5 | 1562.5 | 50000 | 200000 | 1.5 | 6 | 7 |
| P17174 | Cardiometabolic | 390.6 | 390.6 | 25000 | 50000 | 1.8 | 8 | 12 |
| P61916 | Cardiometabolic | 390.6 | 390.6 | 12500 | 50000 | 1.5 | 5 | 8 |
| P19878 | Inflammation | 0.8 | 3.1 | 12500 | 25000 | 3.6 | 8 | 6 |
| P40933 | Inflammation | 1.5 | 3.1 | 50000 | 400000 | 4.2 | 9 | 4 |
| P11274 | Inflammation | 781.3 | 1562.5 | 200000 | 800000 | 2.1 | 7 | 4 |
| P52564 | Inflammation | 24.4 | 48.8 | 25000 | 200000 | 2.7 | 9 | 10 |
| Q9UN19 | Inflammation | 3.1 | 12.2 | 25000 | 200000 | 3.3 | 10 | 9 |
| P24394 | Inflammation | 3.1 | 6.1 | 25000 | 100000 | 3.6 | 10 | 12 |
| Q6ZUJ8 | Inflammation | 781.3 | 1562.5 | 400000 | 800000 | 2.4 | 14 | 13 |
| P01730 | Inflammation | 97.7 | 97.7 | 400000 | 800000 | 3.6 | 8 | 9 |
| Q13241 | Inflammation | 1.5 | 3.1 | 6250 | 25000 | 3.3 | 3 | 4 |
| P35613 | Inflammation | 0.2 | 0.4 | 12500 | 50000 | 4.5 | 3 | 3 |
| P50452 | Inflammation | 12.2 | 12.2 | 12500 | 50000 | 3.0 | 6 | 6 |
| O43915 | Inflammation | 12.2 | 24.4 | 25000 | 100000 | 3.0 | 4 | 4 |
| O00253 | Inflammation | 48.8 | 97.7 | 50000 | 400000 | 2.7 | 6 | 14 |
| P10147 | Inflammation | 0.2 | 0.4 | 781 | 1563 | 3.3 | 9 | 5 |
| Q92609 | Inflammation | | | | | | 7 | 7 |
| Q9GZT9 | Inflammation | 6.1 | 12.2 | 12500 | 25000 | 3.0 | 9 | 5 |
| Q9Y266 | Inflammation | 24.4 | 97.7 | 100000 | 800000 | 3.0 | 4 | 5 |
| Q14242 | Inflammation | 3.1 | 6.1 | 6250 | 25000 | 3.0 | 9 | 5 |
| Q12918 | Inflammation | 0.8 | 1.5 | 3125 | 12500 | 3.3 | 3 | 2 |

| | | | | | | | | |
|---|---|---|---|---|---|---|---|---|
| Q3KPI0 | Inflammation | 0.4 | 0.8 | 1563 | 12500 | 3.3 | 8 | 6 |
| Q9NRM6 | Inflammation | 24.4 | 24.4 | 25000 | 400000 | 3.0 | 8 | 8 |
| Q01344 | Inflammation | 6.1 | 12.2 | 25000 | 400000 | 3.3 | 9 | 8 |
| P02745 | Inflammation | 29.2 | 58.3 | 119500 | 239000 | 3.3 | 4 | 5 |
| Q9HBG7 | Inflammation | 3.1 | 6.1 | 12500 | 50000 | 3.3 | 3 | 4 |
| O94992 | Inflammation | 1.5 | 3.1 | 781 | 1563 | 2.4 | 10 | 7 |
| Q08174 | Inflammation | 195.3 | 390.6 | 200000 | 800000 | 2.7 | 4 | 4 |
| O60449 | Inflammation | 24.4 | 48.8 | 50000 | 400000 | 3.0 | 10 | 7 |
| O15455 | Inflammation | 1.5 | 3.1 | 12500 | 100000 | 3.6 | 4 | 3 |
| P22304 | Inflammation | 97.7 | 195.3 | 400000 | 800000 | 3.3 | 3 | 4 |
| P43234 | Inflammation | 48.8 | 97.7 | 50000 | 400000 | 2.7 | 4 | 4 |
| P14210 | Inflammation | 3.1 | 12.2 | 100000 | 200000 | 3.9 | 4 | 3 |
| Q12866 | Inflammation | 97.7 | 195.3 | 100000 | 800000 | 2.7 | 5 | 6 |
| P51671 | Inflammation | 1.5 | 3.1 | 25000 | 200000 | 3.9 | 3 | 4 |
| P42701 | Inflammation | 97.7 | 195.3 | 100000 | 800000 | 2.7 | 8 | 17 |
| P09874 | Inflammation | 48.8 | 97.7 | 50000 | 200000 | 2.7 | 7 | 14 |
| Q5R372 | Inflammation | 48.8 | 48.8 | 25000 | 200000 | 2.7 | 10 | 19 |
| Q13459 | Inflammation | | | | | | 10 | 19 |
| O95760 | Inflammation | 6.1 | 24.4 | 6250 | 25000 | 2.4 | 8 | 58 |
| P14784 | Inflammation | 97.7 | 390.6 | 200000 | 800000 | 2.7 | 10 | 37 |
| Q8NHJ6 | Inflammation | 97.7 | 195.3 | 100000 | 800000 | 2.7 | 8 | 15 |
| P01584 | Inflammation | 0.4 | 1.5 | 25000 | 100000 | 4.2 | 10 | 20 |
| P60568 | Inflammation | 0.4 | 0.8 | 6250 | 25000 | 3.9 | 7 | 20 |
| O76038 | Inflammation | 97.7 | 390.6 | 200000 | 800000 | 2.7 | 9 | 11 |
| O95715 | Inflammation | | | | | | 8 | 15 |
| Q8N6P7 | Inflammation | 0.4 | 1.5 | 3125 | 12500 | 3.3 | 8 | 22 |
| P22301 | Inflammation | 6.1 | 24.4 | 200000 | 800000 | 3.9 | 10 | 18 |
| Q9UPV0 | Inflammation | 12.2 | 48.8 | 12500 | 25000 | 2.4 | 7 | 18 |
| P28838 | Inflammation | | | | | | 12 | 20 |
| O60934 | Inflammation | | | | | | 11 | 9 |
| P57771 | Inflammation | | | | | | 18 | 14 |
| Q03426 | Inflammation | 390.6 | 781.3 | 400000 | 800000 | 2.7 | 11 | 12 |
| O14904 | Inflammation | 3.1 | 6.1 | 25000 | 100000 | 3.6 | 9 | 11 |

| | | | | | | | | |
|---|---|---|---|---|---|---|---|---|
| Q9Y478 | Inflammation | 48.8 | 97.7 | 25000 | 100000 | 2.4 | 7 | 21 |
| P20809 | Inflammation | 6.1 | 12.2 | 6250 | 100000 | 2.7 | 9 | 9 |
| P05412 | Inflammation | 390.6 | 781.3 | 100000 | 3200000 | 2.1 | 11 | 14 |
| O43707 | Inflammation | 390.6 | 781.3 | 100000 | 400000 | 2.1 | 7 | 14 |
| Q96PD4 | Inflammation | 1.5 | 6.1 | 6250 | 25000 | 3.0 | 8 | 18 |
| P05112 | Inflammation | 0.8 | 1.5 | 6250 | 25000 | 3.6 | 7 | 33 |
| P35225 | Inflammation | 1.5 | 6.1 | 100000 | 800000 | 4.2 | 9 | 22 |
| Q96AX2 | Inflammation | 12.2 | 24.4 | 12500 | 100000 | 2.7 | 9 | 19 |
| Q9NYY1 | Inflammation | 24.4 | 97.7 | 25000 | 800000 | 2.4 | 7 | 16 |
| Q96P31 | Inflammation | 48.8 | 97.7 | 50000 | 800000 | 2.7 | 8 | 18 |
| Q9NP70 | Inflammation | 12.2 | 24.4 | 25000 | 400000 | 3.0 | 9 | 15 |
| Q13007 | Inflammation | 24.4 | 48.8 | 12500 | 25000 | 2.4 | 14 | 16 |
| Q9HCU5 | Inflammation | 390.6 | 781.3 | 200000 | 800000 | 2.4 | 9 | 12 |
| Q8WV07 | Inflammation | 12.2 | 24.4 | 50000 | 100000 | 3.3 | 8 | 17 |
| Q9Y2J8 | Inflammation | 48.8 | 97.7 | 400000 | 800000 | 3.6 | 8 | 15 |
| Q9Y3P8 | Inflammation | 0.04 | 4.5 | 18500 | 37000 | 3.6 | 8 | 16 |
| Q8IU57 | Inflammation | 0.8 | 1.5 | 25000 | 100000 | 4.2 | 11 | 5 |
| P30838 | Inflammation | 0.8 | 3.1 | 3125 | 25000 | 3.0 | 10 | 10 |
| O14867 | Inflammation | 12.2 | 24.4 | 25000 | 200000 | 3.0 | 8 | 7 |
| P19801 | Inflammation | 97.7 | 195.3 | 100000 | 800000 | 2.7 | 10 | 21 |
| Q16552 | Inflammation | 6.1 | 12.2 | 50000 | 100000 | 3.6 | 7 | 7 |
| Q7Z739 | Inflammation | 12.2 | 24.4 | 12500 | 100000 | 2.7 | 9 | 32 |
| O60575 | Inflammation | | | | | | 10 | 6 |
| P26951 | Inflammation | | | | | | 7 | 28 |
| Q8TAD2 | Inflammation | 3.1 | 6.1 | 6250 | 25000 | 3.0 | 8 | 14 |
| Q9P0M4 | Inflammation | 12.2 | 24.4 | 50000 | 100000 | 3.3 | 10 | 14 |
| Q7Z6M3 | Inflammation | | | | | | 7 | 21 |
| Q8TCS8 | Inflammation | | | | | | 9 | 11 |
| Q5T4W7 | Inflammation | 0.8 | 1.5 | 6250 | 25000 | 3.6 | 9 | 9 |
| Q99748 | Inflammation | 0.8 | 1.5 | 3125 | 25000 | 3.3 | 15 | 31 |
| P48061 | Inflammation | | | | | | 8 | 17 |
| Q04759 | Inflammation | | | | | | 13 | 18 |
| Q12933 | Inflammation | | | | | | 9 | 4 |

| | | | | | | | | |
|---|---|---|---|---|---|---|---|---|
| P42768 | Inflammation | | | | | | 8 | 31 |
| O95379 | Inflammation | | | | | | 11 | 16 |
| Q13219 | Inflammation | 195.3 | 390.6 | 50000 | 200000 | 2.1 | 12 | 15 |
| Q13574 | Inflammation | 3.1 | 6.1 | 3125 | 25000 | 2.7 | 8 | 13 |
| P63241 | Inflammation | | | | | | 11 | 10 |
| O43736 | Inflammation | | | | | | 7 | 9 |
| O60542 | Inflammation | 3.1 | 6.1 | 12500 | 100000 | 3.3 | 11 | 5 |
| P13693 | Inflammation | 1562.5 | 6250.0 | 400000 | 800000 | 1.8 | 16 | 19 |
| P09038 | Inflammation | 12.2 | 12.2 | 12500 | 400000 | 3.0 | 4 | 6 |
| Q9Y5A7 | Inflammation | 3.1 | 6.1 | 12500 | 25000 | 3.3 | 8 | 12 |
| Q6UXK5 | Inflammation | 12.2 | 24.4 | 25000 | 400000 | 3.0 | 6 | 17 |
| P01375 | Inflammation | 1.5 | 3.1 | 6250 | 100000 | 3.3 | 9 | 15 |
| Q13651 | Inflammation | 1.5 | 6.1 | 400000 | 800000 | 4.8 | 7 | 17 |
| Q96RJ3 | Inflammation | 12.2 | 24.4 | 6250 | 100000 | 2.4 | 9 | 13 |
| P27540 | Inflammation | | | | | | 8 | 14 |
| Q969V3 | Inflammation | | | | | | 6 | 17 |
| Q9UHF4 | Inflammation | 0.8 | 1.5 | 12500 | 400000 | 3.9 | 8 | 13 |
| Q06520 | Inflammation | 195.3 | 390.6 | 200000 | 400000 | 2.7 | 10 | 8 |
| Q6UB28 | Inflammation | 0.4 | 0.8 | 12500 | 25000 | 4.2 | 8 | 9 |
| Q0Z7S8 | Inflammation | 3.1 | 12.2 | 12500 | 25000 | 3.0 | 10 | 7 |
| O60880 | Inflammation | 6.1 | 12.2 | 6250 | 25000 | 2.7 | 8 | 5 |
| Q12968 | Inflammation | 6.1 | 24.4 | 3125 | 12500 | 2.1 | 8 | 9 |
| P78362 | Inflammation | 3.1 | 6.1 | 1563 | 12500 | 2.4 | 5 | 11 |
| P01903 | Inflammation | 390.6 | 781.3 | 400000 | 800000 | 2.7 | 10 | 9 |
| P78410 | Inflammation | 48.8 | 97.7 | 200000 | 800000 | 3.3 | 10 | 7 |
| O43521-2 | Inflammation | | | | | | 9 | 8 |
| P01583 | Inflammation | 1.5 | 3.1 | 3148 | 12594 | 3.0 | 8 | 47 |
| P01579 | Inflammation | 1.5 | 6.1 | 12500 | 50000 | 3.3 | 15 | 22 |
| Q05084 | Inflammation | 48.8 | 195.3 | 100000 | 400000 | 2.7 | 8 | 16 |
| Q7L8A9 | Inflammation | 195.3 | 390.6 | 100000 | 400000 | 2.4 | 6 | 9 |
| P05113 | Inflammation | 6.1 | 12.2 | 100000 | 400000 | 3.9 | 10 | 21 |
| O43597 | Inflammation | 24.4 | 97.7 | 50000 | 400000 | 2.7 | 9 | 15 |
| Q13261 | Inflammation | 6.1 | 12.2 | 3125 | 25000 | 2.4 | 7 | 23 |

| | | | | | | | | |
|---|---|---|---|---|---|---|---|---|
| P12034 | Inflammation | 3.1 | 6.1 | 50000 | 100000 | 3.9 | 11 | 19 |
| Q92844 | Inflammation | 48.8 | 195.3 | 100000 | 400000 | 2.7 | 8 | 15 |
| O95644 | Inflammation | 390.6 | 781.3 | 400000 | 800000 | 2.7 | 10 | 9 |
| P09919 | Inflammation | 97.7 | 390.6 | 400000 | 800000 | 3.0 | 9 | 14 |
| Q9BXJ7 | Inflammation | | | | | | 8 | 19 |
| Q13291 | Inflammation | 781.3 | 1562.5 | 400000 | 800000 | 2.4 | 14 | 17 |
| P51617 | Inflammation | 781.3 | 3125.0 | 400000 | 800000 | 2.1 | 11 | 14 |
| Q12778 | Inflammation | | | | | | 8 | 8 |
| Q14435 | Inflammation | 781.3 | 1562.5 | 400000 | 800000 | 2.4 | 11 | 14 |
| P30048 | Inflammation | | | | | | 8 | 12 |
| P32456 | Inflammation | 390.6 | 781.3 | 400000 | 800000 | 2.7 | 12 | 20 |
| P01591 | Inflammation | | | | | | 13 | 19 |
| P55957 | Inflammation | 781.3 | 3125.0 | 400000 | 3200000 | 2.1 | 11 | 11 |
| Q12765 | Inflammation | 48.8 | 97.7 | 50000 | 400000 | 2.7 | 9 | 12 |
| Q6ZMH5 | Inflammation | | | | | | 10 | 9 |
| Q8N8S7 | Inflammation | 48.8 | 48.8 | 6250 | 25000 | 2.1 | 10 | 20 |
| Q9Y6K9 | Inflammation | 3.1 | 12.2 | 12500 | 25000 | 3.0 | 13 | 14 |
| P18564 | Inflammation | 1.5 | 3.1 | 6250 | 25000 | 3.3 | 8 | 14 |
| P58294 | Inflammation | 3.1 | 12.2 | 12500 | 25000 | 3.0 | 9 | 5 |
| Q9HB29 | Inflammation | 0.8 | 1.5 | 6250 | 100000 | 3.6 | 8 | 16 |
| P05231 | Inflammation | 0.2 | 0.4 | 3125 | 25000 | 3.9 | 10 | 14 |
| P12872 | Inflammation | 390.6 | 781.3 | 200000 | 400000 | 2.4 | 12 | 10 |
| Q96DB9 | Inflammation | 6.1 | 12.2 | 3125 | 100000 | 2.4 | 8 | 13 |
| Q96LC7 | Inflammation | 12.2 | 48.8 | 200000 | 800000 | 3.6 | 8 | 4 |
| O75475 | Inflammation | | | | | | 9 | 8 |
| P19474 | Inflammation | 24.4 | 97.7 | 400000 | 800000 | 3.6 | 12 | 9 |
| B1AKI9 | Inflammation | 97.7 | 195.3 | 200000 | 400000 | 3.0 | 10 | 10 |
| P13232 | Inflammation | 1.5 | 3.1 | 6250 | 12500 | 3.3 | 12 | 9 |
| P13747 | Inflammation | 12500.0 | 25000.0 | 1600000 | 3200000 | 1.8 | 6 | 4 |
| Q9UNK0 | Inflammation | 12.2 | 48.8 | 12500 | 25000 | 2.4 | 4 | 13 |
| P33241 | Inflammation | 97.7 | 97.7 | 25000 | 400000 | 2.4 | 10 | 8 |
| Q8WTT0 | Inflammation | 24.4 | 48.8 | 25000 | 400000 | 2.7 | 8 | 4 |
| P13725 | Inflammation | 0.2 | 0.4 | 781 | 3125 | 3.3 | 12 | 6 |

| | | | | | | | | |
|---|---|---|---|---|---|---|---|---|
| Q8IVG5 | Inflammation | 195.3 | 390.6 | 200000 | 800000 | 2.7 | 7 | 8 |
| Q8TD46 | Inflammation | 6.1 | 12.2 | 25000 | 100000 | 3.3 | 7 | 7 |
| Q9UHC6 | Inflammation | 12.2 | 24.4 | 12500 | 400000 | 2.7 | 10 | 10 |
| P50995 | Inflammation | 1562.5 | 3125.0 | 800000 | 800000 | 2.4 | 7 | 9 |
| Q6DN72 | Inflammation | 97.7 | 195.3 | 200000 | 800000 | 3.0 | 10 | 5 |
| P23582 | Inflammation | 12.2 | 24.4 | 12500 | 25000 | 2.7 | 11 | 8 |
| Q8NDB2 | Inflammation | 195.3 | 390.6 | 200000 | 400000 | 2.7 | 9 | 16 |
| Q01151 | Inflammation | 1.5 | 3.1 | 12500 | 25000 | 3.6 | 9 | 6 |
| P45984 | Inflammation | | | | | | 12 | 13 |
| Q9NRJ3 | Inflammation | 97.7 | 195.3 | 100000 | 400000 | 2.7 | 11 | 12 |
| Q9NZN5 | Inflammation | 781.3 | 3125.0 | 400000 | 800000 | 2.1 | 8 | 9 |
| Q9HD26 | Inflammation | 12.2 | 24.4 | 12500 | 25000 | 2.7 | 7 | 16 |
| P28827 | Inflammation | 97.7 | 195.3 | 200000 | 800000 | 3.0 | 11 | 11 |
| P29965 | Inflammation | 0.8 | 1.5 | 6250 | 12500 | 3.6 | 4 | 4 |
| P16455 | Inflammation | 195.3 | 390.6 | 100000 | 400000 | 2.4 | 12 | 13 |
| Q9BT73 | Inflammation | 390.6 | 781.3 | 200000 | 400000 | 2.4 | 9 | 10 |
| Q8N608 | Inflammation | 6.1 | 12.2 | 25000 | 200000 | 3.3 | 9 | 9 |
| P28845 | Inflammation | | | | | | 9 | 7 |
| Q9UNE0 | Inflammation | 0.4 | 0.8 | 781 | 12500 | 3.0 | 9 | 5 |
| P20849 | Inflammation | 6.1 | 12.2 | 12500 | 25000 | 3.0 | 11 | 6 |
| Q9HCM2 | Inflammation | 24.4 | 97.7 | 50000 | 400000 | 2.7 | 12 | 6 |
| P01588 | Inflammation | 3.9 | 7.8 | 1000 | 4000 | 2.1 | 11 | 11 |
| P23229 | Inflammation | | | | | | 9 | 7 |
| P80098 | Inflammation | 0.4 | 0.8 | 781 | 25000 | 3.0 | 10 | 8 |
| O76036 | Inflammation | 3.1 | 6.1 | 12500 | 25000 | 3.3 | 9 | 6 |
| P01374 | Inflammation | 0.8 | 1.5 | 3125 | 12500 | 3.3 | 8 | 4 |
| P42575 | Inflammation | 195.3 | 390.6 | 200000 | 800000 | 2.7 | 7 | 5 |
| P24071 | Inflammation | 0.8 | 3.1 | 12500 | 25000 | 3.6 | 9 | 9 |
| Q9NWZ3 | Inflammation | 390.6 | 1562.5 | 400000 | 800000 | 2.4 | 9 | 14 |
| Q6UXB4 | Inflammation | 3.1 | 6.1 | 50000 | 200000 | 3.9 | 10 | 6 |
| P37235 | Inflammation | 48.8 | 97.7 | 50000 | 200000 | 2.7 | 4 | 5 |
| Q9Y258 | Inflammation | 97.7 | 781.3 | 400000 | 800000 | 2.7 | 15 | 17 |
| Q9UKX5 | Inflammation | 97.7 | 390.6 | 200000 | 800000 | 2.7 | 10 | 15 |

| | | | | | | | | |
|---|---|---|---|---|---|---|---|---|
| Q9H0P0 | Inflammation | 97.7 | 195.3 | 200000 | 800000 | 3.0 | 9 | 7 |
| P08727 | Inflammation | 48.8 | 97.7 | 100000 | 400000 | 3.0 | 8 | 9 |
| P20340 | Inflammation | 48.8 | 97.7 | 200000 | 800000 | 3.3 | 8 | 8 |
| Q9UIB8 | Inflammation | 24.4 | 48.8 | 12500 | 200000 | 2.4 | 9 | 7 |
| P78310 | Inflammation | 3.1 | 3.1 | 1563 | 12500 | 2.7 | 12 | 7 |
| P32970 | Inflammation | 12.2 | 48.8 | 100000 | 200000 | 3.3 | 10 | 6 |
| Q29983_Q2 | Inflammation | 6.1 | 12.2 | 12500 | 25000 | 3.0 | 10 | 8 |
| O14788 | Inflammation | 12.2 | 24.4 | 50000 | 200000 | 3.3 | 11 | 7 |
| Q9UDT6 | Inflammation | 195.3 | 195.3 | 50000 | 800000 | 2.4 | 13 | 20 |
| Q9C035 | Inflammation | | | | | | 10 | 17 |
| P26022 | Inflammation | | | | | | 9 | 7 |
| Q07065 | Inflammation | | | | | | 3 | 6 |
| P80162 | Inflammation | | | | | | 6 | 13 |
| P20783 | Inflammation | 0.4 | 0.8 | 3125 | 12500 | 3.6 | 15 | 7 |
| Q14773 | Inflammation | 781.3 | 1562.5 | 400000 | 800000 | 2.4 | 9 | 5 |
| Q16698 | Inflammation | | | | | | 11 | 11 |
| P50591 | Inflammation | 24.4 | 97.7 | 25000 | 50000 | 2.4 | 4 | 4 |
| Q8WXI8 | Inflammation | 3.1 | 6.1 | 12500 | 25000 | 3.3 | 10 | 8 |
| O94856 | Inflammation | | | | | | 3 | 2 |
| P49771 | Inflammation | 0.4 | 0.8 | 1563 | 12500 | 3.3 | 6 | 4 |
| Q14005 | Inflammation | 1.5 | 3.1 | 3125 | 12500 | 3.0 | 3 | 6 |
| Q15517 | Inflammation | 390.6 | 781.3 | 400000 | 800000 | 2.7 | 4 | 4 |
| O15169 | Inflammation | 781.3 | 1562.5 | 400000 | 800000 | 2.4 | 6 | 9 |
| Q9NQ25 | Inflammation | 195.3 | 390.6 | 200000 | 800000 | 2.7 | 10 | 6 |
| Q9UMR7 | Inflammation | 0.8 | 3.1 | 6250 | 25000 | 3.3 | 9 | 7 |
| O43561 | Inflammation | 48.8 | 97.7 | 25000 | 100000 | 2.4 | 7 | 8 |
| P10145 | Inflammation | 0.1 | 0.2 | 1563 | 6250 | 3.9 | 3 | 4 |
| Q96SB3 | Inflammation | 12.2 | 24.4 | 25000 | 800000 | 3.0 | 7 | 12 |
| P41217 | Inflammation | 6.1 | 12.2 | 3125 | 25000 | 2.4 | 10 | 6 |
| P14317 | Inflammation | 195.3 | 195.3 | 25000 | 400000 | 2.1 | 8 | 8 |
| Q9BZW8 | Inflammation | 1.5 | 3.1 | 6250 | 25000 | 3.3 | 4 | 5 |
| Q16719 | Inflammation | 48.8 | 48.8 | 400000 | 800000 | 3.9 | 3 | 11 |
| O00273 | Inflammation | 0.8 | 3.1 | 6250 | 50000 | 3.3 | 5 | 6 |

| | | | | | | | | |
|---|---|---|---|---|---|---|---|---|
| Q13478 | Inflammation | 0.2 | 0.8 | 6250 | 25000 | 3.9 | 3 | 3 |
| O75077 | Inflammation | 12.2 | 24.4 | 50000 | 200000 | 3.3 | 5 | 3 |
| Q9UQV4 | Inflammation | 12.2 | 24.4 | 25000 | 200000 | 3.0 | 5 | 4 |
| P24001 | Inflammation | 0.8 | 3.1 | 12500 | 25000 | 3.6 | 9 | 7 |
| P36959 | Inflammation | | | | | | 4 | 4 |
| P30203 | Inflammation | 1.5 | 1.5 | 6250 | 25000 | 3.6 | 4 | 4 |
| P20273 | Inflammation | 3.1 | 6.1 | 50000 | 200000 | 3.9 | 3 | 3 |
| Q6UXB2 | Inflammation | 3.1 | 6.1 | 12500 | 50000 | 3.3 | 4 | 4 |
| P68106 | Inflammation | 390.6 | 781.3 | 200000 | 400000 | 2.4 | 5 | 26 |
| P12544 | Inflammation | 390.6 | 390.6 | 800000 | 800000 | 3.3 | 5 | 5 |
| O95971 | Inflammation | 1.5 | 3.1 | 12500 | 25000 | 3.6 | 3 | 7 |
| P43489 | Inflammation | 0.8 | 1.5 | 6250 | 12500 | 3.6 | 5 | 4 |
| P01137 | Inflammation | | | | | | 4 | 4 |
| Q15661 | Inflammation | 6.1 | 12.2 | 100000 | 200000 | 3.9 | 4 | 3 |
| Q04637 | Inflammation | 12.2 | 24.4 | 25000 | 200000 | 3.0 | 6 | 10 |
| P48023 | Inflammation | 0.2 | 0.2 | 6250 | 12500 | 4.5 | 4 | 4 |
| P40259 | Inflammation | 0.8 | 1.5 | 6250 | 25000 | 3.6 | 4 | 4 |
| Q03431 | Inflammation | 3.1 | 3.1 | 6250 | 25000 | 3.3 | 7 | 8 |
| Q9Y6Q6 | Inflammation | 0.8 | 1.5 | 6250 | 12500 | 3.6 | 4 | 4 |
| Q96LA5 | Inflammation | 24.4 | 48.8 | 50000 | 200000 | 3.0 | 5 | 3 |
| Q9BXN2 | Inflammation | 3.1 | 6.1 | 3125 | 12500 | 2.7 | 3 | 5 |
| Q9H4D0 | Inflammation | 6.1 | 6.1 | 25000 | 200000 | 3.6 | 5 | 3 |
| P29460 | Inflammation | 0.4 | 0.8 | 3125 | 12500 | 3.6 | 5 | 4 |
| P42702 | Inflammation | 6.1 | 48.8 | 12500 | 400000 | 2.4 | 12 | 7 |
| Q99616 | Inflammation | 0.8 | 6.1 | 3125 | 6250 | 2.7 | 5 | 5 |
| P00813 | Inflammation | 781.3 | 1562.5 | 3200000 | 12800000 | 3.3 | 4 | 3 |
| P30044 | Inflammation | 1562.5 | 3125.0 | 800000 | 800000 | 2.4 | 4 | 25 |
| O60884 | Inflammation | 97.7 | 390.6 | 400000 | 800000 | 3.0 | 6 | 16 |
| P15692 | Inflammation | 0.8 | 1.5 | 6250 | 12500 | 3.6 | 7 | 7 |
| O43508 | Inflammation | 12.2 | 24.4 | 100000 | 400000 | 3.6 | 5 | 4 |
| O15444 | Inflammation | 3.1 | 24.4 | 25000 | 100000 | 3.0 | 5 | 6 |
| P10144 | Inflammation | | | | | | 12 | 7 |
| P01135 | Inflammation | 0.2 | 0.4 | 1563 | 25000 | 3.6 | 12 | 7 |

| | | | | | | | | |
|---|---|---|---|---|---|---|---|---|
| P78556 | Inflammation | 3.1 | 6.1 | 6250 | 25000 | 3.0 | 4 | 5 |
| P03956 | Inflammation | 0.8 | 1.5 | 3125 | 12500 | 3.3 | 4 | 5 |
| P49763 | Inflammation | 0.4 | 0.8 | 6250 | 12500 | 3.9 | 4 | 3 |
| Q9BY76 | Inflammation | 12.2 | 24.4 | 25000 | 50000 | 3.0 | 7 | 4 |
| O95750 | Inflammation | 3.1 | 6.1 | 12500 | 50000 | 3.3 | 6 | 4 |
| O14836 | Inflammation | 6.1 | 24.4 | 50000 | 200000 | 3.3 | 8 | 4 |
| P46109 | Inflammation | 6.1 | 12.2 | 25000 | 100000 | 3.3 | 6 | 8 |
| Q03405 | Inflammation | 0.2 | 0.4 | 1563 | 6250 | 3.6 | 6 | 4 |
| O43598 | Inflammation | 6.1 | 12.2 | 6250 | 12500 | 2.7 | 7 | 8 |
| Q9HCB6 | Inflammation | 48.8 | 97.7 | 100000 | 200000 | 3.0 | 5 | 4 |
| Q9NQ30 | Inflammation | 3.1 | 6.1 | 6250 | 25000 | 3.0 | 8 | 5 |
| Q16651 | Inflammation | 0.4 | 0.8 | 6250 | 12500 | 3.9 | 8 | 3 |
| O00468 | Inflammation | 6.1 | 12.2 | 6250 | 12500 | 2.7 | 7 | 5 |
| P29350 | Inflammation | 781.3 | 1562.5 | 100000 | 200000 | 1.8 | 6 | 7 |
| Q07325 | Inflammation | 0.05 | 0.1 | 6250 | 12500 | 4.8 | 7 | 5 |
| Q9UII2 | Inflammation | 195.3 | 390.6 | 50000 | 200000 | 2.1 | 8 | 7 |
| Q9H008 | Inflammation | 0.8 | 1.5 | 12500 | 25000 | 3.9 | 8 | 5 |
| P19876 | Inflammation | 12.2 | 48.8 | 50000 | 200000 | 3.0 | 10 | 9 |
| Q6UWV6 | Inflammation | 0.8 | 1.5 | 12500 | 200000 | 3.9 | 6 | 4 |
| P09341 | Inflammation | 0.4 | 0.8 | 6250 | 12500 | 3.9 | 8 | 8 |
| Q9H3U7 | Inflammation | 24.4 | 48.8 | 50000 | 200000 | 3.0 | 7 | 3 |
| Q92583 | Inflammation | 0.4 | 0.8 | 781 | 3125 | 3.0 | 7 | 14 |
| Q99538 | Inflammation | 0.8 | 3.1 | 6250 | 50000 | 3.3 | 9 | 5 |
| O00182 | Inflammation | 48.8 | 97.7 | 50000 | 200000 | 2.7 | 7 | 4 |
| Q03403 | Inflammation | 0.8 | 1.5 | 3125 | 12500 | 3.3 | 8 | 6 |
| P53634 | Inflammation | 24.4 | 48.8 | 50000 | 200000 | 3.0 | 8 | 12 |
| Q5ZPR3 | Inflammation | 97.7 | 195.3 | 400000 | 800000 | 3.3 | 7 | 4 |
| P55773 | Inflammation | 6.1 | 12.2 | 6250 | 25000 | 2.7 | 6 | 8 |
| P25116 | Inflammation | 48.8 | 195.3 | 200000 | 400000 | 3.0 | 8 | 5 |
| Q9NZC2 | Inflammation | | | | | | 6 | 10 |
| P34896 | Inflammation | 12.2 | 24.4 | 100000 | 400000 | 3.6 | 8 | 5 |
| Q15389 | Inflammation | 12.2 | 24.4 | 100000 | 200000 | 3.6 | 6 | 3 |
| O00626 | Inflammation | 6.1 | 6.1 | 3125 | 12500 | 2.7 | 14 | 20 |

| | | | | | | | | |
|---|---|---|---|---|---|---|---|---|
| O75888 | Inflammation | 3.1 | 6.1 | 25000 | 100000 | 3.6 | 5 | 5 |
| P47712 | Inflammation | 781.3 | 1562.5 | 400000 | 800000 | 2.4 | 6 | 7 |
| Q15166 | Inflammation | 390.6 | 390.6 | 200000 | 800000 | 2.7 | 5 | 9 |
| Q14118 | Inflammation | 3.1 | 6.1 | 6250 | 12500 | 3.0 | 7 | 4 |
| Q9BZZ2 | Inflammation | 12.2 | 24.4 | 100000 | 200000 | 3.6 | 7 | 4 |
| Q8NFT8 | Inflammation | 1.5 | 3.1 | 25000 | 100000 | 3.9 | 5 | 4 |
| Q99685 | Inflammation | 1562.5 | 12500.0 | 3200000 | 12800000 | 2.4 | 9 | 9 |
| O00585 | Inflammation | 24.4 | 48.8 | 200000 | 800000 | 3.6 | 8 | 4 |
| P19256 | Inflammation | 0.4 | 0.8 | 3125 | 12500 | 3.6 | 6 | 4 |
| P09326 | Inflammation | 6.1 | 12.2 | 100000 | 200000 | 3.9 | 6 | 4 |
| Q5KU26 | Inflammation | 1.5 | 3.1 | 6250 | 25000 | 3.3 | 7 | 6 |
| Q6GTX8 | Inflammation | 0.1 | 0.2 | 391 | 3125 | 3.3 | 7 | 4 |
| P09603 | Inflammation | 0.05 | 0.2 | 3125 | 6250 | 4.2 | 4 | 3 |
| P51888 | Inflammation | 6.1 | 12.2 | 12500 | 200000 | 3.0 | 6 | 4 |
| P16422 | Inflammation | 0.4 | 0.4 | 6250 | 25000 | 4.2 | 7 | 7 |
| P01133 | Inflammation | 0.1 | 0.2 | 781 | 1563 | 3.6 | 13 | 9 |
| P02778 | Inflammation | 0.2 | 6.1 | 6250 | 25000 | 3.0 | 7 | 9 |
| Q92484 | Inflammation | 6.1 | 12.2 | 100000 | 200000 | 3.9 | 6 | 4 |
| Q7KYR7 | Inflammation | 3.1 | 6.1 | 6250 | 25000 | 3.0 | 7 | 4 |
| O43291 | Inflammation | 6.1 | 12.2 | 25000 | 50000 | 3.3 | 8 | 6 |
| Q9Y6N7 | Inflammation | 3.1 | 6.1 | 100000 | 200000 | 4.2 | 6 | 4 |
| Q8WU39 | Inflammation | 0.4 | 0.8 | 6250 | 50000 | 3.9 | 7 | 2 |
| P35625 | Inflammation | 390.6 | 781.3 | 200000 | 400000 | 2.4 | 13 | 13 |
| O43639 | Inflammation | | | | | | 5 | 10 |
| O76096 | Inflammation | 0.4 | 0.8 | 6250 | 12500 | 3.9 | 7 | 5 |
| O00339 | Inflammation | 24.4 | 48.8 | 25000 | 50000 | 2.7 | 8 | 3 |
| O75462 | Inflammation | 390.6 | 781.3 | 100000 | 800000 | 2.1 | 6 | 7 |
| Q9UJU6 | Inflammation | 24.4 | 97.7 | 50000 | 200000 | 2.7 | 10 | 10 |
| Q15109 | Inflammation | 0.8 | 3.1 | 6250 | 25000 | 3.3 | 7 | 5 |
| Q08334 | Inflammation | 1.5 | 6.1 | 3125 | 12500 | 2.7 | 8 | 3 |
| Q9HC38 | Inflammation | 0.8 | 3.1 | 6250 | 25000 | 3.3 | 7 | 4 |
| P21709 | Inflammation | 3.1 | 6.1 | 50000 | 200000 | 3.9 | 7 | 3 |
| P27930 | Inflammation | 0.8 | 1.5 | 6250 | 12500 | 3.6 | 8 | 3 |

| | | | | | | | | |
|---|---|---|---|---|---|---|---|---|
| Q9UJA9 | Inflammation | 6.1 | 6.1 | 3125 | 12500 | 2.7 | 4 | 3 |
| Q9NZV1 | Inflammation | 1.5 | 3.1 | 6250 | 50000 | 3.3 | 8 | 4 |
| P15260 | Inflammation | 1.5 | 3.1 | 3125 | 12500 | 3.0 | 7 | 4 |
| P21860 | Inflammation | 6.1 | 12.2 | 12500 | 50000 | 3.0 | 7 | 5 |
| P09238 | Inflammation | 0.2 | 0.4 | 6250 | 12500 | 4.2 | 7 | 7 |
| Q9NR12 | Inflammation | 24.4 | 48.8 | 25000 | 50000 | 2.7 | 7 | 11 |
| P11684 | Inflammation | 390.6 | 781.3 | 400000 | 800000 | 2.7 | 6 | 4 |
| P18510 | Inflammation | 0.2 | 0.4 | 1563 | 12500 | 3.6 | 6 | 3 |
| Q14210 | Inflammation | 6.1 | 12.2 | 12500 | 200000 | 3.0 | 9 | 5 |
| Q14116 | Inflammation | 0.1 | 0.2 | 6250 | 12500 | 4.5 | 5 | 4 |
| P13236 | Inflammation | 0.2 | 0.4 | 1563 | 200000 | 3.6 | 7 | 5 |
| Q99435 | Inflammation | 12.2 | 24.4 | 50000 | 100000 | 3.3 | 6 | 2 |
| P36941 | Inflammation | 0.4 | 0.8 | 6250 | 12500 | 3.9 | 7 | 5 |
| P30613 | Inflammation | 3.1 | 6.1 | 50000 | 200000 | 3.9 | 6 | 7 |
| P55145 | Inflammation | 3.1 | 6.1 | 12500 | 25000 | 3.3 | 7 | 14 |
| O00241 | Inflammation | 1.5 | 1.5 | 6250 | 25000 | 3.6 | 4 | 4 |
| P29279 | Inflammation | 24.4 | 48.8 | 25000 | 200000 | 2.7 | 4 | 5 |
| P0DMV8 | Inflammation | 48.8 | 97.7 | 50000 | 200000 | 2.7 | 6 | 4 |
| O00300 | Inflammation | 0.1 | 0.2 | 6250 | 25000 | 4.5 | 7 | 3 |
| Q8WXD2 | Inflammation | 12.2 | 24.4 | 25000 | 200000 | 3.0 | 7 | 4 |
| O14773 | Inflammation | 1.5 | 3.1 | 12500 | 100000 | 3.6 | 6 | 3 |
| Q96KG7 | Inflammation | 87.9 | 175.8 | 90000 | 360000 | 2.7 | 5 | 5 |
| Q4KMG0 | Inflammation | 12.2 | 24.4 | 50000 | 200000 | 3.3 | 6 | 4 |
| O95866 | Inflammation | 0.8 | 1.5 | 1563 | 3125 | 3.0 | 12 | 14 |
| P56470 | Inflammation | 1.5 | 6.1 | 6250 | 25000 | 3.0 | 5 | 4 |
| O75563 | Inflammation | 24.4 | 48.8 | 50000 | 200000 | 3.0 | 9 | 11 |
| P01127 | Inflammation | 6.1 | 6.1 | 6250 | 12500 | 3.0 | 10 | 11 |
| Q96PL1 | Inflammation | 3.1 | 12.2 | 50000 | 200000 | 3.6 | 8 | 11 |
| O95633 | Inflammation | 12.2 | 12.2 | 6250 | 25000 | 2.7 | 6 | 5 |
| Q9UKU9 | Inflammation | 781.3 | 1562.5 | 6400000 | 12800000 | 3.6 | 9 | 8 |
| Q9BYZ8 | Inflammation | 48.8 | 97.7 | 200000 | 400000 | 3.3 | 7 | 9 |
| P24387 | Inflammation | 24.4 | 97.7 | 100000 | 800000 | 3.0 | 7 | 5 |
| Q99983 | Inflammation | 195.3 | 390.6 | 50000 | 200000 | 2.1 | 7 | 3 |

| | | | | | | | | |
|---|---|---|---|---|---|---|---|---|
| Q13232 | Inflammation | 6.1 | 12.2 | 6250 | 25000 | 2.7 | 8 | 5 |
| Q9Y3D6 | Inflammation | 195.3 | 781.3 | 800000 | 800000 | 3.0 | 9 | 12 |
| P19883 | Inflammation | 12.2 | 24.4 | 25000 | 50000 | 3.0 | 5 | 6 |
| P15291 | Inflammation | 24.4 | 48.8 | 100000 | 200000 | 3.3 | 10 | 6 |
| P12532 | Inflammation | 48.8 | 97.7 | 400000 | 800000 | 3.6 | 7 | 15 |
| Q9UHX3 | Inflammation | 0.8 | 1.5 | 6250 | 25000 | 3.6 | 7 | 5 |
| Q9NQ76 | Inflammation | 24.4 | 48.8 | 12500 | 25000 | 2.4 | 8 | 3 |
| Q6UXH1 | Inflammation | 3.1 | 6.1 | 12500 | 200000 | 3.3 | 8 | 3 |
| Q99895 | Inflammation | 3.1 | 6.1 | 12500 | 50000 | 3.3 | 7 | 9 |
| P07148 | Inflammation | 3.1 | 6.1 | 100000 | 200000 | 4.2 | 8 | 5 |
| Q16363 | Inflammation | 12.2 | 24.4 | 100000 | 200000 | 3.6 | 7 | 5 |
| Q92956 | Inflammation | 0.8 | 1.5 | 6250 | 25000 | 3.6 | 8 | 4 |
| P22466 | Inflammation | 24.4 | 48.8 | 100000 | 200000 | 3.3 | 8 | 7 |
| Q8TEU8 | Inflammation | 3.1 | 6.1 | 6250 | 25000 | 3.0 | 5 | 4 |
| Q8IYS5 | Inflammation | 3.1 | 6.1 | 6250 | 200000 | 3.0 | 7 | 5 |
| O00175 | Inflammation | 0.8 | 0.8 | 3125 | 6250 | 3.6 | 8 | 10 |
| P25942 | Inflammation | 0.1 | 0.2 | 3125 | 12500 | 4.2 | 9 | 8 |
| P54317 | Inflammation | 3.1 | 6.1 | 25000 | 200000 | 3.6 | 6 | 4 |
| Q9BU40 | Inflammation | 390.6 | 390.6 | 100000 | 200000 | 2.4 | 8 | 6 |
| Q8N907 | Inflammation II | | | | | | 10 | |
| Q04609 | Inflammation II | 6250.0 | 6250.0 | 400000 | 800000 | 1.8 | 12 | 14 |
| P62834 | Inflammation II | | | | | | 11 | 16 |
| P10070 | Inflammation II | 195.3 | 195.3 | 12500 | 25000 | 1.8 | | |
| P61328 | Inflammation II | 24.4 | 48.8 | 3125 | 25000 | 1.8 | | |
| O00206 | Inflammation II | | | | | | | |
| P46013 | Inflammation II | 48.8 | 97.7 | 12500 | 100000 | 2.1 | 13 | 24 |
| P24864 | Inflammation II | 390.6 | 781.3 | 50000 | 400000 | 1.8 | | |
| Q9H832 | Inflammation II | 390.6 | 781.3 | 100000 | 800000 | 2.1 | 13 | 21 |
| P85299 | Inflammation II | 3125.0 | 6250.0 | 400000 | 800000 | 1.8 | 13 | 0.3 |
| P49715 | Inflammation II | | | | | | | |
| Q9Y4C1 | Inflammation II | 1562.5 | 3125.0 | 200000 | 800000 | 1.8 | | |
| Q9NP95 | Inflammation II | 3125.0 | 6250.0 | 800000 | 800000 | 2.1 | 6 | 18 |
| P06401 | Inflammation II | 781.3 | 781.3 | 200000 | 800000 | 2.4 | 10 | 9 |

| | | | | | | | | |
|---|---|---|---|---|---|---|---|---|
| Q6UXL0 | Inflammation II | 390.6 | 781.3 | 200000 | 800000 | 2.4 | | |
| Q9NP85 | Inflammation II | 390.6 | 781.3 | 50000 | 800000 | 1.8 | 6 | 20 |
| P15531 | Inflammation II | 3125.0 | 3125.0 | 200000 | 400000 | 1.8 | | |
| Q6UXM1 | Inflammation II | | | | | | | |
| Q05329 | Inflammation II | 1562.5 | 6250.0 | 400000 | 800000 | 1.8 | | |
| P09693 | Inflammation II | 390.6 | 781.3 | 50000 | 400000 | 1.8 | 10 | 13 |
| P24928 | Inflammation II | | | | | | 6 | |
| Q09472 | Inflammation II | 390.6 | 390.6 | 12500 | 25000 | 1.5 | 3 | |
| Q9HBE5 | Inflammation II | 781.3 | 781.3 | 50000 | 400000 | 1.8 | | |
| P43378 | Inflammation II | 1562.5 | 6250.0 | 400000 | 800000 | 1.8 | | |
| Q92185 | Inflammation II | 390.6 | 1562.5 | 200000 | 800000 | 2.1 | 14 | 20 |
| Q9BY41 | Inflammation II | | | | | | | |
| P10767 | Inflammation II | 1562.5 | 3125.0 | 400000 | 800000 | 2.1 | 13 | 16 |
| Q01201 | Inflammation II | | | | | | 12 | 7 |
| P41273 | Inflammation II | | | | | | 12 | 17 |
| P03372 | Inflammation II | | | | | | 8 | 29 |
| Q9UPW0 | Inflammation II | 195.3 | 390.6 | 25000 | 800000 | 1.8 | 12 | 12 |
| P25490 | Inflammation II | | | | | | | |
| Q6R327 | Inflammation II | | | | | | 4 | 17 |
| Q13190 | Inflammation II | | | | | | 7 | 10 |
| Q6UXZ4 | Inflammation II | 48.8 | 97.7 | 12500 | 100000 | 2.1 | 15 | 14 |
| Q9NQI0 | Inflammation II | 195.3 | 195.3 | 12500 | 100000 | 1.8 | | |
| Q8WX93 | Inflammation II | 781.3 | 1562.5 | 100000 | 400000 | 1.8 | 10 | 27 |
| Q16665 | Inflammation II | 12.2 | 24.4 | 1563 | 12500 | 1.8 | 8 | 20 |
| P15927 | Inflammation II | | | | | | | |
| Q99665 | Inflammation II | 3125.0 | 3125.0 | 400000 | 800000 | 2.1 | 2 | 28 |
| O75365 | Inflammation II | | | | | | 8 | 10 |
| P24530 | Inflammation II | 48.8 | 97.7 | 6250 | 25000 | 1.8 | 8 | 20 |
| Q04837 | Inflammation II | | | | | | 12 | 16 |
| O00401 | Inflammation II | | | | | | | |
| Q9NZS2 | Inflammation II | 6.1 | 12.2 | 3125 | 25000 | 2.4 | 9 | 13 |
| Q9NWV8 | Inflammation II | | | | | | 4 | |
| Q7Z698 | Inflammation II | | | | | | 14 | 21 |

| | | | | | | | | |
|---|---|---|---|---|---|---|---|---|
| Q9Y2I7 | Inflammation II | | | | | | 8 | 12 |
| Q9NRR2 | Inflammation II | 390.6 | 390.6 | 25000 | 800000 | 1.8 | 19 | |
| Q96EB6 | Inflammation II | 195.3 | 390.6 | 25000 | 200000 | 1.8 | 9 | 19 |
| Q12888 | Inflammation II | 390.6 | 1562.5 | 400000 | 800000 | 2.4 | 11 | 18 |
| Q9NY59 | Inflammation II | 3125.0 | 6250.0 | 400000 | 800000 | 1.8 | 10 | |
| Q8WVV4 | Inflammation II | 390.6 | 390.6 | 100000 | 200000 | 2.4 | 17 | 24 |
| Q13114-2 | Inflammation II | | | | | | 10 | 13 |
| Q6EBC2 | Inflammation II | 781.3 | 1562.5 | 200000 | 800000 | 2.1 | 8 | 9 |
| Q14511 | Inflammation II | 195.3 | 195.3 | 12500 | 50000 | 1.8 | 11 | |
| Q8NHP1 | Inflammation II | | | | | | 15 | 23 |
| O95429 | Inflammation II | | | | | | 13 | 14 |
| P36897 | Inflammation II | 390.6 | 390.6 | 50000 | 200000 | 2.1 | 1 | |
| Q8IX19 | Inflammation II | 24.4 | 48.8 | 6250 | 25000 | 2.1 | 21 | 28 |
| Q6B9Z1 | Inflammation II | 24.4 | 48.8 | 12500 | 50000 | 2.4 | 13 | 25 |
| Q9Y3D3 | Inflammation II | 3125.0 | 6250.0 | 400000 | 800000 | 1.8 | | |
| Q8IYW5 | Inflammation II | | | | | | | |
| Q96MM7 | Inflammation II | | | | | | 9 | 24 |
| Q9UHN6 | Inflammation II | 48.8 | 195.3 | 25000 | 200000 | 2.1 | 12 | 14 |
| Q5TBC7 | Inflammation II | 48.8 | 97.7 | 12500 | 50000 | 2.1 | 13 | 15 |
| P09923 | Inflammation II | 390.6 | 781.3 | 25000 | 100000 | 1.5 | 10 | 36 |
| Q96D71 | Inflammation II | | | | | | 14 | 16 |
| Q92574 | Inflammation II | 3125.0 | 6250.0 | 400000 | 800000 | 1.8 | 14 | 21 |
| P98170 | Inflammation II | 3125.0 | 3125.0 | 400000 | 800000 | 2.1 | 8 | 15 |
| P36551 | Inflammation II | 6250.0 | 6250.0 | 800000 | 800000 | 2.1 | 9 | 17 |
| O00148 | Inflammation II | 97.7 | 390.6 | 25000 | 200000 | 1.8 | 5 | 16 |
| O43583 | Inflammation II | 390.6 | 781.3 | 25000 | 100000 | 1.5 | 11 | 21 |
| P48730 | Inflammation II | | | | | | 12 | 36 |
| Q9NS62 | Inflammation II | 1562.5 | 3125.0 | 200000 | 800000 | 1.8 | 7 | 35 |
| Q8NI17 | Inflammation II | 1562.5 | 3125.0 | 200000 | 800000 | 1.8 | 11 | 16 |
| Q93062 | Inflammation II | | | | | | 10 | 16 |
| Q9UIK4 | Inflammation II | 1562.5 | 1562.5 | 100000 | 800000 | 1.8 | 18 | 22 |
| Q8NBK3 | Inflammation II | 3125.0 | 6250.0 | 400000 | 800000 | 1.8 | 8 | 17 |
| P35219 | Inflammation II | 3125.0 | 6250.0 | 800000 | 6400000 | 2.1 | | |

| | | | | | | | | |
|---|---|---|---|---|---|---|---|---|
| O60447 | Inflammation II | 195.3 | 390.6 | 100000 | 200000 | 2.4 | 9 | 15 |
| Q8IV38 | Inflammation II | 781.3 | 1562.5 | 100000 | 400000 | 1.8 | 11 | 23 |
| P54274 | Inflammation II | 781.3 | 1562.5 | 50000 | 400000 | 1.5 | 12 | |
| Q9GZN4 | Inflammation II | | | | | | 12 | 12 |
| O75173 | Inflammation II | 781.3 | 781.3 | 100000 | 800000 | 2.1 | 13 | 25 |
| P17643 | Inflammation II | 6250.0 | 12500.0 | 800000 | 800000 | 1.8 | 10 | 16 |
| Q15465 | Inflammation II | | | | | | | |
| Q7Z5L3 | Inflammation II | 1562.5 | 1562.5 | 100000 | 400000 | 1.8 | 10 | 16 |
| Q96EP0 | Inflammation II | | | | | | 11 | 15 |
| Q9NQ66 | Inflammation II | 6250.0 | 6250.0 | 400000 | 800000 | 1.8 | | |
| Q5JS54 | Inflammation II | | | | | | 12 | 12 |
| O43184 | Inflammation II | 390.6 | 781.3 | 50000 | 800000 | 1.8 | 14 | 14 |
| Q9UHA7 | Inflammation II | 195.3 | 195.3 | 25000 | 200000 | 2.1 | 21 | |
| Q15697 | Inflammation II | 390.6 | 390.6 | 25000 | 200000 | 1.8 | 9 | 18 |
| P17050 | Inflammation II | 3125.0 | 6250.0 | 400000 | 800000 | 1.8 | 13 | 17 |
| Q9H0U9 | Inflammation II | 390.6 | 1562.5 | 100000 | 800000 | 1.8 | 9 | 17 |
| P56645 | Inflammation II | | | | | | 16 | 37 |
| P16671 | Inflammation II | 781.3 | 781.3 | 50000 | 400000 | 1.8 | 9 | 14 |
| P26436 | Inflammation II | 12.2 | 48.8 | 6250 | 50000 | 2.1 | 9 | 24 |
| Q96PX8 | Inflammation II | 195.3 | 195.3 | 25000 | 200000 | 2.1 | 11 | 19 |
| P29536 | Inflammation II | | | | | | 17 | 23 |
| P14902 | Inflammation II | 195.3 | 390.6 | 100000 | 400000 | 2.4 | 13 | 17 |
| Q14160 | Inflammation II | | | | | | 16 | 26 |
| Q16718 | Inflammation II | 781.3 | 1562.5 | 200000 | 800000 | 2.1 | 16 | 17 |
| Q15223 | Inflammation II | | | | | | 9 | 13 |
| P11487 | Inflammation II | 1562.5 | 1562.5 | 100000 | 800000 | 1.8 | | |
| P48546 | Inflammation II | 24.4 | 48.8 | 12500 | 50000 | 2.4 | | |
| Q8WV28 | Inflammation II | 12500.0 | 12500.0 | 800000 | 800000 | 1.8 | 16 | 18 |
| O60500 | Inflammation II | 781.3 | 1562.5 | 100000 | 400000 | 1.8 | 12 | 17 |
| Q96PL5 | Inflammation II | 12.2 | 24.4 | 1563 | 6250 | 1.8 | 13 | 15 |
| Q86UE4 | Inflammation II | 781.3 | 781.3 | 50000 | 100000 | 1.8 | | |
| P20701 | Inflammation II | | | | | | 13 | 30 |
| P52630 | Inflammation II | 781.3 | 1562.5 | 100000 | 800000 | 1.8 | 11 | 24 |

| | | | | | | | | |
|---|---|---|---|---|---|---|---|---|
| O43320 | Inflammation II | 6250.0 | 6250.0 | 400000 | 800000 | 1.8 | 17 | 14 |
| P81534 | Inflammation II | | | | | | 6 | 26 |
| Q9Y2X7 | Inflammation II | | | | | | 15 | 17 |
| Q15399 | Inflammation II | 12.2 | 24.4 | 12500 | 100000 | 2.7 | 14 | 17 |
| Q9H3T2 | Inflammation II | 781.3 | 1562.5 | 200000 | 800000 | 2.1 | 10 | 14 |
| P55211 | Inflammation II | | | | | | | |
| Q99584 | Inflammation II | 3.1 | 12.2 | 12500 | 400000 | 3.0 | 10 | 31 |
| Q9UHI8 | Inflammation II | 390.6 | 781.3 | 50000 | 100000 | 1.8 | 14 | 13 |
| P23743 | Inflammation II | 6250.0 | 12500.0 | 800000 | 800000 | 1.8 | 8 | 18 |
| Q99062 | Inflammation II | 97.7 | 195.3 | 12500 | 100000 | 1.8 | 10 | 11 |
| O75688 | Inflammation II | 1562.5 | 3125.0 | 200000 | 800000 | 1.8 | 9 | 17 |
| Q5T2W1 | Inflammation II | 3125.0 | 6250.0 | 400000 | 800000 | 1.8 | 15 | 18 |
| P49757 | Inflammation II | 390.6 | 1562.5 | 100000 | 800000 | 1.8 | 8 | 30 |
| P11234 | Inflammation II | | | | | | 21 | |
| P06213 | Inflammation II | 1562.5 | 1562.5 | 50000 | 400000 | 1.5 | 6 | 6 |
| O95157 | Inflammation II | 390.6 | 390.6 | 25000 | 200000 | 1.8 | 9 | 12 |
| O60437 | Inflammation II | 48.8 | 97.7 | 6250 | 50000 | 1.8 | 9 | 14 |
| Q9H7Z7 | Inflammation II | | | | | | 17 | 27 |
| O15400 | Inflammation II | | | | | | 10 | 19 |
| Q02880 | Inflammation II | 781.3 | 1562.5 | 50000 | 400000 | 1.5 | 11 | 29 |
| P06730 | Inflammation II | 195.3 | 195.3 | 25000 | 100000 | 2.1 | 11 | 37 |
| Q15762 | Inflammation II | | | | | | 15 | 21 |
| Q9BV40 | Inflammation II | 1562.5 | 3125.0 | 400000 | 3200000 | 2.1 | 12 | 30 |
| P49765 | Inflammation II | | | | | | 9 | 12 |
| P32927 | Inflammation II | 97.7 | 97.7 | 12500 | 100000 | 2.1 | 9 | 22 |
| Q5QGZ9 | Inflammation II | | | | | | 8 | 9 |
| Q9NZH8 | Inflammation II | | | | | | 16 | 16 |
| Q16643 | Inflammation II | 24.4 | 48.8 | 3125 | 800000 | 1.8 | 9 | 17 |
| Q9H171 | Inflammation II | 6250.0 | 12500.0 | 800000 | 800000 | 1.8 | 9 | 10 |
| P09564 | Inflammation II | 781.3 | 781.3 | 200000 | 800000 | 2.4 | 15 | 11 |
| O60238 | Inflammation II | 195.3 | 390.6 | 12500 | 100000 | 1.5 | 15 | 18 |
| P24666 | Inflammation II | 6250.0 | 6250.0 | 400000 | 800000 | 1.8 | | |
| Q96PU5 | Inflammation II | | | | | | 8 | 21 |

| | | | | | | | | |
|---|---|---|---|---|---|---|---|---|
| O94916 | Inflammation II | | | | | | 12 | 18 |
| O95835 | Inflammation II | | | | | | 14 | 38 |
| Q8N556 | Inflammation II | | | | | | 6 | 23 |
| P17301 | Inflammation II | | | | | | 8 | 14 |
| Q8NEU8 | Inflammation II | 195.3 | 390.6 | 50000 | 800000 | 2.1 | | |
| Q02223 | Inflammation II | | | | | | 5 | 12 |
| Q5JS37 | Inflammation II | | | | | | 5 | 7 |
| Q6QNK2 | Inflammation II | 48.8 | 97.7 | 12500 | 50000 | 2.1 | 5 | 10 |
| Q8N8U9 | Inflammation II | 24.4 | 48.8 | 25000 | 100000 | 2.7 | 5 | 11 |
| P20908 | Inflammation II | 12.2 | 12.2 | 6250 | 50000 | 2.7 | 6 | 11 |
| Q9BX67 | Inflammation II | 48.8 | 97.7 | 12500 | 100000 | 2.1 | 14 | 33 |
| P22303 | Inflammation II | 24.4 | 48.8 | 12500 | 100000 | 2.4 | 6 | 16 |
| Q9BUH6 | Inflammation II | 97.7 | 195.3 | 25000 | 100000 | 2.1 | 5 | 9 |
| O76074 | Inflammation II | 1562.5 | 3125.0 | 200000 | 800000 | 1.8 | 5 | 27 |
| Q9BRK3 | Inflammation II | 195.3 | 390.6 | 25000 | 200000 | 1.8 | 5 | 14 |
| Q9H7Y0 | Inflammation II | | | | | | 6 | 10 |
| P01037 | Inflammation II | 390.6 | 781.3 | 50000 | 400000 | 1.8 | 6 | 32 |
| P04083 | Inflammation II | | | | | | 7 | 16 |
| Q8TEA8 | Inflammation II | 781.3 | 781.3 | 50000 | 800000 | 1.8 | | |
| Q9ULI3 | Inflammation II | 390.6 | 781.3 | 50000 | 400000 | 1.8 | 5 | 9 |
| P30040 | Inflammation II | 97.7 | 97.7 | 6250 | 12500 | 1.8 | | |
| Q96QR1 | Inflammation II | | | | | | 6 | 13 |
| O75347 | Inflammation II | 24.4 | 48.8 | 6250 | 50000 | 2.1 | 5 | 27 |
| Q9UGN4 | Inflammation II | 48.8 | 97.7 | 6250 | 50000 | 1.8 | 5 | 9 |
| Q8NDA2 | Inflammation II | 390.6 | 781.3 | 100000 | 400000 | 2.1 | 5 | 21 |
| P06280 | Inflammation II | | | | | | 5 | 11 |
| Q6P5S2 | Inflammation II | 12.2 | 48.8 | 6250 | 25000 | 2.1 | 5 | 18 |
| Q16378 | Inflammation II | | | | | | 7 | 18 |
| P61457 | Inflammation II | | | | | | 5 | 12 |
| Q9UI42 | Inflammation II | | | | | | 6 | 13 |
| P27348 | Inflammation II | 390.6 | 390.6 | 25000 | 200000 | 1.8 | 6 | 29 |
| Q99574 | Inflammation II | 195.3 | 195.3 | 6250 | 50000 | 1.5 | 8 | 12 |
| P06132 | Inflammation II | 390.6 | 390.6 | 25000 | 200000 | 1.8 | 6 | 16 |

| | | | | | | | | |
|---|---|---|---|---|---|---|---|---|
| Q5TDH0 | Inflammation II | 195.3 | 390.6 | 50000 | 100000 | 2.1 | 6 | 17 |
| Q9HCU4 | Inflammation II | | | | | | 5 | 8 |
| P40197 | Inflammation II | 390.6 | 781.3 | 50000 | 200000 | 1.8 | 5 | 16 |
| P04406 | Inflammation II | | | | | | 6 | 21 |
| Q9NS98 | Inflammation II | | | | | | 6 | 13 |
| P00325 | Inflammation II | 3125.0 | 6250.0 | 400000 | 800000 | 1.8 | 9 | 19 |
| P55083 | Inflammation II | 390.6 | 781.3 | 50000 | 200000 | 1.8 | 5 | 6 |
| Q8TDQ7 | Inflammation II | 390.6 | 390.6 | 12500 | 50000 | 1.5 | 6 | 15 |
| Q9H939 | Inflammation II | 1562.5 | 3125.0 | 200000 | 800000 | 1.8 | 10 | 37 |
| Q9BUN1 | Inflammation II | | | | | | 5 | 7 |
| O75190 | Inflammation II | 781.3 | 3125.0 | 200000 | 800000 | 1.8 | 6 | 27 |
| P04090 | Inflammation II | 781.3 | 3125.0 | 800000 | 800000 | 2.4 | 20 | 24 |
| O95393 | Inflammation II | 781.3 | 781.3 | 50000 | 100000 | 1.8 | 7 | 11 |
| O43399 | Inflammation II | 781.3 | 781.3 | 50000 | 800000 | 1.8 | 6 | 37 |
| Q96HD1 | Inflammation II | 195.3 | 195.3 | 25000 | 200000 | 2.1 | 5 | 11 |
| Q02952 | Inflammation II | 781.3 | 1562.5 | 100000 | 1600000 | 1.8 | 8 | 10 |
| Q4VCS5 | Inflammation II | 12.2 | 24.4 | 3125 | 12500 | 2.1 | 7 | 20 |
| P10912 | Inflammation II | | | | | | 5 | 10 |
| Q9BXJ0 | Inflammation II | 1562.5 | 3125.0 | 200000 | 1600000 | 1.8 | 5 | 9 |
| P30101 | Inflammation II | | | | | | 11 | 18 |
| P0C862 | Inflammation II | 195.3 | 195.3 | 12500 | 400000 | 1.8 | 4 | 18 |
| P30047 | Inflammation II | | | | | | 5 | 8 |
| Q15276 | Inflammation II | 48.8 | 195.3 | 25000 | 100000 | 2.1 | 7 | 14 |
| O14745 | Inflammation II | | | | | | 14 | 33 |
| Q6FHJ7 | Inflammation II | | | | | | 5 | 10 |
| Q58EX2 | Inflammation II | 195.3 | 390.6 | 25000 | 100000 | 1.8 | 10 | 15 |
| Q9HB71 | Inflammation II | 390.6 | 781.3 | 50000 | 200000 | 1.8 | 6 | 35 |
| P14091 | Inflammation II | 97.7 | 97.7 | 50000 | 200000 | 2.7 | 6 | 13 |
| O60279 | Inflammation II | 195.3 | 390.6 | 25000 | 100000 | 1.8 | 4 | 16 |
| P20155 | Inflammation II | 48.8 | 48.8 | 6250 | 25000 | 2.1 | 5 | 10 |
| A6NC86 | Inflammation II | 390.6 | 781.3 | 50000 | 200000 | 1.8 | 11 | 16 |
| A2VDF0 | Inflammation II | 48.8 | 195.3 | 12500 | 100000 | 1.8 | 5 | 22 |
| P49862 | Inflammation II | | | | | | 6 | 14 |

| | | | | | | | | |
|---|---|---|---|---|---|---|---|---|
| P09529 | Inflammation II | 390.6 | 390.6 | 25000 | 100000 | 1.8 | 7 | 18 |
| P25686 | Inflammation II | 195.3 | 390.6 | 25000 | 100000 | 1.8 | 6 | 11 |
| Q86SX6 | Inflammation II | 48.8 | 97.7 | 6250 | 50000 | 1.8 | | |
| P22455 | Inflammation II | 390.6 | 781.3 | 50000 | 100000 | 1.8 | 5 | 13 |
| Q53FA7 | Inflammation II | 6.1 | 24.4 | 6250 | 50000 | 2.4 | 5 | 10 |
| O96007 | Inflammation II | 1562.5 | 3125.0 | 200000 | 800000 | 1.8 | 7 | 11 |
| Q8NHV1 | Inflammation II | 12.2 | 48.8 | 12500 | 100000 | 2.4 | 7 | 11 |
| Q9NQ48 | Inflammation II | 24.4 | 97.7 | 50000 | 200000 | 2.7 | 9 | 24 |
| Q96AJ9 | Inflammation II | 195.3 | 195.3 | 12500 | 25000 | 1.8 | 9 | 13 |
| Q6UX06 | Inflammation II | 781.3 | 1562.5 | 400000 | 800000 | 2.4 | 6 | 34 |
| Q96A49 | Inflammation II | | | | | | 9 | 24 |
| P01210 | Inflammation II | 3125.0 | 3125.0 | 200000 | 400000 | 1.8 | 6 | 8 |
| Q92619 | Inflammation II | 1562.5 | 1562.5 | 400000 | 800000 | 2.4 | 9 | 16 |
| P21854 | Inflammation II | 97.7 | 195.3 | 12500 | 100000 | 1.8 | 6 | 14 |
| P52758 | Inflammation II | 48.8 | 97.7 | 3125 | 12500 | 1.5 | 5 | 10 |
| Q9Y4D1 | Inflammation II | 6250.0 | 6250.0 | 400000 | 800000 | 1.8 | 9 | 27 |
| Q9UK23 | Inflammation II | 12.2 | 48.8 | 6250 | 50000 | 2.1 | 5 | 9 |
| Q8IUZ5 | Inflammation II | 3125.0 | 3125.0 | 400000 | 800000 | 2.1 | 8 | 14 |
| Q9NRS6 | Inflammation II | 195.3 | 195.3 | 12500 | 100000 | 1.8 | 11 | 16 |
| Q5VTT5 | Inflammation II | 12.2 | 24.4 | 6250 | 50000 | 2.4 | 5 | 36 |
| P37840 | Inflammation II | | | | | | | |
| P07311 | Inflammation II | 12.2 | 48.8 | 3125 | 12500 | 1.8 | 5 | 15 |
| Q04323 | Inflammation II | | | | | | 10 | 21 |
| Q6P589 | Inflammation II | | | | | | 8 | 22 |
| P32321 | Inflammation II | 195.3 | 390.6 | 25000 | 100000 | 1.8 | 6 | 24 |
| P56192 | Inflammation II | 1562.5 | 3125.0 | 400000 | 800000 | 2.1 | 6 | 16 |
| O15335 | Inflammation II | 195.3 | 390.6 | 25000 | 100000 | 1.8 | 5 | 18 |
| Q9P2T1 | Inflammation II | 97.7 | 195.3 | 25000 | 100000 | 2.1 | 6 | 15 |
| P35611 | Inflammation II | | | | | | 10 | 21 |
| P54764 | Inflammation II | | | | | | 6 | 9 |
| Q13976 | Inflammation II | | | | | | 5 | 31 |
| Q8N0X7 | Inflammation II | 97.7 | 390.6 | 50000 | 400000 | 2.1 | 6 | 16 |
| Q6GMV3 | Inflammation II | 390.6 | 390.6 | 50000 | 800000 | 2.1 | 7 | 26 |

| | | | | | | | | |
|---|---|---|---|---|---|---|---|---|
| P50502 | Inflammation II | 390.6 | 390.6 | 50000 | 800000 | 2.1 | 5 | 9 |
| P32119 | Inflammation II | | | | | | 8 | 12 |
| P0DML2 | Inflammation II | 97.7 | 97.7 | 12500 | 50000 | 2.1 | | |
| P40925 | Inflammation II | 1562.5 | 6250.0 | 400000 | 800000 | 1.8 | 4 | 8 |
| P10599 | Inflammation II | 48.8 | 97.7 | 6250 | 12500 | 1.8 | 24 | 23 |
| Q92686 | Inflammation II | 48.8 | 48.8 | 12500 | 800000 | 2.4 | 7 | 28 |
| Q08830 | Inflammation II | 390.6 | 390.6 | 100000 | 200000 | 2.4 | 4 | 27 |
| Q13790 | Inflammation II | | | | | | 6 | 20 |
| P07333 | Inflammation II | 97.7 | 195.3 | 12500 | 50000 | 1.8 | 5 | 9 |
| P00390 | Inflammation II | 195.3 | 390.6 | 25000 | 100000 | 1.8 | 5 | 6 |
| P0DJD7 | Inflammation II | 12.2 | 24.4 | 25000 | 100000 | 3.0 | 4 | 9 |
| P02748 | Inflammation II | | | | | | 19 | 28 |
| P17927 | Inflammation II | 6.1 | 6.1 | 6250 | 50000 | 3.0 | 6 | 9 |
| P12821 | Inflammation II | 390.6 | 1562.5 | 200000 | 800000 | 2.1 | 7 | 9 |
| Q6UXB8 | Inflammation II | | | | | | 6 | 6 |
| P22897 | Inflammation II | 24.4 | 48.8 | 12500 | 200000 | 2.4 | 5 | 9 |
| P16233 | Inflammation II | 6.1 | 12.2 | 12500 | 50000 | 3.0 | 5 | 13 |
| P50552 | Inflammation II | 6250.0 | 12500.0 | 800000 | 800000 | 1.8 | 9 | 36 |
| P04180 | Inflammation II | 390.6 | 781.3 | 50000 | 400000 | 1.8 | 5 | 9 |
| O43493 | Inflammation II | 97.7 | 195.3 | 12500 | 800000 | 1.8 | 7 | 10 |
| O00602 | Inflammation II | | | | | | 11 | 22 |
| Q93091 | Inflammation II | 6.1 | 24.4 | 3125 | 12500 | 2.1 | 6 | 7 |
| P12955 | Inflammation II | 195.3 | 390.6 | 25000 | 200000 | 1.8 | 6 | 9 |
| Q0ZGT2 | Inflammation II | 48.8 | 97.7 | 25000 | 200000 | 2.4 | | |
| P20061 | Inflammation II | 12.2 | 24.4 | 6250 | 50000 | 2.4 | 5 | 10 |
| P02652 | Inflammation II | | | | | | | |
| P04278 | Inflammation II | 1562.5 | 3125.0 | 400000 | 800000 | 2.1 | 3 | 10 |
| P00742 | Inflammation II | 97.7 | 195.3 | 12500 | 50000 | 1.8 | 8 | 9 |
| P07307 | Inflammation II | 195.3 | 390.6 | 25000 | 200000 | 1.8 | 8 | 14 |
| P08294 | Inflammation II | 3.1 | 12.2 | 1563 | 12500 | 2.1 | 4 | 11 |
| Q86YW5 | Inflammation II | 97.7 | 390.6 | 25000 | 200000 | 1.8 | 8 | 29 |
| P09172 | Inflammation II | 97.7 | 195.3 | 100000 | 400000 | 2.7 | 4 | 32 |
| P27169 | Inflammation II | | | | | | 7 | 11 |

| | | | | | | | | |
|---|---|---|---|---|---|---|---|---|
| P16442 | Inflammation II | 12.2 | 48.8 | 50000 | 200000 | 3.0 | 8 | 9 |
| O95497 | Inflammation II | 24.4 | 48.8 | 6250 | 50000 | 2.1 | 4 | 20 |
| Q04756 | Inflammation II | 48.8 | 195.3 | 12500 | 50000 | 1.8 | 4 | 7 |
| P06276 | Inflammation II | | | | | | 4 | 12 |
| Q9BXR6 | Inflammation II | | | | | | 6 | 15 |
| P54108 | Inflammation II | 390.6 | 390.6 | 25000 | 50000 | 1.8 | 3 | 7 |
| P06396 | Inflammation II | 1562.5 | 3125.0 | 200000 | 800000 | 1.8 | 4 | 8 |
| P0DN86 | Inflammation II | | | | | | 13 | |
| P26927 | Inflammation II | 97.7 | 195.3 | 25000 | 200000 | 2.1 | 4 | 14 |
| P04040 | Inflammation II | | | | | | 7 | 11 |
| P34096 | Inflammation II | 781.3 | 781.3 | 50000 | 200000 | 1.8 | 6 | 9 |
| P07998 | Inflammation II | | | | | | 10 | 11 |
| Q01459 | Inflammation II | 195.3 | 195.3 | 12500 | 50000 | 1.8 | 4 | 5 |
| P06744 | Inflammation II | 195.3 | 390.6 | 50000 | 200000 | 2.1 | 7 | 16 |
| P04114 | Inflammation II | | | | | | 9 | 29 |
| P01019 | Inflammation II | 1562.5 | 3125.0 | 200000 | 800000 | 1.8 | 6 | 31 |
| P0DUB6_P0 | Inflammation II | 195.3 | 195.3 | 12500 | 50000 | 1.8 | 6 | 12 |
| P02765 | Inflammation II | | | | | | 5 | 14 |
| O14791 | Inflammation II | 97.7 | 97.7 | 25000 | 100000 | 2.4 | 4 | 33 |
| P07358 | Inflammation II | | | | | | 12 | 14 |
| P36980 | Inflammation II | 195.3 | 390.6 | 25000 | 100000 | 1.8 | 8 | 16 |
| P08185 | Inflammation II | 390.6 | 781.3 | 200000 | 800000 | 2.4 | 5 | 8 |
| P05543 | Inflammation II | 195.3 | 390.6 | 25000 | 200000 | 1.8 | 7 | 8 |
| P36955 | Inflammation II | 195.3 | 390.6 | 50000 | 800000 | 2.1 | 3 | 8 |
| Q96PD5 | Inflammation II | 1562.5 | 3125.0 | 400000 | 800000 | 2.1 | 4 | 6 |
| Q08380 | Inflammation II | 97.7 | 195.3 | 12500 | 50000 | 1.8 | 7 | 11 |
| P10909 | Inflammation II | 781.3 | 1562.5 | 100000 | 800000 | 1.8 | 5 | 8 |
| P61769 | Inflammation II | 48.8 | 97.7 | 6250 | 25000 | 1.8 | 6 | 5 |
| P43652 | Inflammation II | 97.7 | 195.3 | 50000 | 200000 | 2.4 | 6 | 7 |
| Q92496 | Inflammation II | 48.8 | 97.7 | 12500 | 50000 | 2.1 | 4 | 12 |
| P35542 | Inflammation II | | | | | | 5 | 9 |
| P0DOY2 | Inflammation II | 6.1 | 12.2 | 781 | 50000 | 1.8 | 4 | 5 |
| P00748 | Inflammation II | 97.7 | 195.3 | 25000 | 200000 | 2.1 | 9 | 23 |

| | | | | | | | | |
|---|---|---|---|---|---|---|---|---|
| P11226 | Inflammation II | 1.5 | 12.2 | 3125 | 25000 | 2.4 | 5 | 27 |
| P00734 | Inflammation II | | | | | | 6 | 7 |
| P02649 | Inflammation II | | | | | | 10 | 17 |
| Q9Y5Y7 | Inflammation II | 3.1 | 6.1 | 1563 | 12500 | 2.4 | 6 | 11 |
| P00746 | Inflammation II | 12.2 | 24.4 | 12500 | 50000 | 2.7 | 7 | 6 |
| P07225 | Inflammation II | 195.3 | 390.6 | 50000 | 200000 | 2.1 | 5 | 7 |
| P03952 | Inflammation II | 195.3 | 390.6 | 25000 | 200000 | 1.8 | 5 | 7 |
| Q16610 | Inflammation II | 6250.0 | 6250.0 | 400000 | 800000 | 1.8 | 7 | 15 |
| O75882-2 | Inflammation II | 195.3 | 390.6 | 50000 | 200000 | 2.1 | 5 | 6 |
| P05154 | Inflammation II | | | | | | 5 | 14 |
| O00391 | Inflammation II | 390.6 | 390.6 | 12500 | 50000 | 1.5 | 6 | 7 |
| P02775 | Inflammation II | 24.4 | 48.8 | 12500 | 100000 | 2.4 | 5 | 28 |
| Q96IY4 | Inflammation II | 195.3 | 390.6 | 25000 | 200000 | 1.8 | 6 | 9 |
| P20742 | Inflammation II | 390.6 | 390.6 | 100000 | 800000 | 2.4 | 4 | 6 |
| P06727 | Inflammation II | 781.3 | 1562.5 | 100000 | 800000 | 1.8 | 8 | 16 |
| P09871 | Inflammation II | 195.3 | 390.6 | 50000 | 200000 | 2.1 | 6 | 7 |
| P02776 | Inflammation II | | | | | | 11 | 28 |
| P05452 | Inflammation II | 97.7 | 195.3 | 25000 | 200000 | 2.1 | 5 | 6 |
| Q14624 | Inflammation II | 390.6 | 781.3 | 50000 | 200000 | 1.8 | 4 | 14 |
| P05546 | Inflammation II | 3125.0 | 3125.0 | 400000 | 800000 | 2.1 | 5 | 11 |
| P02647 | Inflammation II | | | | | | 8 | 14 |
| Q15848 | Inflammation II | 781.3 | 781.3 | 25000 | 50000 | 1.5 | 7 | 18 |
| P10643 | Inflammation II | 24.4 | 48.8 | 12500 | 100000 | 2.4 | 9 | 6 |
| P08519 | Inflammation II | 24.4 | 48.8 | 3125 | 50000 | 1.8 | | |
| P00736 | Inflammation II | 390.6 | 1562.5 | 100000 | 200000 | 1.8 | 5 | 6 |
| P05090 | Inflammation II | 781.3 | 1562.5 | 50000 | 200000 | 1.5 | 7 | 17 |
| P14151 | Inflammation II | 3.1 | 6.1 | 1563 | 6250 | 2.4 | 4 | 6 |
| P43251 | Inflammation II | 12.2 | 24.4 | 3125 | 25000 | 2.1 | 5 | 7 |
| Q9NZP8 | Inflammation II | 12.2 | 24.4 | 6250 | 50000 | 2.4 | 5 | 7 |
| P02763 | Inflammation II | 24.4 | 48.8 | 6250 | 25000 | 2.1 | 6 | 5 |
| P19827 | Inflammation II | 195.3 | 195.3 | 25000 | 200000 | 2.1 | 6 | 8 |
| P02654 | Inflammation II | 195.3 | 390.6 | 100000 | 800000 | 2.4 | 9 | 16 |
| P00751 | Inflammation II | 781.3 | 1562.5 | 100000 | 800000 | 1.8 | 6 | 12 |

| | | | | | | | | |
|---|---|---|---|---|---|---|---|---|
| P01009 | Inflammation II | 3125.0 | 6250.0 | 400000 | 800000 | 1.8 | 3 | 4 |
| P49908 | Inflammation II | 97.7 | 195.3 | 25000 | 100000 | 2.1 | 15 | 19 |
| P01031 | Inflammation II | 24.4 | 48.8 | 3125 | 12500 | 1.8 | 7 | 7 |
| P02750 | Inflammation II | 97.7 | 97.7 | 6250 | 25000 | 1.8 | 5 | 6 |
| P04196 | Inflammation II | 97.7 | 195.3 | 25000 | 200000 | 2.1 | 6 | 9 |
| P00747 | Inflammation II | 97.7 | 195.3 | 50000 | 200000 | 2.4 | 4 | 11 |
| P27918 | Inflammation II | 195.3 | 390.6 | 25000 | 200000 | 1.8 | 4 | 7 |
| P02787 | Inflammation II | | | | | | 5 | 5 |
| P02751 | Inflammation II | | | | | | 4 | 23 |
| P02766 | Inflammation II | 195.3 | 195.3 | 12500 | 50000 | 1.8 | 5 | 5 |
| P05160 | Inflammation II | 390.6 | 781.3 | 25000 | 100000 | 1.5 | 6 | 10 |
| P02774 | Inflammation II | 97.7 | 390.6 | 25000 | 200000 | 1.8 | 4 | 7 |
| P08697 | Inflammation II | 390.6 | 781.3 | 100000 | 400000 | 2.1 | 4 | 6 |
| P02671 | Inflammation II | 781.3 | 1562.5 | 200000 | 800000 | 2.1 | 6 | 19 |
| P05155 | Inflammation II | 390.6 | 390.6 | 25000 | 200000 | 1.8 | 4 | 7 |
| P01008 | Inflammation II | | | | | | 4 | 4 |
| P01024 | Inflammation II | | | | | | 7 | 40 |
| O43866 | Inflammation II | 24.4 | 24.4 | 12500 | 50000 | 2.7 | 4 | 13 |
| P29622 | Inflammation II | | | | | | 5 | 6 |
| P02743 | Inflammation II | 195.3 | 390.6 | 25000 | 100000 | 1.8 | 4 | 10 |
| P01011 | Inflammation II | | | | | | 3 | 6 |
| P04217 | Inflammation II | 48.8 | 195.3 | 12500 | 200000 | 1.8 | 4 | 6 |
| P08603 | Inflammation II | | | | | | 5 | 8 |
| P03951 | Inflammation II | 12.2 | 48.8 | 12500 | 50000 | 2.4 | 6 | 9 |
| P05156 | Inflammation II | 195.3 | 390.6 | 50000 | 200000 | 2.1 | 5 | 9 |
| A1E959 | Neurology | 195.3 | 781.3 | 100000 | 200000 | 2.1 | 8 | 14 |
| P06748 | Neurology | 24.4 | 48.8 | 12500 | 200000 | 2.4 | 8 | 10 |
| Q9NRG1 | Neurology | 3125.0 | 6250.0 | 400000 | 800000 | 1.8 | 10 | 11 |
| P58417 | Neurology | 48.8 | 195.3 | 100000 | 200000 | 2.7 | 8 | 13 |
| Q9H3R2 | Neurology | 6.1 | 12.2 | 6250 | 25000 | 2.7 | 9 | 19 |
| Q6UX27 | Neurology | 48.8 | 97.7 | 6250 | 25000 | 1.8 | 9 | 9 |
| Q15043 | Neurology | 97.7 | 390.6 | 12500 | 50000 | 1.5 | 9 | 19 |
| Q9HA65 | Neurology | 195.3 | 390.6 | 50000 | 200000 | 2.1 | 8 | 12 |

| | | | | | | | | |
|---|---|---|---|---|---|---|---|---|
| O60242 | Neurology | 12.2 | 24.4 | 100000 | 800000 | 3.6 | 7 | 6 |
| Q06323 | Neurology | 97.7 | 195.3 | 50000 | 200000 | 2.4 | 8 | 8 |
| Q92765 | Neurology | 97.7 | 195.3 | 25000 | 50000 | 2.1 | 7 | 5 |
| P16278 | Neurology | 195.3 | 390.6 | 100000 | 800000 | 2.4 | 8 | 10 |
| Q9H1C3 | Neurology | 3125.0 | 6250.0 | 1600000 | 3200000 | 2.4 | 9 | 9 |
| Q9NR71 | Neurology | 97.7 | 195.3 | 25000 | 200000 | 2.1 | 11 | 6 |
| O95994 | Neurology | 1562.5 | 1562.5 | 100000 | 800000 | 1.8 | 21 | 13 |
| O60609 | Neurology | 24.4 | 48.8 | 12500 | 50000 | 2.4 | 9 | 8 |
| Q9HD42 | Neurology | | | | | | 5 | 10 |
| Q16762 | Neurology | 6250.0 | 6250.0 | 800000 | 1600000 | 2.1 | 11 | 13 |
| P61244 | Neurology | 97.7 | 390.6 | 25000 | 100000 | 1.8 | 7 | 19 |
| P09211 | Neurology | 12500.0 | 12500.0 | 400000 | 1600000 | 1.5 | 8 | 17 |
| Q6UXK2 | Neurology | 24.4 | 48.8 | 25000 | 200000 | 2.7 | 6 | 15 |
| P28325 | Neurology | 24.4 | 24.4 | 6250 | 25000 | 2.4 | 6 | 6 |
| P78423 | Neurology | 24.4 | 48.8 | 6250 | 12500 | 2.1 | 8 | 8 |
| P29466 | Neurology | 97.7 | 195.3 | 50000 | 100000 | 2.4 | 8 | 21 |
| P07196 | Neurology | 390.6 | 781.3 | 200000 | 800000 | 2.4 | 12 | 12 |
| Q9Y2W6 | Neurology | 6.1 | 12.2 | 6250 | 25000 | 2.7 | 6 | 13 |
| O94985 | Neurology | 390.6 | 1562.5 | 200000 | 800000 | 2.1 | 10 | 11 |
| Q6UWL6 | Neurology | 97.7 | 195.3 | 50000 | 200000 | 2.4 | 8 | 14 |
| Q9UHV9 | Neurology | 97.7 | 195.3 | 100000 | 800000 | 2.7 | 7 | 23 |
| P50135 | Neurology | 12.2 | 48.8 | 25000 | 50000 | 2.7 | 7 | 6 |
| Q9H3S3 | Neurology | 3.1 | 6.1 | 6250 | 50000 | 3.0 | 8 | 5 |
| O14763 | Neurology | 0.8 | 1.5 | 3125 | 12500 | 3.3 | 10 | 7 |
| O15496 | Neurology | 1.5 | 3.1 | 3125 | 12500 | 3.0 | 9 | 6 |
| Q9Y6Y9 | Neurology | 6250.0 | 12500.0 | 1600000 | 3200000 | 2.1 | 9 | 6 |
| O00559 | Neurology | 97.7 | 390.6 | 50000 | 200000 | 2.1 | 8 | 6 |
| P11464 | Neurology | 6.1 | 12.2 | 3125 | 12500 | 2.4 | 8 | 7 |
| P15509 | Neurology | 781.3 | 1562.5 | 400000 | 800000 | 2.4 | 9 | 8 |
| P42892 | Neurology | | | | | | 13 | 17 |
| Q96NZ8 | Neurology | 12.2 | 24.4 | 50000 | 200000 | 3.3 | 10 | 5 |
| P78333 | Neurology | 12.2 | 48.8 | 25000 | 50000 | 2.7 | 10 | 6 |
| P78560 | Neurology | | | | | | 10 | 6 |

| | | | | | | | | |
|---|---|---|---|---|---|---|---|---|
| O14618 | Neurology | 781.3 | 1562.5 | 400000 | 800000 | 2.4 | 9 | 10 |
| P07306 | Neurology | 195.3 | 390.6 | 100000 | 800000 | 2.4 | 7 | 6 |
| P13500 | Neurology | 6.1 | 12.2 | 1563 | 6250 | 2.1 | 9 | 8 |
| P02533 | Neurology | 97.7 | 390.6 | 200000 | 800000 | 2.7 | 6 | 18 |
| Q9NRW1 | Neurology | 195.3 | 781.3 | 50000 | 200000 | 1.8 | 8 | 11 |
| Q6ZMC9 | Neurology | 97.7 | 195.3 | 100000 | 200000 | 2.7 | 6 | 15 |
| P53582 | Neurology | 781.3 | 1562.5 | 200000 | 800000 | 2.1 | 7 | 23 |
| Q6ZMJ4 | Neurology | 48.8 | 195.3 | 200000 | 800000 | 3.0 | 8 | 14 |
| Q8IU54 | Neurology | 6.1 | 12.2 | 25000 | 100000 | 3.3 | 9 | 16 |
| Q8N2G4 | Neurology | 195.3 | 390.6 | 100000 | 400000 | 2.4 | 6 | 12 |
| P20936 | Neurology | 6250.0 | 6250.0 | 3200000 | 6400000 | 2.7 | 9 | 15 |
| P48047 | Neurology | 48.8 | 48.8 | 100000 | 800000 | 3.3 | 6 | 20 |
| Q9BXS1 | Neurology | 195.3 | 781.3 | 200000 | 800000 | 2.4 | 12 | 20 |
| Q4LE39 | Neurology | 12.2 | 24.4 | 12500 | 200000 | 2.7 | 6 | 15 |
| P05455 | Neurology | 390.6 | 1562.5 | 200000 | 800000 | 2.1 | 6 | 10 |
| Q6P1M0 | Neurology | 24.4 | 24.4 | 100000 | 200000 | 3.6 | 8 | 18 |
| Q9NXA8 | Neurology | 1562.5 | 1562.5 | 200000 | 400000 | 2.1 | 12 | 24 |
| Q6PIL6 | Neurology | 48.8 | 48.8 | 12500 | 200000 | 2.4 | 8 | 8 |
| P53985 | Neurology | 781.3 | 1562.5 | 400000 | 800000 | 2.4 | 13 | 9 |
| Q86VW0 | Neurology | 3125.0 | 3125.0 | 400000 | 800000 | 2.1 | 14 | 6 |
| P49023 | Neurology | | | | | | 7 | 14 |
| P51531 | Neurology | 97.7 | 195.3 | 50000 | 200000 | 2.4 | 7 | 12 |
| P08758 | Neurology | | | | | | 10 | 17 |
| P15018 | Neurology | 3.1 | 6.1 | 3125 | 100000 | 2.7 | 6 | 9 |
| P13385 | Neurology | 390.6 | 781.3 | 100000 | 200000 | 2.1 | 8 | 34 |
| P15336 | Neurology | 1406.3 | 2812.5 | 360000 | 720000 | 2.1 | 8 | 16 |
| O96013 | Neurology | 390.6 | 1562.5 | 200000 | 800000 | 2.1 | 12 | 15 |
| Q96JP9 | Neurology | 48.8 | 97.7 | 50000 | 200000 | 2.7 | 8 | 10 |
| P45452 | Neurology | 24.4 | 48.8 | 100000 | 400000 | 3.3 | 8 | 13 |
| P10636 | Neurology | 1562.5 | 1562.5 | 50000 | 200000 | 1.5 | 11 | 22 |
| O14579 | Neurology | 781.3 | 1562.5 | 400000 | 800000 | 2.4 | 8 | 12 |
| P09769 | Neurology | 390.6 | 390.6 | 100000 | 400000 | 2.4 | 8 | 19 |
| P53539 | Neurology | 195.3 | 390.6 | 200000 | 800000 | 2.7 | 6 | 22 |

| | | | | | | | | |
|---|---|---|---|---|---|---|---|---|
| Q9HAW4 | Neurology | | | | | | 17 | 32 |
| O95466 | Neurology | 3125.0 | 3125.0 | 200000 | 800000 | 1.8 | 11 | 19 |
| Q96FQ6 | Neurology | 97.7 | 390.6 | 25000 | 400000 | 1.8 | 13 | 13 |
| O00399 | Neurology | 781.3 | 3125.0 | 200000 | 800000 | 1.8 | 5 | 14 |
| Q8WTV0 | Neurology | 1562.5 | 3125.0 | 400000 | 800000 | 2.1 | 7 | 14 |
| P22676 | Neurology | 195.3 | 781.3 | 200000 | 400000 | 2.4 | 9 | 19 |
| Q9UBB4 | Neurology | 195.3 | 390.6 | 50000 | 200000 | 2.1 | 9 | 13 |
| Q9H2W6 | Neurology | | | | | | 11 | 21 |
| Q07812 | Neurology | 14.2 | 28.3 | 29000 | 116000 | 3.0 | 9 | 21 |
| Q16864 | Neurology | | | | | | 7 | 16 |
| Q9BW30 | Neurology | 195.3 | 390.6 | 25000 | 100000 | 1.8 | 8 | 10 |
| P23276 | Neurology | 1562.5 | 3125.0 | 200000 | 800000 | 1.8 | 6 | 8 |
| Q92597 | Neurology | 48.8 | 195.3 | 100000 | 400000 | 2.7 | 9 | 15 |
| P26440 | Neurology | 3125.0 | 3125.0 | 400000 | 800000 | 2.1 | 10 | 12 |
| Q9UM07 | Neurology | | | | | | 9 | 14 |
| Q9UJY5 | Neurology | | | | | | 11 | 11 |
| P14625 | Neurology | | | | | | 7 | 11 |
| Q5JZY3 | Neurology | 97.7 | 195.3 | 100000 | 800000 | 2.7 | 8 | 15 |
| Q8WUW1 | Neurology | 24.4 | 48.8 | 100000 | 200000 | 3.3 | 9 | 13 |
| Q15126 | Neurology | 3125.0 | 3125.0 | 800000 | 6400000 | 2.4 | 17 | 21 |
| P30519 | Neurology | 24.4 | 48.8 | 100000 | 200000 | 3.3 | 8 | 10 |
| Q86Z14 | Neurology | 1562.5 | 1562.5 | 200000 | 800000 | 2.1 | 12 | 22 |
| Q9H0C8 | Neurology | 781.3 | 1562.5 | 400000 | 800000 | 2.4 | 10 | 17 |
| O14523 | Neurology | 25000.0 | 50000.0 | 6400000 | 12800000 | 2.1 | 5 | |
| O94813 | Neurology | 97.7 | 390.6 | 100000 | 200000 | 2.4 | 7 | 11 |
| P13647 | Neurology | | | | | | 11 | 17 |
| Q9NS71 | Neurology | 1562.5 | 3125.0 | 800000 | 3200000 | 2.4 | 6 | 26 |
| Q13308 | Neurology | 390.6 | 390.6 | 100000 | 200000 | 2.4 | 7 | 11 |
| Q9UKV5 | Neurology | 97.7 | 195.3 | 100000 | 400000 | 2.7 | 7 | 21 |
| Q9NSK7 | Neurology | 48.8 | 195.3 | 100000 | 200000 | 2.7 | 7 | 18 |
| P29474 | Neurology | 24.4 | 48.8 | 100000 | 200000 | 3.3 | 10 | 14 |
| P60484 | Neurology | | | | | | 10 | 11 |
| Q15633 | Neurology | 97.7 | 195.3 | 100000 | 200000 | 2.7 | 7 | 19 |

| | | | | | | | | |
|---|---|---|---|---|---|---|---|---|
| Q9Y5P4 | Neurology | | | | | | 10 | 13 |
| Q9UPY6 | Neurology | | | | | | 6 | 9 |
| P01375 | Neurology | 1.5 | 6.1 | 6250 | 200000 | 3.0 | 10 | 16 |
| P29475 | Neurology | 6.1 | 6.1 | 25000 | 50000 | 3.6 | 11 | 21 |
| P51452 | Neurology | | | | | | 9 | 10 |
| P41208 | Neurology | 3125.0 | 3125.0 | 200000 | 800000 | 1.8 | 11 | 20 |
| O60240 | Neurology | | | | | | 6 | 9 |
| Q9Y6E0 | Neurology | 24.4 | 195.3 | 50000 | 200000 | 2.4 | 5 | 18 |
| P30039 | Neurology | 390.6 | 781.3 | 400000 | 800000 | 2.7 | 7 | 12 |
| Q03393 | Neurology | 24.4 | 48.8 | 25000 | 200000 | 2.7 | 8 | 11 |
| Q13426 | Neurology | 390.6 | 781.3 | 100000 | 200000 | 2.1 | 7 | 11 |
| P39905 | Neurology | 0.4 | 0.8 | 1563 | 12500 | 3.3 | 7 | 22 |
| Q9UK53 | Neurology | 48.8 | 97.7 | 100000 | 200000 | 3.0 | 8 | 15 |
| Q02643 | Neurology | 3125.0 | 3125.0 | 400000 | 800000 | 2.1 | 9 | 17 |
| Q9Y2V2 | Neurology | 781.3 | 1562.5 | 100000 | 400000 | 1.8 | 11 | 17 |
| Q9UJ72 | Neurology | 48.8 | 97.7 | 25000 | 200000 | 2.4 | 8 | 19 |
| Q8NBI3 | Neurology | 97.7 | 195.3 | 50000 | 200000 | 2.4 | 8 | 9 |
| P50453 | Neurology | 1562.5 | 3125.0 | 400000 | 800000 | 2.1 | 8 | 7 |
| P52798 | Neurology | 6.1 | 6.1 | 3125 | 12500 | 2.7 | 7 | 5 |
| Q92932 | Neurology | 12.2 | 48.8 | 25000 | 50000 | 2.7 | 9 | 7 |
| Q02083 | Neurology | 97.7 | 195.3 | 100000 | 800000 | 2.7 | 7 | 7 |
| Q9H5V8 | Neurology | 48.8 | 97.7 | 6250 | 12500 | 1.8 | 7 | 6 |
| Q16740 | Neurology | 48.8 | 195.3 | 100000 | 800000 | 2.7 | 7 | 10 |
| P52943 | Neurology | 781.3 | 1562.5 | 200000 | 800000 | 2.1 | 7 | 6 |
| P36269 | Neurology | 781.3 | 1562.5 | 400000 | 800000 | 2.4 | 9 | 6 |
| Q9Y6D9 | Neurology | | | | | | 9 | 5 |
| P63098 | Neurology | | | | | | 8 | 7 |
| O95256 | Neurology | 12.2 | 12.2 | 6250 | 50000 | 2.7 | 9 | 5 |
| Q9HAN9 | Neurology | | | | | | 7 | 7 |
| Q96IU4 | Neurology | | | | | | 8 | 16 |
| O43155 | Neurology | 12.2 | 48.8 | 25000 | 100000 | 2.7 | 9 | 8 |
| P07741 | Neurology | 390.6 | 781.3 | 400000 | 800000 | 2.7 | 10 | 6 |
| O00220 | Neurology | 6.1 | 12.2 | 6250 | 25000 | 2.7 | 10 | 6 |

| | | | | | | | | |
|---|---|---|---|---|---|---|---|---|
| Q96PP9 | Neurology | 1562.5 | 6250.0 | 800000 | 3200000 | 2.1 | 9 | 10 |
| Q92917 | Neurology | 3125.0 | 6250.0 | 400000 | 3200000 | 1.8 | 8 | 10 |
| Q8IUN9 | Neurology | 12.2 | 48.8 | 12500 | 50000 | 2.4 | 7 | 8 |
| Q96B36 | Neurology | 546.9 | 1093.8 | 70000 | 280000 | 1.8 | 9 | 16 |
| Q92851 | Neurology | 97.7 | 195.3 | 50000 | 200000 | 2.4 | 7 | 12 |
| O95817 | Neurology | 97.7 | 97.7 | 50000 | 200000 | 2.7 | 9 | 6 |
| O95630 | Neurology | 24.4 | 48.8 | 25000 | 200000 | 2.7 | 10 | 7 |
| Q2TAL6 | Neurology | 97.7 | 195.3 | 25000 | 50000 | 2.1 | 8 | 6 |
| Q9UBT3 | Neurology | | | | | | 9 | 5 |
| O00308 | Neurology | 97.7 | 390.6 | 400000 | 800000 | 3.0 | 11 | 10 |
| P09466 | Neurology | 46.4 | 92.8 | 47500 | 380000 | 2.7 | 10 | 13 |
| Q08629 | Neurology | 390.6 | 390.6 | 200000 | 800000 | 2.7 | 9 | 5 |
| Q6UX15 | Neurology | 3.1 | 6.1 | 6250 | 50000 | 3.0 | 9 | 6 |
| Q53H47 | Neurology | 390.6 | 1562.5 | 400000 | 800000 | 2.4 | 8 | 4 |
| P49789 | Neurology | 6.1 | 12.2 | 1563 | 6250 | 2.1 | 10 | 14 |
| P05231 | Neurology | 0.8 | 1.5 | 1563 | 12500 | 3.0 | 10 | 8 |
| P18031 | Neurology | | | | | | 8 | 12 |
| P29218 | Neurology | 195.3 | 781.3 | 200000 | 800000 | 2.4 | 9 | 9 |
| P14868 | Neurology | 48.8 | 97.7 | 100000 | 400000 | 3.0 | 10 | 13 |
| O95727 | Neurology | 12.2 | 48.8 | 12500 | 200000 | 2.4 | 9 | 8 |
| P01138 | Neurology | 48.8 | 97.7 | 12500 | 25000 | 2.1 | 7 | 4 |
| Q9BZM5 | Neurology | 24.4 | 48.8 | 100000 | 200000 | 3.3 | 9 | 8 |
| Q8TCT1 | Neurology | 3.1 | 6.1 | 781 | 6250 | 2.1 | 8 | 6 |
| Q2MKA7 | Neurology | 3.1 | 3.1 | 1563 | 12500 | 2.7 | 8 | 7 |
| Q9NQ88 | Neurology | 24.4 | 195.3 | 100000 | 400000 | 2.7 | 7 | 24 |
| Q9Y680 | Neurology | 97.7 | 195.3 | 100000 | 800000 | 2.7 | 7 | 10 |
| Q86WV1 | Neurology | 48.8 | 195.3 | 50000 | 200000 | 2.4 | 8 | 8 |
| O14944 | Neurology | 6.1 | 6.1 | 1563 | 6250 | 2.4 | 6 | 6 |
| Q13451 | Neurology | 97.7 | 195.3 | 25000 | 50000 | 2.1 | 9 | 6 |
| Q9Y4K4 | Neurology | 12.2 | 48.8 | 6250 | 50000 | 2.1 | 6 | 10 |
| P16444 | Neurology | | | | | | 8 | 5 |
| O76070 | Neurology | 24.4 | 48.8 | 50000 | 800000 | 3.0 | 9 | 8 |
| Q15814 | Neurology | 781.3 | 781.3 | 400000 | 800000 | 2.7 | 8 | 8 |

| | | | | | | | | |
|---|---|---|---|---|---|---|---|---|
| P08962 | Neurology | 24.4 | 48.8 | 12500 | 50000 | 2.4 | 9 | 6 |
| O15232 | Neurology | 12.2 | 12.2 | 25000 | 200000 | 3.3 | 7 | 6 |
| P48740 | Neurology | 390.6 | 781.3 | 100000 | 400000 | 2.1 | 10 | 6 |
| P38484 | Neurology | 24.4 | 48.8 | 12500 | 100000 | 2.4 | 9 | 4 |
| Q6ISS4 | Neurology | 0.8 | 1.5 | 781 | 6250 | 2.7 | 10 | 6 |
| Q6P1N0 | Neurology | 97.7 | 195.3 | 100000 | 800000 | 2.7 | 9 | 6 |
| Q07011 | Neurology | 0.8 | 1.5 | 3125 | 6250 | 3.3 | 8 | 6 |
| Q8WWN9 | Neurology | 12.2 | 97.7 | 25000 | 50000 | 2.4 | 11 | 6 |
| O15197 | Neurology | 12.2 | 48.8 | 25000 | 50000 | 2.7 | 9 | 6 |
| Q9NPH6 | Neurology | 0.8 | 3.1 | 3125 | 12500 | 3.0 | 9 | 6 |
| P10145 | Neurology | 0.2 | 0.4 | 1563 | 6250 | 3.6 | 7 | 5 |
| Q9P0K1 | Neurology | 97.7 | 195.3 | 50000 | 200000 | 2.4 | 9 | 6 |
| Q9H477 | Neurology | 6.1 | 6.1 | 12500 | 50000 | 3.3 | 7 | 11 |
| Q8N474 | Neurology | 24.4 | 48.8 | 50000 | 200000 | 3.0 | 18 | 20 |
| P40222 | Neurology | 3.1 | 6.1 | 1563 | 12500 | 2.4 | 9 | 8 |
| P02771 | Neurology | 12.2 | 24.4 | 25000 | 100000 | 3.0 | 9 | 4 |
| Q02790 | Neurology | 48.8 | 195.3 | 25000 | 50000 | 2.1 | 8 | 7 |
| Q96GW7 | Neurology | 781.3 | 1562.5 | 200000 | 800000 | 2.1 | 7 | 5 |
| Q9NZQ7 | Neurology | 3.1 | 3.1 | 3125 | 50000 | 3.0 | 9 | 6 |
| P01258 | Neurology | 24.4 | 48.8 | 25000 | 50000 | 2.7 | 8 | 4 |
| Q14108 | Neurology | 3.1 | 6.1 | 6250 | 50000 | 3.0 | 8 | 6 |
| P56279 | Neurology | 97.7 | 195.3 | 50000 | 200000 | 2.4 | 10 | 6 |
| Q9UL46 | Neurology | 48.8 | 195.3 | 25000 | 200000 | 2.1 | 7 | 7 |
| Q9BS40 | Neurology | 390.6 | 390.6 | 200000 | 800000 | 2.7 | 10 | 12 |
| P23588 | Neurology | 390.6 | 390.6 | 100000 | 200000 | 2.4 | 8 | 20 |
| P08134 | Neurology | 24.4 | 48.8 | 50000 | 200000 | 3.0 | 8 | 8 |
| Q8NFP4 | Neurology | 48.8 | 97.7 | 25000 | 200000 | 2.4 | 8 | 4 |
| P22079 | Neurology | 3.1 | 12.2 | 25000 | 200000 | 3.3 | 7 | 8 |
| O43557 | Neurology | 48.8 | 195.3 | 50000 | 200000 | 2.4 | 7 | 6 |
| Q6XZF7 | Neurology | 12.2 | 24.4 | 50000 | 200000 | 3.3 | 9 | 6 |
| P37023 | Neurology | 3.1 | 6.1 | 3125 | 12500 | 2.7 | 9 | 5 |
| O95407 | Neurology | 97.7 | 195.3 | 50000 | 200000 | 2.4 | 10 | 8 |
| Q06830 | Neurology | 97.7 | 195.3 | 25000 | 200000 | 2.1 | 10 | 12 |

| | | | | | | | | |
|---|---|---|---|---|---|---|---|---|
| Q8WV92 | Neurology | 3.1 | 6.1 | 25000 | 50000 | 3.6 | 9 | 8 |
| Q92752 | Neurology | 97.7 | 195.3 | 50000 | 200000 | 2.4 | 9 | 7 |
| Q99426 | Neurology | 24.4 | 97.7 | 100000 | 400000 | 3.0 | 7 | 10 |
| P12644 | Neurology | 6.1 | 48.8 | 12500 | 50000 | 2.4 | 18 | 49 |
| Q9BYC5 | Neurology | | | | | | 7 | 6 |
| O43927 | Neurology | 3.1 | 6.1 | 1563 | 6250 | 2.4 | 6 | 10 |
| P15151 | Neurology | 97.7 | 195.3 | 200000 | 800000 | 3.0 | 9 | 8 |
| Q08345 | Neurology | 12.2 | 12.2 | 50000 | 400000 | 3.6 | 7 | 5 |
| P06733 | Neurology | | | | | | 7 | 11 |
| Q96GP6 | Neurology | 48.8 | 97.7 | 200000 | 400000 | 3.3 | 6 | 7 |
| P14384 | Neurology | 12.2 | 24.4 | 25000 | 200000 | 3.0 | 5 | 5 |
| O14594 | Neurology | 3.1 | 6.1 | 6250 | 50000 | 3.0 | 6 | 8 |
| Q96B86 | Neurology | 24.4 | 48.8 | 100000 | 400000 | 3.3 | 7 | 7 |
| Q6P4E1 | Neurology | 24.4 | 48.8 | 50000 | 100000 | 3.0 | 7 | 5 |
| O14625 | Neurology | 1.5 | 6.1 | 6250 | 12500 | 3.0 | 7 | 9 |
| P35237 | Neurology | 24.4 | 97.7 | 50000 | 400000 | 2.7 | 7 | 7 |
| P52823 | Neurology | | | | | | 7 | 7 |
| Q08708 | Neurology | 3.1 | 6.1 | 6250 | 25000 | 3.0 | 7 | 9 |
| P01178 | Neurology | 6.1 | 24.4 | 25000 | 200000 | 3.0 | 5 | 6 |
| P17405 | Neurology | 97.7 | 195.3 | 400000 | 800000 | 3.3 | 6 | 9 |
| P50225 | Neurology | 97.7 | 195.3 | 100000 | 400000 | 2.7 | 5 | 14 |
| P15311 | Neurology | 390.6 | 781.3 | 100000 | 400000 | 2.1 | 6 | 4 |
| P13611 | Neurology | 1.5 | 3.1 | 6250 | 25000 | 3.3 | 7 | 6 |
| Q8NBJ7 | Neurology | 24.4 | 48.8 | 6250 | 200000 | 2.1 | 6 | 5 |
| Q8NBS9 | Neurology | 3125.0 | 6250.0 | 800000 | 3200000 | 2.1 | 10 | 9 |
| P55291 | Neurology | 1562.5 | 3125.0 | 400000 | 800000 | 2.1 | 7 | 6 |
| P22223 | Neurology | 97.7 | 195.3 | 100000 | 200000 | 2.7 | 9 | 5 |
| O14737 | Neurology | 6.1 | 24.4 | 12500 | 100000 | 2.7 | 8 | 13 |
| P57087 | Neurology | 0.4 | 1.5 | 6250 | 25000 | 3.6 | 6 | 5 |
| P21757 | Neurology | 12.2 | 24.4 | 12500 | 100000 | 2.7 | 6 | 8 |
| Q9NP79 | Neurology | 195.3 | 390.6 | 400000 | 800000 | 3.0 | 9 | 15 |
| P05060 | Neurology | 390.6 | 781.3 | 400000 | 800000 | 2.7 | 7 | 7 |
| P78325 | Neurology | 3.1 | 6.1 | 12500 | 25000 | 3.3 | 5 | 7 |

| | | | | | | | | |
|---|---|---|---|---|---|---|---|---|
| O94907 | Neurology | 12.2 | 24.4 | 25000 | 50000 | 3.0 | 7 | 8 |
| P30533 | Neurology | 6.1 | 12.2 | 6250 | 12500 | 2.7 | 5 | 6 |
| Q9P126 | Neurology | 12.2 | 24.4 | 12500 | 50000 | 2.7 | 11 | 12 |
| Q10589 | Neurology | 6.1 | 12.2 | 25000 | 50000 | 3.3 | 7 | 4 |
| P01236 | Neurology | 24.4 | 97.7 | 100000 | 400000 | 3.0 | 6 | 11 |
| Q92854 | Neurology | 97.7 | 195.3 | 50000 | 400000 | 2.4 | 6 | 7 |
| P28906 | Neurology | 97.7 | 195.3 | 200000 | 400000 | 3.0 | 7 | 12 |
| Q14739 | Neurology | 390.6 | 781.3 | 400000 | 800000 | 2.7 | 6 | 7 |
| P01732 | Neurology | | | | | | 6 | 10 |
| Q99731 | Neurology | | | | | | 5 | 12 |
| Q9H446 | Neurology | 24.4 | 48.8 | 100000 | 200000 | 3.3 | 7 | 11 |
| Q9UNZ2 | Neurology | 48.8 | 97.7 | 100000 | 800000 | 3.0 | 8 | 8 |
| P21217_Q1 | Neurology | 1.5 | 6.1 | 6250 | 50000 | 3.0 | 5 | 7 |
| Q9BRF8 | Neurology | 24.4 | 48.8 | 50000 | 200000 | 3.0 | 5 | 13 |
| P14207 | Neurology | 3.1 | 6.1 | 6250 | 25000 | 3.0 | 6 | 5 |
| P22894 | Neurology | 97.7 | 390.6 | 200000 | 800000 | 2.7 | 5 | 5 |
| Q10588 | Neurology | 48.8 | 97.7 | 50000 | 800000 | 2.7 | 6 | 7 |
| P09104 | Neurology | 3125.0 | 6250.0 | 3200000 | 12800000 | 2.7 | 6 | 13 |
| Q6ZMJ2 | Neurology | 48.8 | 97.7 | 25000 | 100000 | 2.4 | 6 | 6 |
| Q16288 | Neurology | 12.2 | 24.4 | 25000 | 100000 | 3.0 | 6 | 5 |
| Q6NW40 | Neurology | 48.8 | 195.3 | 100000 | 200000 | 2.7 | 6 | 5 |
| Q01469 | Neurology | 24.4 | 97.7 | 100000 | 800000 | 3.0 | 14 | 34 |
| O00214 | Neurology | 12.2 | 48.8 | 25000 | 100000 | 2.7 | 5 | 6 |
| Q9HCK4 | Neurology | 12.2 | 24.4 | 25000 | 200000 | 3.0 | 6 | 6 |
| P13473 | Neurology | 97.7 | 195.3 | 200000 | 800000 | 3.0 | 5 | 6 |
| P12429 | Neurology | 195.3 | 390.6 | 100000 | 200000 | 2.4 | 7 | 8 |
| Q96CD2 | Neurology | 12.2 | 24.4 | 25000 | 200000 | 3.0 | 5 | 6 |
| P28908 | Neurology | 12.2 | 48.8 | 50000 | 200000 | 3.0 | 5 | 6 |
| P48052 | Neurology | 97.7 | 195.3 | 400000 | 800000 | 3.3 | 6 | 6 |
| P25774 | Neurology | 1.5 | 12.2 | 12500 | 50000 | 3.0 | 7 | 6 |
| O94779 | Neurology | 24.4 | 48.8 | 25000 | 100000 | 2.7 | 5 | 6 |
| O75509 | Neurology | 24.4 | 48.8 | 25000 | 100000 | 2.7 | 6 | 6 |
| P04216 | Neurology | 0.4 | 0.8 | 6250 | 25000 | 3.9 | 6 | 6 |

| | | | | | | | | |
|---|---|---|---|---|---|---|---|---|
| P04233 | Neurology | 97.7 | 195.3 | 25000 | 100000 | 2.1 | 5 | 5 |
| Q9NZD4 | Neurology | 6.1 | 6.1 | 6250 | 25000 | 3.0 | 7 | 5 |
| Q14112 | Neurology | 1562.5 | 3125.0 | 800000 | 800000 | 2.4 | 10 | 10 |
| P01215 | Neurology | 0.8 | 1.5 | 6250 | 50000 | 3.6 | 7 | 6 |
| Q96RD9 | Neurology | 6.1 | 12.2 | 25000 | 800000 | 3.3 | 7 | 3 |
| Q9NPH3 | Neurology | 6.1 | 12.2 | 12500 | 50000 | 3.0 | 7 | 3 |
| P16871 | Neurology | 6.1 | 12.2 | 12500 | 200000 | 3.0 | 7 | 15 |
| Q9P232 | Neurology | 6.1 | 24.4 | 12500 | 50000 | 2.7 | 6 | 4 |
| Q8N6Q3 | Neurology | 0.8 | 1.5 | 6250 | 25000 | 3.6 | 8 | 4 |
| P06734 | Neurology | 1.5 | 3.1 | 6250 | 25000 | 3.3 | 7 | 6 |
| P00749 | Neurology | 0.4 | 0.8 | 6250 | 12500 | 3.9 | 5 | 3 |
| Q9Y240 | Neurology | 12.2 | 24.4 | 25000 | 100000 | 3.0 | 8 | 6 |
| Q8TCZ2 | Neurology | 0.2 | 0.8 | 3125 | 6250 | 3.6 | 6 | 7 |
| P20774 | Neurology | 6.1 | 24.4 | 25000 | 50000 | 3.0 | 6 | 4 |
| Q96FE7 | Neurology | 6.1 | 12.2 | 6250 | 25000 | 2.7 | 5 | 4 |
| Q6UXH9 | Neurology | 24.4 | 48.8 | 25000 | 50000 | 2.7 | 7 | 5 |
| Q969Z4 | Neurology | 3.1 | 12.2 | 6250 | 25000 | 2.7 | 8 | 4 |
| Q9NQ38 | Neurology | 97.7 | 195.3 | 25000 | 50000 | 2.1 | 7 | 3 |
| P20333 | Neurology | 3.1 | 12.2 | 12500 | 50000 | 3.0 | 7 | 4 |
| P14174 | Neurology | 6.1 | 12.2 | 3125 | 100000 | 2.4 | 7 | 14 |
| Q9UBW5 | Neurology | 3125.0 | 6250.0 | 400000 | 800000 | 1.8 | 8 | 12 |
| P12081 | Neurology | 48.8 | 195.3 | 100000 | 200000 | 2.7 | 6 | 16 |
| P04118 | Neurology | 24.4 | 48.8 | 50000 | 400000 | 3.0 | 9 | 15 |
| P30740 | Neurology | 97.7 | 390.6 | 400000 | 800000 | 3.0 | 7 | 8 |
| P23381 | Neurology | 48.8 | 97.7 | 200000 | 400000 | 3.3 | 7 | 17 |
| Q86T13 | Neurology | 48.8 | 97.7 | 200000 | 800000 | 3.3 | 5 | 5 |
| Q99972 | Neurology | | | | | | 6 | 8 |
| Q15155 | Neurology | 6.1 | 12.2 | 25000 | 200000 | 3.3 | 5 | 5 |
| Q15818 | Neurology | 6.1 | 12.2 | 25000 | 200000 | 3.3 | 7 | 5 |
| Q9UGT4 | Neurology | 3.1 | 6.1 | 12500 | 50000 | 3.3 | 7 | 6 |
| P05164 | Neurology | 97.7 | 195.3 | 12500 | 25000 | 1.8 | 6 | 6 |
| P55103 | Neurology | 24.4 | 24.4 | 12500 | 50000 | 2.7 | 8 | 7 |
| P22692 | Neurology | 390.6 | 390.6 | 100000 | 400000 | 2.4 | 8 | 5 |

| | | | | | | | | |
|---|---|---|---|---|---|---|---|---|
| P11215 | Neurology | 97.7 | 195.3 | 200000 | 400000 | 3.0 | 8 | 6 |
| P02749 | Neurology | | | | | | 9 | 12 |
| P50895 | Neurology | 24.4 | 48.8 | 6250 | 12500 | 2.1 | 6 | 6 |
| P19440 | Neurology | 6.1 | 24.4 | 50000 | 400000 | 3.3 | 6 | 5 |
| P31948 | Neurology | 24.4 | 97.7 | 25000 | 200000 | 2.4 | 9 | 7 |
| O76076 | Neurology | | | | | | 11 | 13 |
| P30086 | Neurology | 48.8 | 97.7 | 50000 | 400000 | 2.7 | 9 | 10 |
| Q96F46 | Neurology | 3.1 | 6.1 | 12500 | 25000 | 3.3 | 8 | 5 |
| P08254 | Neurology | 6.1 | 12.2 | 6250 | 25000 | 2.7 | 6 | 4 |
| P14778 | Neurology | 0.4 | 0.4 | 6250 | 12500 | 4.2 | 5 | 5 |
| Q04900 | Neurology | 24.4 | 48.8 | 12500 | 25000 | 2.4 | 8 | 5 |
| Q02487 | Neurology | 6.1 | 24.4 | 25000 | 200000 | 3.0 | 8 | 5 |
| O60462 | Neurology | 24.4 | 48.8 | 25000 | 200000 | 2.7 | 10 | 7 |
| O15389 | Neurology | 97.7 | 195.3 | 100000 | 200000 | 2.7 | 8 | 6 |
| P05067 | Neurology | 195.3 | 390.6 | 200000 | 400000 | 2.7 | 5 | 7 |
| Q6YHK3 | Neurology | 48.8 | 97.7 | 200000 | 800000 | 3.3 | 5 | 8 |
| P04179 | Neurology | 390.6 | 1562.5 | 800000 | 800000 | 2.7 | 7 | 4 |
| P00352 | Neurology | 48.8 | 97.7 | 100000 | 200000 | 3.0 | 6 | 6 |
| O43278 | Neurology | 6.1 | 24.4 | 25000 | 400000 | 3.0 | 5 | 3 |
| O76061 | Neurology | 1.5 | 3.1 | 6250 | 25000 | 3.3 | 6 | 5 |
| Q14126 | Neurology | 195.3 | 390.6 | 25000 | 50000 | 1.8 | 7 | 5 |
| Q9Y279 | Neurology | 3.1 | 6.1 | 6250 | 25000 | 3.0 | 6 | 5 |
| P11717 | Neurology | 97.7 | 97.7 | 50000 | 200000 | 2.7 | 7 | 6 |
| P15289 | Neurology | 24.4 | 97.7 | 25000 | 50000 | 2.4 | 5 | 6 |
| Q99727 | Neurology | 3.1 | 6.1 | 3125 | 12500 | 2.7 | 7 | 5 |
| Q02747 | Neurology | 1.5 | 3.1 | 6250 | 25000 | 3.3 | 5 | 6 |
| O15117 | Neurology | 48.8 | 97.7 | 50000 | 200000 | 2.7 | 8 | 8 |
| Q9HCN6 | Neurology | 3.1 | 3.1 | 6250 | 25000 | 3.3 | 7 | 6 |
| Q5T2D2 | Neurology | 3.1 | 6.1 | 6250 | 25000 | 3.0 | 7 | 5 |
| P22105 | Neurology | 97.7 | 390.6 | 50000 | 400000 | 2.1 | 6 | 3 |
| P16284 | Neurology | 6.1 | 12.2 | 50000 | 100000 | 3.6 | 8 | 4 |
| Q9Y624 | Neurology | 0.4 | 0.8 | 3125 | 6250 | 3.6 | 11 | 6 |
| P30043 | Neurology | 97.7 | 390.6 | 200000 | 400000 | 2.7 | 8 | 6 |

| | | | | | | | | |
|---|---|---|---|---|---|---|---|---|
| P14209 | Neurology | 1.5 | 3.1 | 6250 | 25000 | 3.3 | 6 | 3 |
| P01833 | Neurology | 48.8 | 97.7 | 50000 | 200000 | 2.7 | 6 | 4 |
| Q8IWV2 | Neurology | 48.8 | 97.7 | 50000 | 400000 | 2.7 | 6 | 5 |
| P22749 | Neurology | 3.1 | 6.1 | 12500 | 50000 | 3.3 | 8 | 9 |
| P14780 | Neurology | 97.7 | 195.3 | 50000 | 400000 | 2.4 | 7 | 5 |
| P04155 | Neurology | 3.1 | 6.1 | 6250 | 25000 | 3.0 | 7 | 5 |
| P28799 | Neurology | 12.2 | 48.8 | 12500 | 25000 | 2.4 | 7 | 5 |
| P00918 | Neurology | 97.7 | 195.3 | 200000 | 800000 | 3.0 | 8 | 6 |
| Q99497 | Neurology | 6.1 | 12.2 | 3125 | 12500 | 2.4 | 7 | 9 |
| P17538 | Neurology | 1.5 | 3.1 | 12500 | 25000 | 3.6 | 6 | 4 |
| Q9UKK9 | Neurology | 48.8 | 97.7 | 25000 | 50000 | 2.4 | 7 | 10 |
| Q08431 | Neurology | 48.8 | 97.7 | 200000 | 800000 | 3.3 | 8 | 5 |
| Q9UKJ1 | Neurology | 0.8 | 1.5 | 12500 | 25000 | 3.9 | 5 | 4 |
| Q16881 | Neurology | 1.5 | 6.1 | 6250 | 25000 | 3.0 | 8 | 4 |
| Q14696 | Neurology | 1562.5 | 3125.0 | 800000 | 800000 | 2.4 | 8 | 13 |
| P35442 | Neurology | 24.4 | 48.8 | 100000 | 800000 | 3.3 | 6 | 4 |
| Q8TDQ0 | Neurology | 1.5 | 3.1 | 6250 | 25000 | 3.3 | 6 | 4 |
| Q13093 | Neurology | 195.3 | 390.6 | 200000 | 800000 | 2.7 | 7 | 5 |
| P08648 | Neurology | 24.4 | 48.8 | 50000 | 400000 | 3.0 | 10 | 9 |
| P63313 | Neurology | 48.8 | 97.7 | 12500 | 25000 | 2.1 | 7 | 6 |
| P20827 | Neurology | 97.7 | 195.3 | 12500 | 25000 | 1.8 | 8 | 7 |
| P00995 | Neurology | 24.4 | 48.8 | 12500 | 25000 | 2.4 | 8 | 5 |
| Q8N149 | Neurology | 24.4 | 48.8 | 12500 | 50000 | 2.4 | 6 | 4 |
| P07108 | Neurology | 12.2 | 24.4 | 12500 | 25000 | 2.7 | 7 | 7 |
| P19438 | Neurology | 1.5 | 6.1 | 12500 | 25000 | 3.3 | 7 | 3 |
| P23280 | Neurology | 6.1 | 12.2 | 12500 | 25000 | 3.0 | 9 | 7 |
| Q6UXG3 | Neurology | 24.4 | 48.8 | 6250 | 25000 | 2.1 | 7 | 6 |
| O43505 | Neurology | 24.4 | 97.7 | 50000 | 400000 | 2.7 | 7 | 6 |
| O15394 | Neurology | 24.4 | 48.8 | 25000 | 200000 | 2.7 | 7 | 5 |
| Q5SXM8 | Neurology II | 3125.0 | 3125.0 | 200000 | 800000 | 1.8 | 19 | 26 |
| Q96Q89 | Neurology II | 12.2 | 48.8 | 12500 | 100000 | 2.4 | 18 | 12 |
| Q9NQW8 | Neurology II | 781.3 | 781.3 | 50000 | 200000 | 1.8 | 15 | 17 |
| P26441 | Neurology II | 195.3 | 390.6 | 400000 | 800000 | 3.0 | | |

| | | | | | | | | |
|---|---|---|---|---|---|---|---|---|
| Q96GG9 | Neurology II | 1562.5 | 3125.0 | 200000 | 800000 | 1.8 | 18 | |
| P48169 | Neurology II | 3125.0 | 3125.0 | 100000 | 400000 | 1.5 | | |
| P49792 | Neurology II | 3125.0 | 3125.0 | 400000 | 800000 | 2.1 | | |
| Q03721 | Neurology II | 195.3 | 390.6 | 25000 | 100000 | 1.8 | 6 | 10 |
| Q96L15 | Neurology II | 390.6 | 1562.5 | 100000 | 400000 | 1.8 | 13 | |
| Q8N111 | Neurology II | 6.1 | 24.4 | 6250 | 25000 | 2.4 | 15 | 13 |
| Q96P66 | Neurology II | 48.8 | 97.7 | 12500 | 50000 | 2.1 | 9 | 7 |
| P35221 | Neurology II | 6250.0 | 6250.0 | 200000 | 800000 | 1.5 | 7 | 20 |
| Q13936 | Neurology II | 781.3 | 781.3 | 100000 | 800000 | 2.1 | 11 | 36 |
| O95372 | Neurology II | | | | | | 8 | 32 |
| P04234 | Neurology II | 390.6 | 390.6 | 50000 | 400000 | 2.1 | 4 | |
| P25440 | Neurology II | 1562.5 | 3125.0 | 200000 | 400000 | 1.8 | 7 | 6 |
| O95049 | Neurology II | 1562.5 | 3125.0 | 400000 | 800000 | 2.1 | 12 | 20 |
| P08908 | Neurology II | 1562.5 | 1562.5 | 100000 | 800000 | 1.8 | 11 | 20 |
| Q30KQ4 | Neurology II | 195.3 | 195.3 | 12500 | 100000 | 1.8 | 10 | 4 |
| Q13509 | Neurology II | 1562.5 | 1562.5 | 200000 | 800000 | 2.1 | 7 | |
| Q8IVU1 | Neurology II | 1562.5 | 1562.5 | 100000 | 800000 | 1.8 | 13 | |
| Q6P5Q4 | Neurology II | 781.3 | 1562.5 | 50000 | 400000 | 1.5 | 19 | 35 |
| Q9NZA1 | Neurology II | 12.2 | 24.4 | 6250 | 25000 | 2.4 | 8 | 6 |
| P0C7M8 | Neurology II | 195.3 | 195.3 | 50000 | 200000 | 2.4 | 13 | |
| P63172 | Neurology II | 390.6 | 781.3 | 50000 | 100000 | 1.8 | 14 | 19 |
| Q02641 | Neurology II | 781.3 | 781.3 | 50000 | 400000 | 1.8 | 7 | |
| Q14894 | Neurology II | | | | | | 13 | 17 |
| O76039 | Neurology II | 1562.5 | 1562.5 | 100000 | 800000 | 1.8 | 7 | 33 |
| Q7RTW8 | Neurology II | 97.7 | 195.3 | 50000 | 400000 | 2.4 | 10 | 20 |
| Q5U5Z8 | Neurology II | 390.6 | 781.3 | 50000 | 800000 | 1.8 | 21 | 22 |
| P07954 | Neurology II | | | | | | 10 | 27 |
| Q8N8R5 | Neurology II | 1562.5 | 3125.0 | 400000 | 800000 | 2.1 | 14 | 15 |
| O15350 | Neurology II | 1562.5 | 1562.5 | 800000 | 800000 | 2.7 | 4 | 7 |
| Q9ULW2 | Neurology II | 200000.0 | 200000.0 | 6400000 | 12800000 | 1.5 | | |
| Q9Y3E2 | Neurology II | 1562.5 | 3125.0 | 400000 | 800000 | 2.1 | 19 | 11 |
| Q9BZ29 | Neurology II | 195.3 | 390.6 | 100000 | 400000 | 2.4 | 12 | 34 |
| P47874 | Neurology II | 6.1 | 24.4 | 3125 | 25000 | 2.1 | 5 | 10 |

| | | | | | | | | |
|---|---|---|---|---|---|---|---|---|
| Q8IWB1 | Neurology II | 195.3 | 195.3 | 12500 | 800000 | 1.8 | 13 | 15 |
| P23435 | Neurology II | 6250.0 | 6250.0 | 400000 | 1600000 | 1.8 | 14 | |
| Q99700 | Neurology II | 781.3 | 781.3 | 50000 | 400000 | 1.8 | 12 | 28 |
| Q8WTQ1 | Neurology II | 97.7 | 390.6 | 25000 | 100000 | 1.8 | 10 | 21 |
| Q9P2M7 | Neurology II | 781.3 | 1562.5 | 100000 | 200000 | 1.8 | 9 | 8 |
| Q14129 | Neurology II | 390.6 | 781.3 | 50000 | 100000 | 1.8 | 13 | 13 |
| Q13002 | Neurology II | 12.2 | 24.4 | 50000 | 100000 | 3.3 | 12 | 17 |
| Q9H492 | Neurology II | 3125.0 | 3125.0 | 800000 | 800000 | 2.4 | 10 | |
| Q6QNY1 | Neurology II | 195.3 | 390.6 | 25000 | 100000 | 1.8 | 24 | 5 |
| P26378 | Neurology II | 24.4 | 195.3 | 12500 | 50000 | 1.8 | 15 | 24 |
| Q86T26 | Neurology II | 390.6 | 781.3 | 50000 | 800000 | 1.8 | 4 | |
| Q99653 | Neurology II | 12500.0 | 12500.0 | 400000 | 800000 | 1.5 | 13 | 22 |
| Q6QEF8 | Neurology II | 3125.0 | 12500.0 | 800000 | 800000 | 1.8 | 14 | 21 |
| Q12774 | Neurology II | 390.6 | 390.6 | 25000 | 50000 | 1.8 | 16 | 25 |
| Q8WVC0 | Neurology II | 781.3 | 781.3 | 100000 | 800000 | 2.1 | 8 | 13 |
| Q9NXH3 | Neurology II | 781.3 | 781.3 | 50000 | 200000 | 1.8 | 10 | 19 |
| Q13084 | Neurology II | 1562.5 | 1562.5 | 800000 | 800000 | 2.7 | 9 | 6 |
| P30419 | Neurology II | | | | | | 8 | 32 |
| Q8N967 | Neurology II | 24.4 | 48.8 | 12500 | 50000 | 2.4 | 9 | 9 |
| Q8IZJ0 | Neurology II | 3125.0 | 3125.0 | 400000 | 800000 | 2.1 | 6 | 10 |
| Q15042 | Neurology II | 1562.5 | 1562.5 | 50000 | 400000 | 1.5 | 8 | 11 |
| Q96PV0 | Neurology II | 48.8 | 97.7 | 12500 | 100000 | 2.1 | 5 | 13 |
| P05549 | Neurology II | 3125.0 | 6250.0 | 200000 | 800000 | 1.5 | 13 | |
| Q0VDD7 | Neurology II | 25000.0 | 25000.0 | 1600000 | 12800000 | 1.8 | 11 | 27 |
| P28222 | Neurology II | | | | | | 8 | 4 |
| O15382 | Neurology II | 195.3 | 390.6 | 50000 | 400000 | 2.1 | | |
| Q687X5 | Neurology II | 48.8 | 48.8 | 12500 | 100000 | 2.4 | 11 | 17 |
| Q5T871 | Neurology II | | | | | | 4 | |
| Q9UL42 | Neurology II | 12500.0 | 12500.0 | 800000 | 800000 | 1.8 | 7 | 22 |
| O95196 | Neurology II | 390.6 | 390.6 | 50000 | 800000 | 2.1 | 16 | 23 |
| Q9C0A0 | Neurology II | 390.6 | 390.6 | 25000 | 100000 | 1.8 | 7 | 17 |
| O15013 | Neurology II | 390.6 | 1562.5 | 100000 | 200000 | 1.8 | 6 | 22 |
| P12319 | Neurology II | 390.6 | 781.3 | 50000 | 200000 | 1.8 | 12 | 11 |

| | | | | | | | | |
|---|---|---|---|---|---|---|---|---|
| P54284 | Neurology II | 781.3 | 1562.5 | 100000 | 200000 | 1.8 | 11 | 35 |
| Q96I25 | Neurology II | | | | | | 13 | 25 |
| Q01432 | Neurology II | | | | | | 4 | 21 |
| Q9Y6X8 | Neurology II | 390.6 | 390.6 | 50000 | 100000 | 2.1 | 6 | 8 |
| Q8WW22 | Neurology II | | | | | | 8 | 16 |
| P04439 | Neurology II | | | | | | 9 | 9 |
| Q9NWU2 | Neurology II | 781.3 | 781.3 | 25000 | 100000 | 1.5 | 6 | |
| Q5SW96 | Neurology II | 12500.0 | 12500.0 | 800000 | 1600000 | 1.8 | | |
| O15240 | Neurology II | | | | | | 9 | 10 |
| Q92859 | Neurology II | 97.7 | 97.7 | 25000 | 50000 | 2.4 | 8 | 12 |
| Q14627 | Neurology II | 1562.5 | 1562.5 | 800000 | 800000 | 2.7 | 10 | |
| Q9Y3B9 | Neurology II | 781.3 | 781.3 | 100000 | 800000 | 2.1 | | |
| Q96A32 | Neurology II | 3125.0 | 3125.0 | 100000 | 800000 | 1.5 | 14 | 25 |
| Q9NPB3 | Neurology II | 1562.5 | 3125.0 | 200000 | 800000 | 1.8 | 16 | 25 |
| Q9ULU8 | Neurology II | 195.3 | 390.6 | 100000 | 200000 | 2.4 | 10 | 13 |
| Q6GQQ9 | Neurology II | 390.6 | 781.3 | 200000 | 800000 | 2.4 | 8 | 19 |
| Q96HC4 | Neurology II | 390.6 | 781.3 | 100000 | 400000 | 2.1 | 7 | 37 |
| Q9BV20 | Neurology II | | | | | | 9 | 36 |
| Q14184 | Neurology II | 195.3 | 390.6 | 100000 | 200000 | 2.4 | 8 | 15 |
| P43007 | Neurology II | 12500.0 | 25000.0 | 1600000 | 6400000 | 1.8 | 9 | 20 |
| Q68J44 | Neurology II | 390.6 | 390.6 | 50000 | 200000 | 2.1 | 16 | 22 |
| P14649 | Neurology II | 1562.5 | 1562.5 | 100000 | 400000 | 1.8 | 12 | 16 |
| Q96G03 | Neurology II | 390.6 | 781.3 | 200000 | 800000 | 2.4 | 11 | 28 |
| Q9NY46 | Neurology II | 781.3 | 781.3 | 50000 | 800000 | 1.8 | | |
| O75718 | Neurology II | | | | | | 4 | |
| B6A8C7 | Neurology II | 195.3 | 390.6 | 50000 | 800000 | 2.1 | | |
| Q9Y4C0 | Neurology II | 390.6 | 390.6 | 25000 | 800000 | 1.8 | 6 | 31 |
| Q7L311 | Neurology II | 781.3 | 1562.5 | 100000 | 800000 | 1.8 | | |
| Q15735 | Neurology II | 195.3 | 195.3 | 50000 | 100000 | 2.4 | 16 | 29 |
| P48431 | Neurology II | 781.3 | 1562.5 | 100000 | 400000 | 1.8 | 2 | |
| Q6ZVN8 | Neurology II | 781.3 | 1562.5 | 100000 | 200000 | 1.8 | 12 | 10 |
| O43665 | Neurology II | 195.3 | 195.3 | 25000 | 100000 | 2.1 | 8 | 24 |
| Q6Y7W6 | Neurology II | 390.6 | 390.6 | 50000 | 200000 | 2.1 | 9 | 16 |

| | | | | | | | | |
|---|---|---|---|---|---|---|---|---|
| Q8WXW3-4 | Neurology II | 195.3 | 195.3 | 50000 | 200000 | 2.4 | 11 | 31 |
| Q9H4X1 | Neurology II | 1562.5 | 3125.0 | 200000 | 800000 | 1.8 | 8 | 31 |
| Q16625 | Neurology II | 195.3 | 195.3 | 25000 | 100000 | 2.1 | 9 | 12 |
| P78318 | Neurology II | 1562.5 | 1562.5 | 100000 | 400000 | 1.8 | 11 | 21 |
| Q6ZVM7 | Neurology II | | | | | | 12 | 37 |
| Q9H6S1 | Neurology II | 781.3 | 1562.5 | 400000 | 800000 | 2.4 | 10 | 25 |
| P52179 | Neurology II | 195.3 | 781.3 | 50000 | 200000 | 1.8 | 12 | |
| Q99250 | Neurology II | 195.3 | 390.6 | 25000 | 100000 | 1.8 | 9 | 23 |
| O75631 | Neurology II | 781.3 | 781.3 | 50000 | 200000 | 1.8 | | |
| Q96J84 | Neurology II | 1562.5 | 1562.5 | 200000 | 800000 | 2.1 | 9 | 14 |
| Q9UKV0 | Neurology II | | | | | | 8 | 22 |
| Q19T08 | Neurology II | | | | | | 0.1 | |
| Q9ULH4 | Neurology II | | | | | | 21 | |
| O95295 | Neurology II | 781.3 | 781.3 | 100000 | 400000 | 2.1 | 11 | 23 |
| A8MVW0 | Neurology II | 781.3 | 1562.5 | 100000 | 400000 | 1.8 | 9 | 28 |
| Q9BZE9 | Neurology II | 390.6 | 781.3 | 200000 | 800000 | 2.4 | 14 | 22 |
| P61764 | Neurology II | 1562.5 | 3125.0 | 400000 | 800000 | 2.1 | | |
| P68400 | Neurology II | | | | | | 7 | 22 |
| Q13224 | Neurology II | 1562.5 | 6250.0 | 400000 | 800000 | 1.8 | 9 | 12 |
| O75312 | Neurology II | 781.3 | 781.3 | 50000 | 800000 | 1.8 | 13 | 23 |
| P60880 | Neurology II | 97.7 | 390.6 | 50000 | 400000 | 2.1 | 14 | 23 |
| A6NGG8 | Neurology II | 390.6 | 781.3 | 50000 | 100000 | 1.8 | 13 | 8 |
| P05787 | Neurology II | 24.4 | 195.3 | 400000 | 800000 | 3.3 | 13 | 28 |
| Q99856 | Neurology II | | | | | | 12 | |
| Q96PH6 | Neurology II | 3125.0 | 6250.0 | 200000 | 800000 | 1.5 | 3 | 1 |
| P52788 | Neurology II | 3125.0 | 3125.0 | 200000 | 400000 | 1.8 | 9 | 21 |
| Q9Y6U3 | Neurology II | 48.8 | 48.8 | 25000 | 100000 | 2.7 | 12 | 16 |
| Q9Y285 | Neurology II | 97.7 | 781.3 | 100000 | 200000 | 2.1 | 10 | 19 |
| Q9HCM4 | Neurology II | 781.3 | 1562.5 | 100000 | 400000 | 1.8 | 19 | 31 |
| P26715 | Neurology II | 195.3 | 390.6 | 50000 | 100000 | 2.1 | 2 | |
| Q6P9F5 | Neurology II | | | | | | | |
| Q8WXG9 | Neurology II | 781.3 | 781.3 | 100000 | 400000 | 2.1 | 19 | 34 |
| O15020 | Neurology II | 195.3 | 390.6 | 25000 | 100000 | 1.8 | 7 | |

| | | | | | | | | |
|---|---|---|---|---|---|---|---|---|
| P55283 | Neurology II | 390.6 | 781.3 | 50000 | 200000 | 1.8 | 9 | 2 |
| P10523 | Neurology II | 3125.0 | 3125.0 | 200000 | 400000 | 1.8 | | |
| O14503 | Neurology II | 195.3 | 390.6 | 50000 | 400000 | 2.1 | 11 | 26 |
| O43396 | Neurology II | | | | | | 11 | 32 |
| Q8IY33 | Neurology II | 1562.5 | 1562.5 | 50000 | 800000 | 1.5 | 16 | 15 |
| Q2M3V2 | Neurology II | 781.3 | 1562.5 | 200000 | 800000 | 2.1 | 14 | 38 |
| P48775 | Neurology II | 12.2 | 48.8 | 12500 | 50000 | 2.4 | 12 | 16 |
| P61366 | Neurology II | 3125.0 | 3125.0 | 100000 | 200000 | 1.5 | 11 | 21 |
| Q10571 | Neurology II | 781.3 | 781.3 | 50000 | 800000 | 1.8 | 11 | 24 |
| Q8N2Q7 | Neurology II | 1562.5 | 1562.5 | 100000 | 800000 | 1.8 | 13 | 16 |
| Q12809 | Neurology II | 1562.5 | 1562.5 | 50000 | 400000 | 1.5 | 6 | 7 |
| O15212 | Neurology II | 3125.0 | 6250.0 | 800000 | 800000 | 2.1 | 18 | 18 |
| P61266 | Neurology II | 390.6 | 1562.5 | 200000 | 800000 | 2.1 | 13 | 10 |
| Q86UP6 | Neurology II | 6.1 | 24.4 | 6250 | 50000 | 2.4 | 8 | 33 |
| P0DKB5 | Neurology II | 25000.0 | 25000.0 | 1600000 | 6400000 | 1.8 | 16 | 34 |
| P11229 | Neurology II | 1562.5 | 3125.0 | 200000 | 800000 | 1.8 | 12 | 5 |
| Q5VT99 | Neurology II | 1562.5 | 3125.0 | 400000 | 800000 | 2.1 | 9 | 10 |
| Q16650 | Neurology II | 1562.5 | 1562.5 | 200000 | 800000 | 2.1 | 6 | |
| Q6ZUT3 | Neurology II | 1562.5 | 1562.5 | 50000 | 800000 | 1.5 | 7 | 3 |
| Q6UWJ8 | Neurology II | 390.6 | 781.3 | 50000 | 200000 | 1.8 | | |
| P63211 | Neurology II | 390.6 | 390.6 | 25000 | 100000 | 1.8 | 11 | 14 |
| P20823 | Neurology II | 6250.0 | 6250.0 | 400000 | 800000 | 1.8 | 9 | 8 |
| Q16401 | Neurology II | 390.6 | 390.6 | 100000 | 800000 | 2.4 | 11 | 13 |
| O75121 | Neurology II | | | | | | 6 | 17 |
| O15234 | Neurology II | | | | | | 10 | 21 |
| Q86UW9 | Neurology II | 1562.5 | 3125.0 | 200000 | 800000 | 1.8 | 13 | 9 |
| Q86WK6 | Neurology II | 781.3 | 781.3 | 25000 | 800000 | 1.5 | 11 | 21 |
| O15083 | Neurology II | 97.7 | 195.3 | 100000 | 200000 | 2.7 | 16 | 23 |
| Q53GD3 | Neurology II | 195.3 | 390.6 | 100000 | 200000 | 2.4 | 5 | |
| Q8NFZ4 | Neurology II | 3125.0 | 3125.0 | 200000 | 800000 | 1.8 | 10 | |
| P54105 | Neurology II | 1562.5 | 1562.5 | 100000 | 800000 | 1.8 | 10 | 23 |
| Q93015 | Neurology II | 781.3 | 3125.0 | 200000 | 400000 | 1.8 | 11 | 30 |
| Q9UHL0 | Neurology II | | | | | | | |

| | | | | | | | | |
|---|---|---|---|---|---|---|---|---|
| Q8N6Q1 | Neurology II | 390.6 | 1562.5 | 100000 | 200000 | 1.8 | 14 | 19 |
| Q16520 | Neurology II | 48.8 | 97.7 | 25000 | 50000 | 2.4 | 11 | 9 |
| Q96KJ4 | Neurology II | 781.3 | 1562.5 | 100000 | 800000 | 1.8 | 11 | 16 |
| Q9UGI9 | Neurology II | 12500.0 | 12500.0 | 400000 | 1600000 | 1.5 | | |
| Q53EL9 | Neurology II | 781.3 | 781.3 | 50000 | 200000 | 1.8 | 15 | 21 |
| P04062 | Neurology II | 50000.0 | 50000.0 | 1600000 | 6400000 | 1.5 | | |
| Q7L0J3 | Neurology II | 195.3 | 390.6 | 100000 | 400000 | 2.4 | 19 | 29 |
| P01270 | Neurology II | | | | | | | |
| Q8IY31 | Neurology II | 781.3 | 781.3 | 25000 | 100000 | 1.5 | 12 | 13 |
| P10301 | Neurology II | 390.6 | 390.6 | 100000 | 200000 | 2.4 | 16 | 30 |
| P98164 | Neurology II | 1562.5 | 1562.5 | 100000 | 800000 | 1.8 | 23 | 34 |
| Q92834 | Neurology II | 781.3 | 781.3 | 50000 | 200000 | 1.8 | 15 | 16 |
| O14967 | Neurology II | 6.1 | 48.8 | 25000 | 50000 | 2.7 | 16 | 22 |
| Q9H461 | Neurology II | | | | | | 22 | 13 |
| O43474 | Neurology II | 781.3 | 1562.5 | 200000 | 800000 | 2.1 | 12 | 14 |
| Q9NY72 | Neurology II | 6250.0 | 6250.0 | 800000 | 800000 | 2.1 | | |
| P43220 | Neurology II | | | | | | | |
| P41732 | Neurology II | | | | | | 11 | |
| Q17R60 | Neurology II | 781.3 | 1562.5 | 100000 | 400000 | 1.8 | 10 | 10 |
| Q9UKW4 | Neurology II | 97.7 | 195.3 | 25000 | 100000 | 2.1 | 7 | 19 |
| Q9UJC5 | Neurology II | 6250.0 | 12500.0 | 800000 | 3200000 | 1.8 | 13 | 28 |
| Q96BJ3 | Neurology II | | | | | | | |
| O60218 | Neurology II | 6250.0 | 6250.0 | 200000 | 800000 | 1.5 | 16 | |
| P52799 | Neurology II | 390.6 | 781.3 | 100000 | 800000 | 2.1 | 22 | 17 |
| P43146 | Neurology II | | | | | | 11 | 11 |
| P49069 | Neurology II | 781.3 | 1562.5 | 100000 | 200000 | 1.8 | 12 | 26 |
| P09683 | Neurology II | | | | | | 9 | 26 |
| Q7Z3D4 | Neurology II | 97.7 | 195.3 | 100000 | 400000 | 2.7 | 14 | 39 |
| P24386 | Neurology II | 195.3 | 390.6 | 25000 | 100000 | 1.8 | | |
| Q96QH8 | Neurology II | 12.2 | 24.4 | 6250 | 25000 | 2.4 | 9 | 11 |
| Q9UM54 | Neurology II | | | | | | 8 | 23 |
| P08651 | Neurology II | 3125.0 | 3125.0 | 800000 | 800000 | 2.4 | 10 | 19 |
| Q9HCY8 | Neurology II | 195.3 | 390.6 | 50000 | 400000 | 2.1 | 10 | 17 |

| | | | | | | | | |
|---|---|---|---|---|---|---|---|---|
| O60662 | Neurology II | 390.6 | 390.6 | 200000 | 400000 | 2.7 | 14 | 20 |
| Q96JB5 | Neurology II | | | | | | 12 | 16 |
| A6NFN3 | Neurology II | 24.4 | 48.8 | 6250 | 100000 | 2.1 | 9 | 10 |
| Q9P2M1 | Neurology II | | | | | | 10 | 11 |
| Q9HBL6 | Neurology II | 3125.0 | 3125.0 | 100000 | 200000 | 1.5 | 9 | 20 |
| P10746 | Neurology II | 195.3 | 390.6 | 25000 | 800000 | 1.8 | | |
| Q2UY09 | Neurology II | 781.3 | 781.3 | 100000 | 400000 | 2.1 | 13 | 20 |
| Q9NPD7 | Neurology II | | | | | | 6 | |
| Q9BY14 | Neurology II | 12.2 | 24.4 | 6250 | 25000 | 2.4 | 12 | 19 |
| Q9BZJ3 | Neurology II | 195.3 | 390.6 | 50000 | 200000 | 2.1 | 12 | 14 |
| P48436 | Neurology II | 781.3 | 1562.5 | 100000 | 400000 | 1.8 | 16 | |
| Q9NUG6 | Neurology II | 3125.0 | 3125.0 | 100000 | 800000 | 1.5 | 14 | 19 |
| P09455 | Neurology II | 390.6 | 781.3 | 50000 | 800000 | 1.8 | 12 | 20 |
| Q0P6D2 | Neurology II | 3125.0 | 3125.0 | 200000 | 800000 | 1.8 | 10 | 22 |
| Q8WWM7 | Neurology II | 3125.0 | 3125.0 | 400000 | 800000 | 2.1 | 7 | 11 |
| Q07617 | Neurology II | | | | | | 11 | 15 |
| Q9BX66 | Neurology II | 1562.5 | 1562.5 | 50000 | 200000 | 1.5 | 6 | 17 |
| Q15398 | Neurology II | 195.3 | 195.3 | 50000 | 400000 | 2.4 | 6 | 23 |
| P35556 | Neurology II | | | | | | | |
| Q8IWP9 | Neurology II | | | | | | | |
| Q9UNY4 | Neurology II | 390.6 | 390.6 | 25000 | 100000 | 1.8 | | |
| Q9UH03 | Neurology II | 781.3 | 1562.5 | 800000 | 800000 | 2.7 | 7 | 18 |
| P53367 | Neurology II | 390.6 | 1562.5 | 400000 | 800000 | 2.4 | 13 | 29 |
| P60763 | Neurology II | | | | | | 9 | 11 |
| P63010 | Neurology II | | | | | | 12 | 20 |
| O14578 | Neurology II | 48.8 | 48.8 | 25000 | 100000 | 2.7 | 15 | 30 |
| O60939 | Neurology II | 781.3 | 1562.5 | 100000 | 800000 | 1.8 | | |
| P20916 | Neurology II | | | | | | 19 | 22 |
| Q9BX10 | Neurology II | 781.3 | 781.3 | 50000 | 200000 | 1.8 | 12 | 24 |
| Q86TM3 | Neurology II | 390.6 | 781.3 | 100000 | 800000 | 2.1 | 12 | |
| Q15102 | Neurology II | 6250.0 | 6250.0 | 800000 | 800000 | 2.1 | 10 | |
| P54277 | Neurology II | 48.8 | 97.7 | 12500 | 50000 | 2.1 | 21 | 24 |
| P50897 | Neurology II | 3125.0 | 3125.0 | 200000 | 800000 | 1.8 | 9 | 26 |

| | | | | | | | | |
|---|---|---|---|---|---|---|---|---|
| Q8TC05 | Neurology II | 1562.5 | 1562.5 | 100000 | 200000 | 1.8 | 11 | 19 |
| O95202 | Neurology II | 781.3 | 781.3 | 50000 | 200000 | 1.8 | 11 | 22 |
| Q9UQ16 | Neurology II | 390.6 | 390.6 | 100000 | 800000 | 2.4 | 9 | 22 |
| Q9H6Q3 | Neurology II | 3125.0 | 6250.0 | 800000 | 800000 | 2.1 | 10 | 35 |
| Q15714 | Neurology II | 390.6 | 390.6 | 25000 | 100000 | 1.8 | 13 | 19 |
| Q8IVM0 | Neurology II | 781.3 | 1562.5 | 400000 | 800000 | 2.4 | 10 | 13 |
| Q86YD3 | Neurology II | 97.7 | 97.7 | 25000 | 100000 | 2.4 | 9 | 11 |
| P29536 | Neurology II | 24.4 | 48.8 | 12500 | 50000 | 2.4 | 8 | 23 |
| O95954 | Neurology II | 781.3 | 3125.0 | 100000 | 800000 | 1.5 | 5 | 19 |
| Q5T848 | Neurology II | 781.3 | 781.3 | 50000 | 200000 | 1.8 | 9 | 9 |
| B2RUY7 | Neurology II | 48.8 | 97.7 | 12500 | 50000 | 2.1 | 9 | 16 |
| Q14151 | Neurology II | 781.3 | 1562.5 | 100000 | 400000 | 1.8 | 9 | 10 |
| Q92888 | Neurology II | 12500.0 | 12500.0 | 6400000 | 12800000 | 2.7 | 6 | 23 |
| Q96A00 | Neurology II | | | | | | 11 | 32 |
| Q15366 | Neurology II | 25000.0 | 25000.0 | 800000 | 3200000 | 1.5 | 7 | 23 |
| O60869 | Neurology II | | | | | | 13 | 22 |
| O75167 | Neurology II | 781.3 | 781.3 | 100000 | 800000 | 2.1 | | |
| O95467 | Neurology II | | | | | | 7 | 25 |
| P22102 | Neurology II | 781.3 | 1562.5 | 50000 | 200000 | 1.5 | 5 | 23 |
| Q07021 | Neurology II | 48.8 | 195.3 | 50000 | 100000 | 2.4 | 8 | 12 |
| Q08378 | Neurology II | 781.3 | 781.3 | 400000 | 800000 | 2.7 | 10 | 17 |
| O75792 | Neurology II | 1562.5 | 3125.0 | 400000 | 800000 | 2.1 | 9 | 21 |
| O43432 | Neurology II | 1562.5 | 6250.0 | 400000 | 800000 | 1.8 | 11 | 21 |
| Q9BUJ2 | Neurology II | 97.7 | 195.3 | 25000 | 50000 | 2.1 | 7 | 15 |
| Q96PE7 | Neurology II | 6250.0 | 6250.0 | 400000 | 800000 | 1.8 | 11 | 12 |
| Q6UWW0 | Neurology II | 97.7 | 390.6 | 100000 | 400000 | 2.4 | 8 | 12 |
| P51687 | Neurology II | 97.7 | 390.6 | 50000 | 100000 | 2.1 | 9 | 18 |
| Q01826 | Neurology II | 195.3 | 390.6 | 100000 | 400000 | 2.4 | 7 | 7 |
| P04808 | Neurology II | 1562.5 | 6250.0 | 400000 | 800000 | 1.8 | 5 | 26 |
| Q8WWV3 | Neurology II | 781.3 | 781.3 | 200000 | 800000 | 2.4 | 8 | 35 |
| Q9H6H4 | Neurology II | 195.3 | 390.6 | 100000 | 800000 | 2.4 | 6 | 9 |
| O60469 | Neurology II | 195.3 | 195.3 | 50000 | 200000 | 2.4 | 9 | 7 |
| O75592 | Neurology II | 390.6 | 390.6 | 100000 | 800000 | 2.4 | 9 | 20 |

| | | | | | | | | |
|---|---|---|---|---|---|---|---|---|
| Q9H777 | Neurology II | 390.6 | 390.6 | 50000 | 400000 | 2.1 | 9 | 25 |
| Q14197 | Neurology II | 781.3 | 1562.5 | 50000 | 800000 | 1.5 | 12 | 22 |
| Q5JSP0 | Neurology II | 97.7 | 195.3 | 25000 | 100000 | 2.1 | 15 | 30 |
| O43776 | Neurology II | | | | | | 11 | 10 |
| P31751 | Neurology II | 6250.0 | 12500.0 | 800000 | 800000 | 1.8 | | |
| P29377 | Neurology II | 97.7 | 195.3 | 25000 | 200000 | 2.1 | 10 | 14 |
| P14902 | Neurology II | 195.3 | 390.6 | 200000 | 400000 | 2.7 | 14 | 12 |
| Q99460 | Neurology II | | | | | | 15 | 26 |
| Q9BXI9 | Neurology II | 1562.5 | 1562.5 | 200000 | 800000 | 2.1 | 14 | 30 |
| Q5T5Y3 | Neurology II | 97.7 | 195.3 | 25000 | 100000 | 2.1 | 11 | 29 |
| Q6PKH6 | Neurology II | 781.3 | 781.3 | 200000 | 800000 | 2.4 | 6 | 11 |
| Q13277 | Neurology II | 1562.5 | 1562.5 | 200000 | 800000 | 2.1 | 7 | 5 |
| P21579 | Neurology II | 195.3 | 195.3 | 50000 | 800000 | 2.4 | 7 | 10 |
| Q53GL0 | Neurology II | 1562.5 | 6250.0 | 400000 | 800000 | 1.8 | 8 | 16 |
| O60890 | Neurology II | 3125.0 | 3125.0 | 200000 | 800000 | 1.8 | 9 | 21 |
| O15269 | Neurology II | 1562.5 | 3125.0 | 800000 | 800000 | 2.4 | 7 | 12 |
| Q9H1P3 | Neurology II | 781.3 | 1562.5 | 200000 | 800000 | 2.1 | 5 | 7 |
| Q96RU2 | Neurology II | 195.3 | 390.6 | 100000 | 400000 | 2.4 | 8 | 8 |
| P01350 | Neurology II | | | | | | 6 | 30 |
| P78352 | Neurology II | 195.3 | 390.6 | 25000 | 100000 | 1.8 | 8 | 11 |
| P26718 | Neurology II | 48.8 | 97.7 | 25000 | 50000 | 2.4 | 10 | 13 |
| O77932 | Neurology II | 97.7 | 97.7 | 6250 | 25000 | 1.8 | 6 | 16 |
| Q92599 | Neurology II | 195.3 | 390.6 | 200000 | 800000 | 2.7 | 5 | 9 |
| P13995 | Neurology II | 195.3 | 195.3 | 12500 | 50000 | 1.8 | 6 | 15 |
| Q8N163 | Neurology II | 781.3 | 1562.5 | 400000 | 800000 | 2.4 | 9 | 16 |
| Q8IWY9 | Neurology II | 50000.0 | 50000.0 | 3200000 | 12800000 | 1.8 | | |
| Q86VP1 | Neurology II | 24.4 | 24.4 | 12500 | 25000 | 2.7 | 6 | 11 |
| Q9H251 | Neurology II | 1562.5 | 3125.0 | 100000 | 800000 | 1.5 | 8 | 13 |
| Q14149 | Neurology II | 1562.5 | 3125.0 | 800000 | 800000 | 2.4 | 5 | 5 |
| P43320 | Neurology II | 97.7 | 195.3 | 100000 | 400000 | 2.7 | 6 | 14 |
| P51649 | Neurology II | 3125.0 | 6250.0 | 200000 | 400000 | 1.5 | 11 | 15 |
| Q9NWM8 | Neurology II | 781.3 | 781.3 | 25000 | 100000 | 1.5 | 9 | 33 |
| Q14160 | Neurology II | 390.6 | 781.3 | 50000 | 400000 | 1.8 | 10 | 24 |

| | | | | | | | | |
|---|---|---|---|---|---|---|---|---|
| Q9BUP0 | Neurology II | 195.3 | 390.6 | 50000 | 200000 | 2.1 | 9 | 15 |
| O94830 | Neurology II | 195.3 | 390.6 | 25000 | 100000 | 1.8 | 7 | 16 |
| Q8IWQ3 | Neurology II | 3125.0 | 3125.0 | 100000 | 400000 | 1.5 | 8 | 21 |
| Q9NPG4 | Neurology II | | | | | | 6 | 8 |
| Q6UXV0 | Neurology II | 12.2 | 24.4 | 12500 | 50000 | 2.7 | 7 | 17 |
| Q4ZHG4 | Neurology II | | | | | | 8 | 17 |
| Q13938 | Neurology II | 48.8 | 48.8 | 12500 | 200000 | 2.4 | 7 | 9 |
| P47813 | Neurology II | | | | | | 12 | 25 |
| Q9UKY0 | Neurology II | 48.8 | 48.8 | 12500 | 25000 | 2.4 | 3 | 13 |
| P54315 | Neurology II | | | | | | 9 | 22 |
| Q15256 | Neurology II | 48.8 | 48.8 | 25000 | 100000 | 2.7 | 6 | 10 |
| O14530 | Neurology II | 6250.0 | 6250.0 | 200000 | 800000 | 1.5 | 5 | 17 |
| Q15025 | Neurology II | 97.7 | 195.3 | 25000 | 200000 | 2.1 | 4 | 12 |
| P13861 | Neurology II | 781.3 | 1562.5 | 400000 | 800000 | 2.4 | 4 | 13 |
| P10092 | Neurology II | 195.3 | 195.3 | 200000 | 800000 | 3.0 | 8 | 10 |
| Q9ULA0 | Neurology II | 390.6 | 390.6 | 50000 | 400000 | 2.1 | 3 | 19 |
| P59780 | Neurology II | 25000.0 | 25000.0 | 3200000 | 12800000 | 2.1 | 7 | 11 |
| Q8WYQ3 | Neurology II | 195.3 | 195.3 | 6250 | 25000 | 1.5 | 10 | 19 |
| Q9NPE2 | Neurology II | 781.3 | 781.3 | 200000 | 400000 | 2.4 | 6 | 7 |
| Q6UX71 | Neurology II | 781.3 | 781.3 | 50000 | 400000 | 1.8 | 10 | 19 |
| Q6XQN6 | Neurology II | | | | | | 6 | 9 |
| P46937 | Neurology II | 781.3 | 1562.5 | 50000 | 100000 | 1.5 | 6 | 9 |
| Q9BXI3 | Neurology II | 97.7 | 97.7 | 6250 | 25000 | 1.8 | 6 | 11 |
| Q9P2X3 | Neurology II | 390.6 | 781.3 | 100000 | 400000 | 2.1 | 3 | 11 |
| Q99704 | Neurology II | 6250.0 | 6250.0 | 200000 | 800000 | 1.5 | | |
| Q6P1J6 | Neurology II | 97.7 | 97.7 | 12500 | 100000 | 2.1 | 9 | 21 |
| O60235 | Neurology II | 12.2 | 24.4 | 6250 | 25000 | 2.4 | 8 | 8 |
| Q03252 | Neurology II | 1562.5 | 1562.5 | 800000 | 800000 | 2.7 | 6 | 13 |
| Q8TAT2 | Neurology II | 390.6 | 390.6 | 12500 | 50000 | 1.5 | 7 | 8 |
| Q5F1R6 | Neurology II | 390.6 | 390.6 | 25000 | 100000 | 1.8 | 11 | 17 |
| Q03014 | Neurology II | 6.1 | 6.1 | 6250 | 25000 | 3.0 | 7 | 26 |
| Q6QNY0 | Neurology II | 781.3 | 1562.5 | 50000 | 200000 | 1.5 | 7 | 17 |
| Q9BPX1 | Neurology II | 390.6 | 781.3 | 200000 | 800000 | 2.4 | 6 | 22 |

| | | | | | | | | |
|---|---|---|---|---|---|---|---|---|
| P32455 | Neurology II | 3125.0 | 3125.0 | 3200000 | 6400000 | 3.0 | 14 | 25 |
| Q9Y3C0 | Neurology II | 195.3 | 390.6 | 25000 | 200000 | 1.8 | 5 | 17 |
| P62760 | Neurology II | 195.3 | 390.6 | 25000 | 100000 | 1.8 | 7 | 17 |
| Q2L4Q9 | Neurology II | 3.1 | 6.1 | 12500 | 25000 | 3.3 | 7 | 10 |
| P55210 | Neurology II | | | | | | 7 | 19 |
| P51460 | Neurology II | | | | | | | |
| Q5VV43 | Neurology II | 195.3 | 195.3 | 25000 | 100000 | 2.1 | 7 | 11 |
| A0FGR8 | Neurology II | 6250.0 | 12500.0 | 800000 | 800000 | 1.8 | 9 | 18 |
| P11137 | Neurology II | 25000.0 | 50000.0 | 1600000 | 6400000 | 1.5 | 5 | 10 |
| Q9BRJ6 | Neurology II | | | | | | 5 | 7 |
| Q7Z4V5 | Neurology II | | | | | | 10 | 14 |
| P08579 | Neurology II | 390.6 | 781.3 | 50000 | 400000 | 1.8 | 9 | 13 |
| Q9BQT9 | Neurology II | 3125.0 | 6250.0 | 200000 | 800000 | 1.5 | 6 | 15 |
| Q9UBC9 | Neurology II | | | | | | 12 | 18 |
| P0CG30 | Neurology II | 48.8 | 48.8 | 12500 | 100000 | 2.4 | 5 | 39 |
| Q8IXS6 | Neurology II | 390.6 | 781.3 | 50000 | 400000 | 1.8 | 5 | 9 |
| O75146 | Neurology II | | | | | | 7 | 10 |
| Q9NRY6 | Neurology II | 1562.5 | 1562.5 | 200000 | 800000 | 2.1 | 7 | 16 |
| Q96CN9 | Neurology II | 97.7 | 195.3 | 50000 | 200000 | 2.4 | 4 | 36 |
| Q9NXV2 | Neurology II | 97.7 | 195.3 | 50000 | 100000 | 2.4 | 8 | 11 |
| P07320 | Neurology II | 3.1 | 24.4 | 3125 | 25000 | 2.1 | 5 | 15 |
| P54252 | Neurology II | 1562.5 | 3125.0 | 100000 | 800000 | 1.5 | 6 | 13 |
| Q8IXM2 | Neurology II | 195.3 | 390.6 | 50000 | 200000 | 2.1 | 7 | 22 |
| A8MVW5 | Neurology II | 390.6 | 390.6 | 50000 | 400000 | 2.1 | 7 | 26 |
| O60220 | Neurology II | 390.6 | 781.3 | 50000 | 100000 | 1.8 | 10 | 27 |
| Q8TF65 | Neurology II | 195.3 | 195.3 | 25000 | 100000 | 2.1 | 7 | 8 |
| P53814 | Neurology II | 1562.5 | 1562.5 | 50000 | 400000 | 1.5 | 5 | 12 |
| Q07817 | Neurology II | | | | | | | |
| Q00722 | Neurology II | 390.6 | 781.3 | 50000 | 200000 | 1.8 | | |
| P40313 | Neurology II | 97.7 | 97.7 | 12500 | 25000 | 2.1 | 7 | 22 |
| Q8N4C8 | Neurology II | | | | | | | |
| P54819 | Neurology II | | | | | | 11 | 37 |
| Q9NR46 | Neurology II | | | | | | 6 | 16 |

| | | | | | | | | |
|---|---|---|---|---|---|---|---|---|
| P09110 | Oncology | 1562.5 | 3125.0 | 400000 | 800000 | 2.1 | 14 | 20 |
| O14713 | Oncology | | | | | | 11 | 14 |
| Q13145 | Oncology | | | | | | 7 | 33 |
| Q9NX58 | Oncology | | | | | | 9 | 32 |
| O95498 | Oncology | 781.3 | 781.3 | 200000 | 800000 | 2.4 | 6 | 10 |
| Q07954 | Oncology | 12.2 | 24.4 | 12500 | 50000 | 2.7 | 9 | 11 |
| Q9BTE6 | Oncology | 48.8 | 195.3 | 200000 | 400000 | 3.0 | 8 | 16 |
| Q6UWW8 | Oncology | 195.3 | 390.6 | 200000 | 400000 | 2.7 | 9 | 53 |
| P01229 | Oncology | 6250.0 | 12500.0 | 800000 | 1600000 | 1.8 | 10 | 16 |
| P05937 | Oncology | 0.8 | 1.5 | 6250 | 25000 | 3.6 | 8 | 13 |
| P08069 | Oncology | 97.7 | 195.3 | 100000 | 800000 | 2.7 | 6 | 12 |
| P00519 | Oncology | 12.2 | 24.4 | 50000 | 200000 | 3.3 | 9 | 11 |
| Q96I82 | Oncology | 97.7 | 195.3 | 100000 | 200000 | 2.7 | 7 | 11 |
| Q9BS26 | Oncology | 97.7 | 195.3 | 100000 | 400000 | 2.7 | 7 | 10 |
| Q14241 | Oncology | 3.1 | 6.1 | 12500 | 50000 | 3.3 | 7 | 10 |
| Q9HAV7 | Oncology | 12.2 | 24.4 | 50000 | 200000 | 3.3 | 7 | 13 |
| Q9BSL1 | Oncology | 48.8 | 195.3 | 25000 | 800000 | 2.1 | 10 | 11 |
| Q9C0C4 | Oncology | 24.4 | 48.8 | 50000 | 800000 | 3.0 | 10 | 8 |
| O00592 | Oncology | 97.7 | 195.3 | 12500 | 100000 | 1.8 | 8 | 8 |
| Q9UK85 | Oncology | 48.8 | 97.7 | 50000 | 800000 | 2.7 | 8 | 11 |
| Q7Z5R6 | Oncology | 195.3 | 390.6 | 100000 | 400000 | 2.4 | 10 | 10 |
| P30041 | Oncology | | | | | | 6 | 15 |
| P82980 | Oncology | 97.7 | 195.3 | 12500 | 50000 | 1.8 | 8 | 11 |
| P47992 | Oncology | | | | | | 4 | 7 |
| Q9NSA1 | Oncology | 12.2 | 24.4 | 25000 | 200000 | 3.0 | 4 | 5 |
| Q9NTU7 | Oncology | 97.7 | 195.3 | 200000 | 800000 | 3.0 | 4 | 27 |
| Q14213_Q8 | Oncology | 97.7 | 390.6 | 400000 | 800000 | 3.0 | 3 | 8 |
| P13726 | Oncology | 0.8 | 1.5 | 6250 | 25000 | 3.6 | 7 | 4 |
| P06756 | Oncology | 48.8 | 97.7 | 12500 | 100000 | 2.1 | 3 | 4 |
| P61218 | Oncology | 12.2 | 24.4 | 6250 | 12500 | 2.4 | 10 | 11 |
| Q96NA2 | Oncology | 24.4 | 48.8 | 25000 | 50000 | 2.7 | 5 | 4 |
| P50579 | Oncology | 781.3 | 1562.5 | 200000 | 800000 | 2.1 | 4 | 8 |
| Q9UBG3 | Oncology | 1.5 | 3.1 | 6250 | 12500 | 3.3 | 3 | 4 |

| | | | | | | | | |
|---|---|---|---|---|---|---|---|---|
| Q14790 | Oncology | 0.8 | 1.5 | 6250 | 12500 | 3.6 | 6 | 13 |
| P35637 | Oncology | 100000.0 | 100000.0 | 6400000 | 12800000 | 1.8 | 8 | 16 |
| Q13490 | Oncology | 24.4 | 195.3 | 50000 | 200000 | 2.4 | 11 | 24 |
| Q9UQB8 | Oncology | | | | | | 9 | 11 |
| Q6EIG7 | Oncology | 1.5 | 3.1 | 6250 | 12500 | 3.3 | 10 | 18 |
| P80075 | Oncology | 0.8 | 1.5 | 1563 | 6250 | 3.0 | 4 | 5 |
| O00292 | Oncology | | | | | | 4 | 10 |
| Q9BSG5 | Oncology | 97.7 | 195.3 | 50000 | 200000 | 2.4 | 9 | 14 |
| Q99075 | Oncology | 0.8 | 1.5 | 3125 | 12500 | 3.3 | 4 | 4 |
| Q9Y5W5 | Oncology | 48.8 | 48.8 | 25000 | 400000 | 2.7 | 4 | 6 |
| P42658 | Oncology | 97.7 | 195.3 | 200000 | 800000 | 3.0 | 9 | 13 |
| Q99717 | Oncology | 195.3 | 390.6 | 25000 | 200000 | 1.8 | 8 | 12 |
| O43699 | Oncology | 12.2 | 24.4 | 25000 | 200000 | 3.0 | 3 | 5 |
| Q86SJ6 | Oncology | 12.2 | 24.4 | 6250 | 25000 | 2.4 | 6 | 11 |
| P35318 | Oncology | 781.3 | 1562.5 | 400000 | 800000 | 2.4 | 4 | 8 |
| P35813 | Oncology | 390.6 | 781.3 | 50000 | 200000 | 1.8 | 8 | 14 |
| Q7L5Y9 | Oncology | | | | | | 9 | 42 |
| P01375 | Oncology | 6.1 | 12.2 | 12500 | 100000 | 3.0 | 7 | 20 |
| Q9Y265 | Oncology | 781.3 | 3125.0 | 400000 | 800000 | 2.1 | 11 | 15 |
| P42331 | Oncology | 195.3 | 390.6 | 100000 | 800000 | 2.4 | 10 | 16 |
| P06850 | Oncology | 1562.5 | 3125.0 | 400000 | 800000 | 2.1 | 12 | 27 |
| Q8IUK5 | Oncology | 48.8 | 97.7 | 25000 | 200000 | 2.4 | 12 | 14 |
| Q9BSW2 | Oncology | 24.4 | 195.3 | 25000 | 200000 | 2.1 | 11 | 16 |
| O95388 | Oncology | 12.2 | 24.4 | 6250 | 25000 | 2.4 | 3 | 4 |
| Q2VWP7 | Oncology | 6.1 | 12.2 | 6250 | 100000 | 2.7 | 5 | 6 |
| Q00796 | Oncology | | | | | | 6 | 11 |
| O95786 | Oncology | 1562.5 | 3125.0 | 800000 | 800000 | 2.4 | 8 | 13 |
| Q9UHF1 | Oncology | | | | | | 11 | 18 |
| P14136 | Oncology | 97.7 | 195.3 | 400000 | 800000 | 3.3 | 14 | 23 |
| P31994 | Oncology | | | | | | 12 | 27 |
| P55789 | Oncology | 781.3 | 1562.5 | 100000 | 800000 | 1.8 | 9 | 17 |
| P55273 | Oncology | | | | | | 8 | 23 |
| Q9Y243 | Oncology | 48.8 | 97.7 | 50000 | 200000 | 2.7 | 9 | 24 |

| | | | | | | | | |
|---|---|---|---|---|---|---|---|---|
| P22307 | Oncology | 24.4 | 195.3 | 100000 | 200000 | 2.7 | 12 | 12 |
| P43628 | Oncology | 24.4 | 195.3 | 25000 | 200000 | 2.1 | 9 | 18 |
| P31350 | Oncology | 390.6 | 781.3 | 50000 | 400000 | 1.8 | 11 | 20 |
| P39748 | Oncology | | | | | | 11 | 25 |
| O14964 | Oncology | 3125.0 | 6250.0 | 800000 | 800000 | 2.1 | 11 | 19 |
| Q9NRA1 | Oncology | 195.3 | 781.3 | 100000 | 800000 | 2.1 | 10 | 16 |
| Q05516 | Oncology | 24.4 | 48.8 | 50000 | 200000 | 3.0 | 9 | 18 |
| P48643 | Oncology | | | | | | 9 | 38 |
| P46060 | Oncology | 6250.0 | 6250.0 | 1600000 | 12800000 | 2.4 | 10 | 11 |
| O75569 | Oncology | 3125.0 | 6250.0 | 400000 | 800000 | 1.8 | 11 | 15 |
| Q6UX82 | Oncology | 24.4 | 48.8 | 25000 | 200000 | 2.7 | 10 | 16 |
| Q99683 | Oncology | 6250.0 | 6250.0 | 1600000 | 12800000 | 2.4 | 13 | 21 |
| P01242 | Oncology | 390.6 | 781.3 | 100000 | 200000 | 2.1 | 14 | 19 |
| Q08AG7 | Oncology | 48.8 | 195.3 | 50000 | 100000 | 2.4 | 9 | 27 |
| Q96DU3 | Oncology | 390.6 | 781.3 | 50000 | 800000 | 1.8 | 13 | 13 |
| P43629 | Oncology | 6.1 | 12.2 | 50000 | 100000 | 3.6 | 15 | 35 |
| O43752 | Oncology | 390.6 | 781.3 | 100000 | 800000 | 2.1 | 11 | 13 |
| O60828 | Oncology | 195.3 | 390.6 | 50000 | 200000 | 2.1 | 8 | 22 |
| P35070 | Oncology | 3.1 | 6.1 | 3125 | 12500 | 2.7 | 10 | 20 |
| Q8IWL2 | Oncology | 3125.0 | 3125.0 | 100000 | 400000 | 1.5 | 10 | 29 |
| Q7Z7D3 | Oncology | 24.4 | 48.8 | 50000 | 200000 | 3.0 | 11 | 41 |
| P34130 | Oncology | 6.1 | 12.2 | 6250 | 200000 | 2.7 | 7 | 26 |
| Q9UKR0 | Oncology | 24.4 | 48.8 | 12500 | 100000 | 2.4 | 8 | 19 |
| Q6NXT1 | Oncology | 97.7 | 195.3 | 100000 | 200000 | 2.7 | 9 | 15 |
| P54727 | Oncology | 1562.5 | 3125.0 | 400000 | 800000 | 2.1 | 12 | 16 |
| Q6BAA4 | Oncology | 97.7 | 195.3 | 50000 | 200000 | 2.4 | 11 | 17 |
| Q92982 | Oncology | 97.7 | 195.3 | 100000 | 800000 | 2.7 | 11 | 20 |
| Q8NBZ7 | Oncology | 1562.5 | 3125.0 | 800000 | 800000 | 2.4 | 8 | 22 |
| P41586 | Oncology | 24.4 | 48.8 | 6250 | 800000 | 2.1 | 8 | 9 |
| O75787 | Oncology | | | | | | 12 | 14 |
| Q15797 | Oncology | 48.8 | 195.3 | 400000 | 800000 | 3.3 | 14 | 24 |
| Q96NB1 | Oncology | 24.4 | 48.8 | 100000 | 800000 | 3.3 | 10 | 23 |
| Q07960 | Oncology | 24.4 | 97.7 | 12500 | 50000 | 2.1 | 8 | 15 |

| | | | | | | | | |
|---|---|---|---|---|---|---|---|---|
| P50749 | Oncology | 195.3 | 390.6 | 100000 | 800000 | 2.4 | 10 | 18 |
| Q6PGN9 | Oncology | | | | | | 9 | 16 |
| P06731 | Oncology | | | | | | 15 | 23 |
| Q8IWL1 | Oncology | 195.3 | 781.3 | 100000 | 200000 | 2.1 | 12 | 15 |
| O14662 | Oncology | | | | | | 10 | 20 |
| Q7Z6M1 | Oncology | 1562.5 | 3125.0 | 800000 | 6400000 | 2.4 | 10 | 11 |
| Q9UQQ2 | Oncology | 781.3 | 1562.5 | 100000 | 800000 | 1.8 | 9 | 16 |
| P25786 | Oncology | 6250.0 | 12500.0 | 400000 | 800000 | 1.5 | 15 | 35 |
| Q9H4P4 | Oncology | 312.5 | 625.0 | 320000 | 640000 | 2.7 | 10 | 11 |
| O75493 | Oncology | 1562.5 | 3125.0 | 400000 | 800000 | 2.1 | 11 | 18 |
| Q9NS15 | Oncology | 97.7 | 195.3 | 25000 | 100000 | 2.1 | 11 | 27 |
| A4D1B5 | Oncology | | | | | | 10 | 18 |
| P49788 | Oncology | 97.7 | 781.3 | 100000 | 800000 | 2.1 | 10 | 14 |
| P21810 | Oncology | 24.4 | 48.8 | 25000 | 100000 | 2.7 | 14 | 21 |
| Q7LG56 | Oncology | | | | | | 12 | 28 |
| Q9P0J1 | Oncology | | | | | | 8 | 16 |
| Q9Y5V3 | Oncology | 3.1 | 3.1 | 1563 | 12500 | 2.7 | 9 | 54 |
| Q8N5S9 | Oncology | 97.7 | 195.3 | 25000 | 200000 | 2.1 | 11 | 21 |
| Q7Z434 | Oncology | 390.6 | 781.3 | 100000 | 800000 | 2.1 | 10 | 13 |
| P07332 | Oncology | 6250.0 | 12500.0 | 400000 | 800000 | 1.5 | 5 | |
| O15116 | Oncology | 48.8 | 195.3 | 25000 | 100000 | 2.1 | 9 | 43 |
| P43490 | Oncology | 3125.0 | 3125.0 | 800000 | 1600000 | 2.4 | 13 | 16 |
| O75380 | Oncology | 48.8 | 97.7 | 25000 | 100000 | 2.4 | 11 | 20 |
| O60907 | Oncology | 24.4 | 48.8 | 12500 | 100000 | 2.4 | 11 | 19 |
| Q01543 | Oncology | 1.5 | 3.1 | 1563 | 3125 | 2.7 | 7 | 37 |
| Q9UKS7 | Oncology | 6.1 | 6.1 | 6250 | 25000 | 3.0 | 8 | 21 |
| Q06787 | Oncology | 97.7 | 97.7 | 200000 | 800000 | 3.3 | 8 | 18 |
| P04637 | Oncology | 781.3 | 1562.5 | 400000 | 800000 | 2.4 | 10 | 23 |
| Q8WUX2 | Oncology | 97.7 | 195.3 | 50000 | 200000 | 2.4 | 7 | 19 |
| Q9Y6A5 | Oncology | 24.4 | 195.3 | 25000 | 800000 | 2.1 | 8 | 25 |
| P34949 | Oncology | 195.3 | 390.6 | 200000 | 800000 | 2.7 | 9 | 12 |
| Q8WYN0 | Oncology | 1562.5 | 3125.0 | 800000 | 800000 | 2.4 | 8 | 15 |
| Q96PQ0 | Oncology | 1562.5 | 1562.5 | 200000 | 800000 | 2.1 | 10 | 24 |

| | | | | | | | | |
|---|---|---|---|---|---|---|---|---|
| P15121 | Oncology | 1562.5 | 1562.5 | 400000 | 800000 | 2.4 | 11 | 18 |
| P36888 | Oncology | 48.8 | 97.7 | 25000 | 100000 | 2.4 | 10 | 11 |
| Q9Y662 | Oncology | 390.6 | 781.3 | 400000 | 800000 | 2.7 | 8 | 12 |
| Q8TE58 | Oncology | 195.3 | 781.3 | 200000 | 800000 | 2.4 | 9 | 14 |
| Q9BYE9 | Oncology | 3125.0 | 6250.0 | 800000 | 800000 | 2.1 | 7 | 12 |
| P05231 | Oncology | 0.8 | 1.5 | 3125 | 12500 | 3.3 | 9 | 8 |
| Q7L5N7 | Oncology | | | | | | 8 | 15 |
| P55008 | Oncology | | | | | | 11 | 20 |
| P40198 | Oncology | | | | | | 16 | 31 |
| Q9Y223 | Oncology | 781.3 | 1562.5 | 400000 | 800000 | 2.4 | 8 | 11 |
| Q9Y5L3 | Oncology | 6.1 | 12.2 | 6250 | 25000 | 2.7 | 9 | 27 |
| P05783 | Oncology | 6.1 | 12.2 | 12500 | 100000 | 3.0 | 7 | 11 |
| Q8TD06 | Oncology | 50000.0 | 50000.0 | 6400000 | 12800000 | 2.1 | 11 | 13 |
| Q9Y2Z0 | Oncology | | | | | | 11 | 23 |
| Q9P0V8 | Oncology | 6.1 | 48.8 | 6250 | 25000 | 2.1 | 10 | 18 |
| P51580 | Oncology | 48.8 | 195.3 | 50000 | 200000 | 2.4 | 10 | 14 |
| O43524 | Oncology | 1562.5 | 6250.0 | 200000 | 400000 | 1.5 | 13 | 13 |
| O75695 | Oncology | 3125.0 | 6250.0 | 400000 | 800000 | 1.8 | 8 | 10 |
| O00233 | Oncology | 48.8 | 195.3 | 100000 | 400000 | 2.7 | 9 | 9 |
| Q9GZY6 | Oncology | 781.3 | 1562.5 | 400000 | 800000 | 2.4 | 7 | 16 |
| Q5VIR6 | Oncology | 97.7 | 195.3 | 50000 | 200000 | 2.4 | 8 | 9 |
| Q9UJ71 | Oncology | 48.8 | 48.8 | 50000 | 400000 | 3.0 | 10 | 13 |
| Q86WD7 | Oncology | 6.1 | 12.2 | 50000 | 200000 | 3.6 | 7 | 16 |
| Q15427 | Oncology | 97.7 | 195.3 | 25000 | 100000 | 2.1 | 9 | 14 |
| P10606 | Oncology | 97.7 | 195.3 | 50000 | 100000 | 2.4 | 5 | 14 |
| P51692 | Oncology | 24.4 | 48.8 | 200000 | 400000 | 3.6 | 14 | 20 |
| P0CG37 | Oncology | 97.7 | 195.3 | 100000 | 200000 | 2.7 | 10 | 29 |
| Q9H4A9 | Oncology | 97.7 | 390.6 | 50000 | 400000 | 2.1 | 10 | 11 |
| P08473 | Oncology | 24.4 | 48.8 | 25000 | 100000 | 2.7 | 9 | 9 |
| Q9NUY8 | Oncology | 48.8 | 97.7 | 12500 | 25000 | 2.1 | 10 | 26 |
| P17948 | Oncology | 781.3 | 1562.5 | 200000 | 400000 | 2.1 | 8 | 9 |
| P10747 | Oncology | 195.3 | 390.6 | 100000 | 800000 | 2.4 | 11 | 15 |
| Q16772 | Oncology | 24.4 | 48.8 | 25000 | 100000 | 2.7 | 10 | 13 |

| | | | | | | | | |
|---|---|---|---|---|---|---|---|---|
| Q9BUE0 | Oncology | | | | | | 10 | 14 |
| O00186 | Oncology | | | | | | 8 | 24 |
| Q3B7J2 | Oncology | 100000.0 | 400000.0 | 12800000 | 12800000 | 1.5 | 8 | 16 |
| Q6P2H3 | Oncology | 87.9 | 175.8 | 45000 | 180000 | 2.4 | 9 | 31 |
| O00221 | Oncology | 24.4 | 48.8 | 25000 | 200000 | 2.7 | 10 | 15 |
| Q9BQ51 | Oncology | 24.4 | 97.7 | 6250 | 400000 | 1.8 | 9 | 12 |
| O94760 | Oncology | 97.7 | 390.6 | 100000 | 800000 | 2.4 | 11 | 11 |
| Q9UHD8 | Oncology | | | | | | 7 | 9 |
| P30260 | Oncology | 6.1 | 12.2 | 50000 | 200000 | 3.6 | 10 | 29 |
| Q9Y639 | Oncology | 390.6 | 781.3 | 50000 | 200000 | 1.8 | 12 | 15 |
| O95831 | Oncology | 97.7 | 195.3 | 25000 | 800000 | 2.1 | 11 | 8 |
| Q6UXD5 | Oncology | 48.8 | 195.3 | 100000 | 400000 | 2.7 | 7 | 7 |
| O75054 | Oncology | 6.1 | 12.2 | 25000 | 200000 | 3.3 | 8 | 6 |
| Q9Y570 | Oncology | 390.6 | 781.3 | 400000 | 800000 | 2.7 | 9 | 17 |
| P07947 | Oncology | 24.4 | 48.8 | 12500 | 50000 | 2.4 | 11 | 16 |
| P15848 | Oncology | 48.8 | 195.3 | 100000 | 800000 | 2.7 | 8 | 12 |
| Q11201 | Oncology | 3125.0 | 6250.0 | 800000 | 800000 | 2.1 | 9 | 8 |
| P55039 | Oncology | 3125.0 | 6250.0 | 800000 | 800000 | 2.1 | 7 | 12 |
| Q8IX05 | Oncology | 195.3 | 390.6 | 25000 | 800000 | 1.8 | 10 | 9 |
| Q12846 | Oncology | 195.3 | 390.6 | 100000 | 800000 | 2.4 | 9 | 10 |
| Q96RT1 | Oncology | 97.7 | 195.3 | 12500 | 25000 | 1.8 | 7 | 13 |
| O15357 | Oncology | 97.7 | 195.3 | 50000 | 100000 | 2.4 | 6 | 19 |
| P23515 | Oncology | 6.1 | 12.2 | 12500 | 25000 | 3.0 | 12 | 12 |
| P28907 | Oncology | 6.1 | 12.2 | 6250 | 25000 | 2.7 | 7 | 14 |
| O60911 | Oncology | 24.4 | 48.8 | 6250 | 12500 | 2.1 | 4 | 6 |
| Q7Z5A7 | Oncology | 48.8 | 97.7 | 6250 | 25000 | 1.8 | 9 | 14 |
| P16870 | Oncology | 97.7 | 97.7 | 50000 | 800000 | 2.7 | 5 | 4 |
| O60760 | Oncology | 781.3 | 1562.5 | 400000 | 800000 | 2.4 | 10 | 12 |
| Q96EK5 | Oncology | 390.6 | 781.3 | 400000 | 800000 | 2.7 | 11 | 19 |
| Q8N9I9 | Oncology | 97.7 | 195.3 | 400000 | 800000 | 3.3 | 10 | 9 |
| O60825 | Oncology | 97.7 | 195.3 | 400000 | 800000 | 3.3 | 8 | 13 |
| Q9UBM4 | Oncology | 12.2 | 24.4 | 25000 | 100000 | 3.0 | 10 | 12 |
| O60763 | Oncology | 195.3 | 390.6 | 100000 | 400000 | 2.4 | 11 | 23 |

| | | | | | | | | |
|---|---|---|---|---|---|---|---|---|
| P07949 | Oncology | 24.4 | 48.8 | 25000 | 400000 | 2.7 | 8 | 10 |
| Q8N386 | Oncology | 6.1 | 24.4 | 12500 | 50000 | 2.7 | 9 | 11 |
| Q8NEZ2 | Oncology | | | | | | 8 | 10 |
| P15514 | Oncology | 1.5 | 3.1 | 6250 | 25000 | 3.3 | 8 | 10 |
| P18627 | Oncology | | | | | | 10 | 8 |
| Q86SF2 | Oncology | 781.3 | 781.3 | 50000 | 800000 | 1.8 | 9 | 9 |
| O00622 | Oncology | 48.8 | 97.7 | 25000 | 200000 | 2.4 | 9 | 9 |
| O75144 | Oncology | 48.8 | 97.7 | 12500 | 800000 | 2.1 | 9 | 13 |
| Q13576 | Oncology | | | | | | 10 | 17 |
| O00748 | Oncology | | | | | | 10 | 13 |
| P58499 | Oncology | 97.7 | 195.3 | 12500 | 800000 | 1.8 | 6 | 10 |
| P26010 | Oncology | 12.2 | 48.8 | 50000 | 200000 | 3.0 | 4 | 6 |
| Q9UKR3 | Oncology | 24.4 | 48.8 | 6250 | 50000 | 2.1 | 8 | 11 |
| P49441 | Oncology | 1562.5 | 3125.0 | 400000 | 800000 | 2.1 | 11 | 29 |
| O43570 | Oncology | 24.4 | 48.8 | 12500 | 100000 | 2.4 | 9 | 11 |
| P37108 | Oncology | 390.6 | 781.3 | 50000 | 800000 | 1.8 | 9 | 11 |
| P38936 | Oncology | 390.6 | 781.3 | 400000 | 800000 | 2.7 | 9 | 12 |
| Q13561 | Oncology | | | | | | 11 | 9 |
| O14828 | Oncology | 195.3 | 195.3 | 50000 | 400000 | 2.4 | 10 | 9 |
| P07948 | Oncology | 6.1 | 24.4 | 3125 | 6250 | 2.1 | 10 | 11 |
| Q9NZT2 | Oncology | 48.8 | 97.7 | 200000 | 800000 | 3.3 | 10 | 10 |
| P01275 | Oncology | | | | | | 15 | 31 |
| P50583 | Oncology | 97.7 | 195.3 | 12500 | 800000 | 1.8 | 8 | 13 |
| Q9Y653 | Oncology | 195.3 | 390.6 | 100000 | 400000 | 2.4 | 7 | 14 |
| Q8N129 | Oncology | 97.7 | 195.3 | 12500 | 800000 | 1.8 | 10 | 7 |
| Q49AH0 | Oncology | 12.2 | 12.2 | 12500 | 50000 | 3.0 | 9 | 10 |
| P29317 | Oncology | 12.2 | 24.4 | 25000 | 200000 | 3.0 | 7 | 9 |
| Q9Y5K8 | Oncology | | | | | | 2 | 8 |
| O00451 | Oncology | | | | | | 9 | 9 |
| P29459_P29 | Oncology | 24.4 | 48.8 | 12500 | 100000 | 2.4 | 8 | 12 |
| Q99795 | Oncology | 6.1 | 12.2 | 6250 | 12500 | 2.7 | 9 | 10 |
| Q99536 | Oncology | | | | | | 9 | 12 |
| Q9GZV9 | Oncology | 97.7 | 195.3 | 50000 | 200000 | 2.4 | 5 | 6 |

| | | | | | | | | |
|---|---|---|---|---|---|---|---|---|
| Q9H156 | Oncology | 24.4 | 48.8 | 200000 | 800000 | 3.6 | 4 | 6 |
| P98073 | Oncology | 48.8 | 195.3 | 25000 | 100000 | 2.1 | 9 | 9 |
| Q9P0G3 | Oncology | | | | | | 7 | 5 |
| O43715 | Oncology | 48.8 | 97.7 | 25000 | 100000 | 2.4 | 4 | 3 |
| Q9ULX7 | Oncology | 6.1 | 12.2 | 50000 | 400000 | 3.6 | 4 | 4 |
| Q86SR1 | Oncology | 781.3 | 1562.5 | 400000 | 800000 | 2.4 | 11 | 15 |
| Q9C005 | Oncology | 48.8 | 97.7 | 6250 | 25000 | 1.8 | 5 | 4 |
| Q13421 | Oncology | | | | | | 9 | 9 |
| Q15116 | Oncology | 3.1 | 6.1 | 12500 | 25000 | 3.3 | 4 | 10 |
| Q9UJM8 | Oncology | 97.7 | 195.3 | 400000 | 800000 | 3.3 | 4 | 7 |
| P05187 | Oncology | 3.1 | 6.1 | 12500 | 50000 | 3.3 | 8 | 11 |
| P25685 | Oncology | 3125.0 | 6250.0 | 800000 | 1600000 | 2.1 | 8 | 15 |
| Q8WXI7 | Oncology | | | | | | 14 | 22 |
| P10145 | Oncology | 0.2 | 0.4 | 1563 | 12500 | 3.6 | 5 | 4 |
| O43827 | Oncology | 48.8 | 97.7 | 25000 | 800000 | 2.4 | 3 | 5 |
| P39900 | Oncology | 3.1 | 6.1 | 12500 | 100000 | 3.3 | 5 | 5 |
| P09105 | Oncology | | | | | | 4 | 9 |
| P13521 | Oncology | 97.7 | 195.3 | 200000 | 400000 | 3.0 | 4 | 5 |
| P50120 | Oncology | 48.8 | 195.3 | 12500 | 25000 | 1.8 | 8 | 8 |
| P09960 | Oncology | 12500.0 | 25000.0 | 1600000 | 3200000 | 1.8 | 4 | 19 |
| Q9HAV5 | Oncology | 0.4 | 0.8 | 3125 | 12500 | 3.6 | 4 | 5 |
| P05089 | Oncology | 390.6 | 781.3 | 50000 | 200000 | 1.8 | 4 | 8 |
| Q9H4F8 | Oncology | 781.3 | 1562.5 | 200000 | 800000 | 2.1 | 4 | 7 |
| Q02742 | Oncology | 781.3 | 781.3 | 400000 | 800000 | 2.7 | 10 | 9 |
| O14558 | Oncology | 48.8 | 97.7 | 25000 | 400000 | 2.4 | 3 | 6 |
| Q14203 | Oncology | 195.3 | 781.3 | 400000 | 800000 | 2.7 | 4 | 9 |
| Q9Y336 | Oncology | 12.2 | 24.4 | 6250 | 25000 | 2.4 | 5 | 3 |
| P01303 | Oncology | 1562.5 | 3125.0 | 800000 | 800000 | 2.4 | 6 | 12 |
| Q9H6B4 | Oncology | 24.4 | 48.8 | 12500 | 400000 | 2.4 | 3 | 8 |
| P47929 | Oncology | 25000.0 | 25000.0 | 800000 | 1600000 | 1.5 | 6 | 13 |
| Q6UWN8 | Oncology | 12.2 | 24.4 | 12500 | 100000 | 2.7 | 3 | 5 |
| P40121 | Oncology | 781.3 | 781.3 | 400000 | 800000 | 2.7 | 3 | 8 |
| Q16595 | Oncology | 3125.0 | 6250.0 | 800000 | 800000 | 2.1 | 4 | 5 |

| | | | | | | | | |
|---|---|---|---|---|---|---|---|---|
| Q8IXJ6 | Oncology | 24.4 | 48.8 | 50000 | 100000 | 3.0 | 10 | 12 |
| P80511 | Oncology | 781.3 | 1562.5 | 400000 | 800000 | 2.4 | 9 | 17 |
| Q86SJ2 | Oncology | 48.8 | 195.3 | 12500 | 50000 | 1.8 | 9 | 7 |
| P98082 | Oncology | 195.3 | 390.6 | 50000 | 200000 | 2.1 | 4 | 12 |
| Q9BXY4 | Oncology | 24.4 | 48.8 | 50000 | 200000 | 3.0 | 3 | 6 |
| P06127 | Oncology | 0.8 | 1.5 | 3125 | 12500 | 3.3 | 5 | 6 |
| P80303 | Oncology | 97.7 | 195.3 | 12500 | 400000 | 1.8 | 4 | 5 |
| Q9NS68 | Oncology | 3.1 | 6.1 | 3125 | 12500 | 2.7 | 4 | 5 |
| Q9H6S3 | Oncology | 97.7 | 195.3 | 100000 | 400000 | 2.7 | 4 | 4 |
| O15263 | Oncology | | | | | | 12 | 12 |
| Q9Y5K2 | Oncology | 0.2 | 0.4 | 6250 | 25000 | 4.2 | 4 | 5 |
| Q9P1Z2 | Oncology | 195.3 | 390.6 | 100000 | 400000 | 2.4 | 5 | 7 |
| Q16653 | Oncology | 1.5 | 3.1 | 1563 | 3125 | 2.7 | 3 | 7 |
| P08397 | Oncology | 390.6 | 781.3 | 200000 | 800000 | 2.4 | 5 | 8 |
| Q7Z5L0 | Oncology | 1562.5 | 3125.0 | 200000 | 800000 | 1.8 | 13 | 11 |
| Q96JA1 | Oncology | 48.8 | 97.7 | 100000 | 800000 | 3.0 | 4 | 5 |
| Q16790 | Oncology | 6.1 | 6.1 | 6250 | 50000 | 3.0 | 4 | 6 |
| P09758 | Oncology | 1.5 | 3.1 | 6250 | 25000 | 3.3 | 5 | 5 |
| O60243 | Oncology | 390.6 | 781.3 | 200000 | 400000 | 2.4 | 8 | 8 |
| Q9NPH0 | Oncology | 12.2 | 24.4 | 50000 | 400000 | 3.3 | 5 | 5 |
| Q96I15 | Oncology | 97.7 | 195.3 | 100000 | 800000 | 2.7 | 3 | 5 |
| P16562 | Oncology | 6.1 | 12.2 | 12500 | 200000 | 3.0 | 4 | 6 |
| P27695 | Oncology | 6.1 | 12.2 | 6250 | 200000 | 2.7 | 3 | 5 |
| Q02246 | Oncology | 24.4 | 48.8 | 12500 | 50000 | 2.4 | 4 | 5 |
| Q9BZR6 | Oncology | 48.8 | 97.7 | 200000 | 800000 | 3.3 | 4 | 7 |
| P62166 | Oncology | 12.2 | 24.4 | 25000 | 400000 | 3.0 | 4 | 5 |
| Q10471 | Oncology | 195.3 | 390.6 | 50000 | 400000 | 2.1 | 3 | 6 |
| Q8WWY7 | Oncology | 48.8 | 48.8 | 6250 | 12500 | 2.1 | 4 | 16 |
| Q6PCB0 | Oncology | 48.8 | 195.3 | 400000 | 800000 | 3.3 | 4 | 4 |
| P51858 | Oncology | 24.4 | 24.4 | 6250 | 25000 | 2.4 | 5 | 10 |
| Q16775 | Oncology | 781.3 | 781.3 | 200000 | 800000 | 2.4 | 6 | 14 |
| Q8TDQ1 | Oncology | 12.2 | 48.8 | 12500 | 25000 | 2.4 | 6 | 6 |
| P02760 | Oncology | 1562.5 | 12500.0 | 12800000 | 12800000 | 3.0 | 5 | 6 |

| | | | | | | | | |
|---|---|---|---|---|---|---|---|---|
| Q9H3G5 | Oncology | 97.7 | 390.6 | 200000 | 800000 | 2.7 | 4 | 7 |
| Q496F6 | Oncology | 390.6 | 781.3 | 400000 | 800000 | 2.7 | 3 | 4 |
| P35052 | Oncology | 195.3 | 390.6 | 100000 | 400000 | 2.4 | 3 | 3 |
| P56159 | Oncology | 24.4 | 48.8 | 50000 | 200000 | 3.0 | 3 | 4 |
| P35475 | Oncology | 12.2 | 24.4 | 3125 | 12500 | 2.1 | 4 | 9 |
| P32926 | Oncology | 3.1 | 6.1 | 6250 | 50000 | 3.0 | 6 | 5 |
| Q96D42 | Oncology | 3.1 | 6.1 | 12500 | 50000 | 3.3 | 3 | 4 |
| P20472 | Oncology | 12.2 | 12.2 | 6250 | 25000 | 2.7 | 9 | 9 |
| O15123 | Oncology | 195.3 | 390.6 | 200000 | 800000 | 2.7 | 5 | 6 |
| P29017 | Oncology | 97.7 | 195.3 | 25000 | 400000 | 2.1 | 8 | 12 |
| Q14508 | Oncology | 12.2 | 24.4 | 25000 | 400000 | 3.0 | 7 | 6 |
| O43895 | Oncology | 12.2 | 48.8 | 50000 | 200000 | 3.0 | 6 | 6 |
| Q9UBX1 | Oncology | 97.7 | 195.3 | 50000 | 400000 | 2.4 | 7 | 8 |
| P07237 | Oncology | 24.4 | 24.4 | 12500 | 50000 | 2.7 | 8 | 13 |
| Q6FI81 | Oncology | 195.3 | 390.6 | 200000 | 400000 | 2.7 | 6 | 8 |
| P41439 | Oncology | 0.2 | 0.4 | 12500 | 50000 | 4.5 | 6 | 10 |
| Q5JTD0 | Oncology | 195.3 | 390.6 | 400000 | 800000 | 3.0 | 6 | 6 |
| O14974 | Oncology | 97.7 | 195.3 | 25000 | 100000 | 2.1 | 9 | 22 |
| P37173 | Oncology | 1.5 | 3.1 | 12500 | 25000 | 3.6 | 7 | 7 |
| Q15303 | Oncology | 3.1 | 6.1 | 12500 | 50000 | 3.3 | 7 | 4 |
| Q92832 | Oncology | 48.8 | 97.7 | 200000 | 800000 | 3.3 | 7 | 7 |
| Q96NY8 | Oncology | 1.5 | 3.1 | 3125 | 12500 | 3.0 | 7 | 6 |
| Q96J42 | Oncology | 48.8 | 97.7 | 25000 | 400000 | 2.4 | 8 | 10 |
| Q9H8J5 | Oncology | 48.8 | 97.7 | 50000 | 400000 | 2.7 | 6 | 9 |
| P21741 | Oncology | | | | | | 8 | 14 |
| Q9BYH1 | Oncology | 195.3 | 390.6 | 100000 | 400000 | 2.4 | 7 | 7 |
| Q16543 | Oncology | 390.6 | 781.3 | 100000 | 400000 | 2.1 | 8 | 23 |
| Q9NZ53 | Oncology | 48.8 | 97.7 | 25000 | 100000 | 2.4 | 8 | 13 |
| P20851 | Oncology | 6.1 | 12.2 | 12500 | 100000 | 3.0 | 8 | 6 |
| P41271 | Oncology | 24.4 | 48.8 | 12500 | 25000 | 2.4 | 7 | 8 |
| P35916 | Oncology | 781.3 | 1562.5 | 400000 | 800000 | 2.4 | 7 | 5 |
| P20138 | Oncology | 12.2 | 48.8 | 12500 | 50000 | 2.4 | 7 | 9 |
| Q96SM3 | Oncology | 195.3 | 390.6 | 200000 | 400000 | 2.7 | 8 | 10 |

| | | | | | | | | |
|---|---|---|---|---|---|---|---|---|
| P35968 | Oncology | 3.1 | 6.1 | 25000 | 50000 | 3.6 | 6 | 10 |
| Q02763 | Oncology | 48.8 | 97.7 | 100000 | 400000 | 3.0 | 7 | 9 |
| P21589 | Oncology | 6.1 | 12.2 | 25000 | 100000 | 3.3 | 7 | 8 |
| O95721 | Oncology | 48.8 | 97.7 | 100000 | 400000 | 3.0 | 12 | 13 |
| P09486 | Oncology | 195.3 | 390.6 | 200000 | 800000 | 2.7 | 8 | 10 |
| Q9UP79 | Oncology | 48.8 | 97.7 | 50000 | 200000 | 2.7 | 7 | 8 |
| P32004 | Oncology | 97.7 | 195.3 | 100000 | 400000 | 2.7 | 6 | 8 |
| O43464 | Oncology | 12.2 | 24.4 | 12500 | 25000 | 2.7 | 10 | 10 |
| Q7Z4W1 | Oncology | 195.3 | 390.6 | 100000 | 800000 | 2.4 | 5 | 8 |
| Q9HAT2 | Oncology | 195.3 | 390.6 | 50000 | 400000 | 2.1 | 6 | 5 |
| Q8NCC3 | Oncology | 24.4 | 48.8 | 50000 | 400000 | 3.0 | 7 | 6 |
| P21802 | Oncology | 12.2 | 24.4 | 25000 | 400000 | 3.0 | 6 | 6 |
| Q14512 | Oncology | | | | | | 7 | 8 |
| Q9NP84 | Oncology | 97.7 | 195.3 | 200000 | 800000 | 3.0 | 9 | 11 |
| O00244 | Oncology | 48.8 | 97.7 | 12500 | 800000 | 2.1 | 9 | 15 |
| Q96PD2 | Oncology | 6.1 | 12.2 | 12500 | 50000 | 3.0 | 8 | 7 |
| P78552 | Oncology | 24.4 | 48.8 | 12500 | 25000 | 2.4 | 6 | 9 |
| P01298 | Oncology | 97.7 | 97.7 | 12500 | 50000 | 2.1 | 7 | 12 |
| P13688 | Oncology | 6.1 | 12.2 | 25000 | 400000 | 3.3 | 8 | 8 |
| P26447 | Oncology | | | | | | 8 | 12 |
| O75629 | Oncology | 97.7 | 195.3 | 25000 | 400000 | 2.1 | 6 | 12 |
| P09958 | Oncology | 48.8 | 97.7 | 50000 | 100000 | 2.7 | 7 | 7 |
| P48307 | Oncology | 0.8 | 1.5 | 6250 | 12500 | 3.6 | 7 | 8 |
| P18084 | Oncology | 97.7 | 195.3 | 100000 | 400000 | 2.7 | 8 | 14 |
| P15328 | Oncology | 0.8 | 1.5 | 3125 | 12500 | 3.3 | 8 | 9 |
| O60259 | Oncology | 3.1 | 6.1 | 6250 | 25000 | 3.0 | 7 | 11 |
| Q9UJ68 | Oncology | 0.8 | 1.5 | 391 | 1563 | 2.4 | 7 | 9 |
| P26842 | Oncology | 24.4 | 48.8 | 25000 | 100000 | 2.7 | 7 | 8 |
| P06870 | Oncology | 1.5 | 3.1 | 6250 | 25000 | 3.3 | 7 | 8 |
| P12931 | Oncology | 24.4 | 48.8 | 12500 | 25000 | 2.4 | 7 | 14 |
| O43240 | Oncology | 24.4 | 48.8 | 25000 | 50000 | 2.7 | 8 | 25 |
| O95274 | Oncology | 6.1 | 12.2 | 6250 | 25000 | 2.7 | 6 | 15 |
| O00548 | Oncology | 6.1 | 12.2 | 50000 | 100000 | 3.6 | 7 | 9 |

| | | | | | | | | |
|---|---|---|---|---|---|---|---|---|
| P49767 | Oncology | 97.7 | 97.7 | 12500 | 400000 | 2.1 | 11 | 13 |
| P04626 | Oncology | 1.5 | 3.1 | 12500 | 50000 | 3.6 | 7 | 8 |
| Q16674 | Oncology | 31.7 | 63.5 | 65000 | 130000 | 3.0 | 6 | 6 |
| Q9UBX7 | Oncology | 48.8 | 97.7 | 25000 | 100000 | 2.4 | 7 | 40 |
| Q92876 | Oncology | 1.5 | 12.2 | 3125 | 12500 | 2.4 | 7 | 5 |
| Q6NT46 | Oncology II | 97.7 | 195.3 | 12500 | 400000 | 1.8 | 10 | 4 |
| Q7Z460 | Oncology II | | | | | | | |
| Q7Z4W2 | Oncology II | 6.1 | 12.2 | 1563 | 25000 | 2.1 | 9 | 23 |
| Q9UPY8 | Oncology II | | | | | | 2 | 3 |
| O00337 | Oncology II | | | | | | 5 | 11 |
| P23771 | Oncology II | 100000.0 | 200000.0 | 6400000 | 12800000 | 1.5 | | |
| Q5VSG8 | Oncology II | | | | | | 16 | 28 |
| P43630 | Oncology II | 390.6 | 781.3 | 100000 | 400000 | 2.1 | 8 | 19 |
| A7E2Y1 | Oncology II | 390.6 | 781.3 | 50000 | 400000 | 1.8 | 15 | 4 |
| P30304 | Oncology II | 390.6 | 781.3 | 50000 | 100000 | 1.8 | 6 | |
| Q9P2D8 | Oncology II | 781.3 | 781.3 | 100000 | 400000 | 2.1 | 14 | 20 |
| Q14123 | Oncology II | | | | | | 9 | 21 |
| P12004 | Oncology II | 6250.0 | 6250.0 | 400000 | 3200000 | 1.8 | 3 | |
| O60941 | Oncology II | 97.7 | 97.7 | 12500 | 50000 | 2.1 | 12 | 13 |
| O43504 | Oncology II | | | | | | 9 | 13 |
| P19075 | Oncology II | 3.1 | 12.2 | 6250 | 25000 | 2.7 | 4 | 25 |
| O60502 | Oncology II | 1562.5 | 3125.0 | 200000 | 800000 | 1.8 | 8 | 15 |
| Q13443 | Oncology II | 25000.0 | 25000.0 | 1600000 | 3200000 | 1.8 | 9 | 19 |
| Q93033 | Oncology II | 390.6 | 781.3 | 200000 | 800000 | 2.4 | 6 | 15 |
| Q8NHZ8 | Oncology II | 1562.5 | 1562.5 | 50000 | 800000 | 1.5 | 6 | 40 |
| Q92499 | Oncology II | | | | | | 9 | 15 |
| O75781 | Oncology II | 195.3 | 195.3 | 12500 | 50000 | 1.8 | 6 | 6 |
| Q6UWK7 | Oncology II | | | | | | 10 | 18 |
| Q9UMS0 | Oncology II | 48.8 | 97.7 | 100000 | 400000 | 3.0 | | |
| Q96KB5 | Oncology II | 97.7 | 195.3 | 100000 | 400000 | 2.7 | 8 | 8 |
| Q8ND71 | Oncology II | 195.3 | 390.6 | 25000 | 200000 | 1.8 | 16 | 21 |
| O75330 | Oncology II | 3125.0 | 6250.0 | 800000 | 800000 | 2.1 | 9 | 13 |
| Q14677 | Oncology II | 6250.0 | 12500.0 | 800000 | 800000 | 1.8 | 9 | 21 |

| | | | | | | | | |
|---|---|---|---|---|---|---|---|---|
| Q9GZT3 | Oncology II | | | | | | 12 | 22 |
| Q00994 | Oncology II | 195.3 | 390.6 | 25000 | 100000 | 1.8 | | |
| Q86SQ0 | Oncology II | | | | | | 13 | 21 |
| Q9NPI5 | Oncology II | | | | | | | |
| Q9H9E1 | Oncology II | | | | | | | |
| O43889 | Oncology II | 195.3 | 390.6 | 25000 | 800000 | 1.8 | 12 | 7 |
| P48165 | Oncology II | 3125.0 | 3125.0 | 200000 | 800000 | 1.8 | | |
| Q9UQE7 | Oncology II | | | | | | 9 | |
| O15264 | Oncology II | 6250.0 | 6250.0 | 800000 | 800000 | 2.1 | 0.2 | |
| P06493 | Oncology II | | | | | | 15 | 8 |
| Q15652 | Oncology II | 1562.5 | 3125.0 | 200000 | 800000 | 1.8 | 4 | 14 |
| P00167 | Oncology II | 195.3 | 390.6 | 50000 | 400000 | 2.1 | 11 | 18 |
| Q8N130 | Oncology II | 1562.5 | 3125.0 | 200000 | 800000 | 1.8 | 11 | |
| Q6PUV4 | Oncology II | 781.3 | 1562.5 | 100000 | 400000 | 1.8 | | |
| Q9H741 | Oncology II | | | | | | 13 | 19 |
| P78395 | Oncology II | | | | | | 5 | |
| Q3MIW9 | Oncology II | 390.6 | 390.6 | 25000 | 100000 | 1.8 | 10 | 18 |
| Q9UJZ1 | Oncology II | | | | | | 11 | 8 |
| Q12836 | Oncology II | 24.4 | 97.7 | 12500 | 50000 | 2.1 | 1 | |
| A6NGN9 | Oncology II | 781.3 | 1562.5 | 200000 | 800000 | 2.1 | 8 | 14 |
| A8MVZ5 | Oncology II | 3125.0 | 3125.0 | 200000 | 800000 | 1.8 | | |
| Q99259 | Oncology II | 6250.0 | 12500.0 | 800000 | 800000 | 1.8 | | |
| Q8N4E4 | Oncology II | 195.3 | 390.6 | 50000 | 200000 | 2.1 | 11 | 8 |
| Q8WZ55 | Oncology II | 195.3 | 390.6 | 25000 | 200000 | 1.8 | | |
| P18848 | Oncology II | 48.8 | 48.8 | 6250 | 400000 | 2.1 | 12 | |
| O00534 | Oncology II | 781.3 | 1562.5 | 100000 | 400000 | 1.8 | 10 | 17 |
| P31689 | Oncology II | | | | | | 0.3 | |
| P31371 | Oncology II | | | | | | 1 | |
| Q8IYV9 | Oncology II | 97.7 | 195.3 | 200000 | 800000 | 3.0 | | |
| Q13017 | Oncology II | | | | | | 9 | 14 |
| Q12899 | Oncology II | | | | | | | |
| Q15014 | Oncology II | 781.3 | 781.3 | 50000 | 400000 | 1.8 | 11 | 11 |
| Q9H867 | Oncology II | | | | | | 8 | 23 |

| | | | | | | | | |
|---|---|---|---|---|---|---|---|---|
| Q14674 | Oncology II | 390.6 | 781.3 | 50000 | 400000 | 1.8 | 11 | 18 |
| P17980 | Oncology II | | | | | | | |
| Q9NS37 | Oncology II | 6250.0 | 6250.0 | 800000 | 800000 | 2.1 | | |
| O43739 | Oncology II | 781.3 | 781.3 | 50000 | 400000 | 1.8 | | |
| Q9H293 | Oncology II | 195.3 | 390.6 | 100000 | 400000 | 2.4 | 4 | |
| Q587J8 | Oncology II | | | | | | 12 | 21 |
| Q4VC05 | Oncology II | 781.3 | 781.3 | 50000 | 800000 | 1.8 | | |
| Q9NZQ9 | Oncology II | 1562.5 | 3125.0 | 200000 | 800000 | 1.8 | 7 | |
| O75794 | Oncology II | 1562.5 | 1562.5 | 100000 | 800000 | 1.8 | 16 | |
| Q8NDC4 | Oncology II | 390.6 | 390.6 | 100000 | 400000 | 2.4 | 10 | 20 |
| Q8TE77 | Oncology II | 6250.0 | 12500.0 | 800000 | 3200000 | 1.8 | 9 | 3 |
| Q13127 | Oncology II | | | | | | 8 | 28 |
| O94986 | Oncology II | 97.7 | 195.3 | 25000 | 50000 | 2.1 | 16 | 36 |
| Q9GZP4 | Oncology II | 1562.5 | 1562.5 | 100000 | 400000 | 1.8 | 7 | 20 |
| Q8TDX7 | Oncology II | 6250.0 | 6250.0 | 800000 | 800000 | 2.1 | 9 | 15 |
| A8MTB9 | Oncology II | 195.3 | 390.6 | 50000 | 200000 | 2.1 | 10 | 12 |
| Q9Y6I3 | Oncology II | | | | | | 11 | 20 |
| Q5VT06 | Oncology II | | | | | | 18 | |
| P01100 | Oncology II | 48.8 | 97.7 | 50000 | 200000 | 2.7 | | |
| P21781 | Oncology II | 195.3 | 195.3 | 12500 | 800000 | 1.8 | 14 | 13 |
| P29353 | Oncology II | | | | | | | |
| P08700 | Oncology II | 195.3 | 781.3 | 200000 | 800000 | 2.4 | | |
| P34910 | Oncology II | 781.3 | 1562.5 | 100000 | 400000 | 1.8 | 8 | 16 |
| Q6P996 | Oncology II | 3125.0 | 3125.0 | 200000 | 400000 | 1.8 | | |
| Q9UN42 | Oncology II | | | | | | 14 | 12 |
| Q6PH85 | Oncology II | 1562.5 | 1562.5 | 100000 | 800000 | 1.8 | 2 | |
| Q9UI15 | Oncology II | | | | | | 15 | |
| O75409 | Oncology II | 97.7 | 195.3 | 25000 | 50000 | 2.1 | | |
| Q969P6 | Oncology II | 3125.0 | 6250.0 | 400000 | 800000 | 1.8 | 9 | 10 |
| Q9Y3C4 | Oncology II | 3125.0 | 6250.0 | 400000 | 3200000 | 1.8 | 11 | |
| Q12849 | Oncology II | 390.6 | 781.3 | 50000 | 100000 | 1.8 | 16 | 21 |
| A8MYV0 | Oncology II | 48.8 | 97.7 | 25000 | 100000 | 2.4 | | |
| P49756 | Oncology II | 3125.0 | 3125.0 | 200000 | 800000 | 1.8 | 4 | |

| | | | | | | | | |
|---|---|---|---|---|---|---|---|---|
| Q7Z6A9 | Oncology II | 195.3 | 195.3 | 12500 | 50000 | 1.8 | 18 | 33 |
| O43422 | Oncology II | 48.8 | 97.7 | 25000 | 100000 | 2.4 | 14 | 26 |
| Q86TS9 | Oncology II | | | | | | 8 | |
| P07992 | Oncology II | 781.3 | 3125.0 | 200000 | 800000 | 1.8 | | |
| Q96SD1 | Oncology II | | | | | | 12 | |
| Q13087 | Oncology II | 781.3 | 1562.5 | 200000 | 800000 | 2.1 | 13 | 18 |
| Q9H0R8 | Oncology II | | | | | | | |
| Q9ULR5 | Oncology II | | | | | | 19 | 18 |
| Q13972 | Oncology II | 97.7 | 195.3 | 25000 | 50000 | 2.1 | | |
| Q9NQP4 | Oncology II | | | | | | 14 | 22 |
| Q06609 | Oncology II | 24.4 | 97.7 | 50000 | 200000 | 2.7 | 7 | |
| Q9UBU8 | Oncology II | | | | | | | |
| Q14554 | Oncology II | | | | | | 14 | 31 |
| P27701 | Oncology II | 195.3 | 390.6 | 25000 | 50000 | 1.8 | 13 | 15 |
| Q03111 | Oncology II | 781.3 | 781.3 | 100000 | 800000 | 2.1 | 21 | |
| O95793 | Oncology II | | | | | | 9 | 17 |
| Q14055 | Oncology II | 3125.0 | 6250.0 | 400000 | 800000 | 1.8 | 12 | 25 |
| Q9UNP9 | Oncology II | 1562.5 | 3125.0 | 400000 | 800000 | 2.1 | 14 | 6 |
| Q00653 | Oncology II | 195.3 | 390.6 | 25000 | 200000 | 1.8 | | |
| Q9UHJ6 | Oncology II | 1562.5 | 3125.0 | 200000 | 800000 | 1.8 | 12 | 11 |
| P42681 | Oncology II | 6250.0 | 12500.0 | 800000 | 800000 | 1.8 | 8 | 25 |
| Q68DV7 | Oncology II | 390.6 | 781.3 | 100000 | 400000 | 2.1 | 12 | 18 |
| O60603 | Oncology II | 195.3 | 390.6 | 100000 | 400000 | 2.4 | 16 | 30 |
| P01112 | Oncology II | 97.7 | 195.3 | 25000 | 50000 | 2.1 | 6 | 9 |
| Q03518 | Oncology II | | | | | | 10 | 3 |
| O15078 | Oncology II | 390.6 | 781.3 | 50000 | 100000 | 1.8 | 12 | 10 |
| Q99487 | Oncology II | 3125.0 | 6250.0 | 400000 | 800000 | 1.8 | 14 | 17 |
| Q9UHL9 | Oncology II | 781.3 | 1562.5 | 100000 | 400000 | 1.8 | 8 | 13 |
| O43903 | Oncology II | 1562.5 | 1562.5 | 200000 | 800000 | 2.1 | 10 | 15 |
| P47928 | Oncology II | 390.6 | 781.3 | 100000 | 200000 | 2.1 | 8 | 14 |
| O14879 | Oncology II | 781.3 | 781.3 | 100000 | 200000 | 2.1 | 8 | 14 |
| Q9H2G2 | Oncology II | | | | | | 14 | 17 |
| P49137 | Oncology II | 1562.5 | 1562.5 | 200000 | 800000 | 2.1 | 13 | 16 |

| | | | | | | | | |
|---|---|---|---|---|---|---|---|---|
| B0FP48 | Oncology II | 195.3 | 390.6 | 25000 | 50000 | 1.8 | 9 | 16 |
| P43166 | Oncology II | 1562.5 | 3125.0 | 400000 | 800000 | 2.1 | | |
| Q13445 | Oncology II | 12500.0 | 25000.0 | 800000 | 6400000 | 1.5 | 20 | |
| P11310 | Oncology II | | | | | | 14 | 27 |
| P49662 | Oncology II | 97.7 | 195.3 | 200000 | 400000 | 3.0 | 10 | 24 |
| Q6N021 | Oncology II | 195.3 | 390.6 | 25000 | 50000 | 1.8 | 12 | 12 |
| Q8TCU4 | Oncology II | 390.6 | 390.6 | 25000 | 50000 | 1.8 | 14 | 34 |
| P59282 | Oncology II | | | | | | 12 | 9 |
| Q8WWU5 | Oncology II | 1562.5 | 1562.5 | 200000 | 800000 | 2.1 | 8 | |
| Q9UK41 | Oncology II | 3125.0 | 6250.0 | 400000 | 800000 | 1.8 | 12 | 18 |
| A6NLU5 | Oncology II | | | | | | 5 | 21 |
| O75665 | Oncology II | | | | | | 7 | 11 |
| O15164 | Oncology II | 195.3 | 390.6 | 50000 | 100000 | 2.1 | 14 | 21 |
| O95777 | Oncology II | 6250.0 | 12500.0 | 800000 | 12800000 | 1.8 | 8 | 9 |
| O43247 | Oncology II | | | | | | | |
| P04183 | Oncology II | | | | | | 10 | 3 |
| Q16181 | Oncology II | | | | | | 14 | 21 |
| O15294 | Oncology II | 12500.0 | 25000.0 | 800000 | 800000 | 1.5 | 3 | 11 |
| Q9HCM3 | Oncology II | | | | | | 20 | 14 |
| Q96F10 | Oncology II | 97.7 | 97.7 | 25000 | 50000 | 2.4 | 13 | 17 |
| Q2M296 | Oncology II | | | | | | 15 | 34 |
| O60237 | Oncology II | 781.3 | 781.3 | 50000 | 200000 | 1.8 | 17 | 20 |
| P51815 | Oncology II | 1562.5 | 3125.0 | 200000 | 800000 | 1.8 | 6 | 17 |
| O95696 | Oncology II | 1562.5 | 1562.5 | 100000 | 800000 | 1.8 | 13 | |
| P07766 | Oncology II | 97.7 | 195.3 | 100000 | 800000 | 2.7 | 15 | |
| Q8TEW0 | Oncology II | | | | | | 12 | |
| P19526 | Oncology II | 3125.0 | 3125.0 | 200000 | 800000 | 1.8 | 11 | 14 |
| P10398 | Oncology II | 1562.5 | 1562.5 | 100000 | 800000 | 1.8 | 12 | 7 |
| P78358 | Oncology II | 48.8 | 97.7 | 25000 | 50000 | 2.4 | 17 | |
| Q8WUY3 | Oncology II | 195.3 | 390.6 | 50000 | 200000 | 2.1 | 14 | 17 |
| P22528 | Oncology II | | | | | | 11 | 18 |
| O15211 | Oncology II | 390.6 | 390.6 | 25000 | 50000 | 1.8 | 13 | 33 |
| P15248 | Oncology II | 48.8 | 97.7 | 12500 | 50000 | 2.1 | 11 | 12 |

| | | | | | | | | |
|---|---|---|---|---|---|---|---|---|
| O75293 | Oncology II | 3125.0 | 3125.0 | 200000 | 3200000 | 1.8 | 9 | 16 |
| Q6UY09 | Oncology II | 48.8 | 97.7 | 25000 | 100000 | 2.4 | 11 | 16 |
| O00422 | Oncology II | 195.3 | 390.6 | 100000 | 400000 | 2.4 | 14 | 28 |
| Q5VUJ9 | Oncology II | | | | | | 13 | 13 |
| Q9Y4G8 | Oncology II | | | | | | 15 | 8 |
| Q14204 | Oncology II | 195.3 | 390.6 | 100000 | 400000 | 2.4 | 6 | 11 |
| P25092 | Oncology II | 390.6 | 390.6 | 50000 | 200000 | 2.1 | 14 | |
| Q32MZ4 | Oncology II | | | | | | 11 | 21 |
| Q7Z6I6 | Oncology II | 3125.0 | 3125.0 | 200000 | 800000 | 1.8 | 16 | |
| P50454 | Oncology II | 6250.0 | 6250.0 | 400000 | 800000 | 1.8 | | |
| Q96NB3 | Oncology II | 6250.0 | 6250.0 | 200000 | 800000 | 1.5 | 7 | 6 |
| Q9UMX5 | Oncology II | 1562.5 | 3125.0 | 200000 | 800000 | 1.8 | 8 | 20 |
| Q9ULD2 | Oncology II | 12.2 | 24.4 | 6250 | 50000 | 2.4 | 20 | 15 |
| P43357 | Oncology II | 781.3 | 1562.5 | 100000 | 800000 | 1.8 | | |
| P78317 | Oncology II | | | | | | | |
| Q9Y2J4 | Oncology II | 1562.5 | 3125.0 | 400000 | 800000 | 2.1 | 9 | 10 |
| Q495A1 | Oncology II | 97.7 | 195.3 | 12500 | 200000 | 1.8 | 15 | 18 |
| O14717 | Oncology II | 97.7 | 195.3 | 100000 | 200000 | 2.7 | 14 | 26 |
| P30279 | Oncology II | 1562.5 | 3125.0 | 400000 | 800000 | 2.1 | 11 | 14 |
| P26639 | Oncology II | 3125.0 | 6250.0 | 800000 | 800000 | 2.1 | 7 | 19 |
| Q9UJ99 | Oncology II | 12500.0 | 12500.0 | 800000 | 3200000 | 1.8 | 10 | 36 |
| Q8IV48 | Oncology II | | | | | | 11 | 35 |
| P33764 | Oncology II | 97.7 | 195.3 | 25000 | 50000 | 2.1 | 9 | 25 |
| Q9NYJ8 | Oncology II | 3125.0 | 6250.0 | 400000 | 800000 | 1.8 | 14 | 22 |
| P53420 | Oncology II | | | | | | 19 | |
| Q6UXC1 | Oncology II | 48.8 | 97.7 | 25000 | 50000 | 2.4 | 6 | 18 |
| Q15796 | Oncology II | 25000.0 | 50000.0 | 3200000 | 12800000 | 1.8 | 8 | 13 |
| Q92973 | Oncology II | | | | | | 12 | 9 |
| Q6ZVL6 | Oncology II | 1562.5 | 1562.5 | 100000 | 200000 | 1.8 | | |
| Q9HC77 | Oncology II | 195.3 | 195.3 | 25000 | 50000 | 2.1 | | |
| O43768 | Oncology II | 12500.0 | 12500.0 | 800000 | 1600000 | 1.8 | 12 | 25 |
| Q8WWF5 | Oncology II | 781.3 | 781.3 | 100000 | 200000 | 2.1 | 7 | 23 |
| P51948 | Oncology II | 390.6 | 781.3 | 200000 | 800000 | 2.4 | 7 | 8 |

| | | | | | | | | |
|---|---|---|---|---|---|---|---|---|
| P49366 | Oncology II | 781.3 | 781.3 | 25000 | 200000 | 1.5 | 13 | 20 |
| Q92817 | Oncology II | 390.6 | 390.6 | 100000 | 200000 | 2.4 | 14 | 14 |
| P78540 | Oncology II | 100000.0 | 100000.0 | 6400000 | 12800000 | 1.8 | 9 | 14 |
| Q6P995 | Oncology II | 195.3 | 390.6 | 100000 | 400000 | 2.4 | 9 | 8 |
| Q9NQ84 | Oncology II | 781.3 | 1562.5 | 400000 | 800000 | 2.4 | 11 | 20 |
| P09914 | Oncology II | | | | | | | |
| O95997 | Oncology II | 390.6 | 781.3 | 100000 | 800000 | 2.1 | 14 | 6 |
| Q7Z6P3 | Oncology II | 1562.5 | 3125.0 | 200000 | 800000 | 1.8 | 12 | 19 |
| Q9BW66 | Oncology II | 6250.0 | 12500.0 | 1600000 | 6400000 | 2.1 | 8 | 19 |
| O43663 | Oncology II | 1562.5 | 3125.0 | 200000 | 800000 | 1.8 | 15 | 15 |
| P18754 | Oncology II | 390.6 | 781.3 | 50000 | 200000 | 1.8 | 8 | 12 |
| P20702 | Oncology II | 1562.5 | 1562.5 | 100000 | 800000 | 1.8 | 9 | 37 |
| Q99963 | Oncology II | 6250.0 | 6250.0 | 400000 | 800000 | 1.8 | | |
| Q96AT9 | Oncology II | | | | | | 12 | 18 |
| P49454 | Oncology II | 390.6 | 390.6 | 50000 | 200000 | 2.1 | 13 | 26 |
| Q6P5Z2 | Oncology II | 1562.5 | 1562.5 | 200000 | 800000 | 2.1 | 17 | 12 |
| O75843 | Oncology II | 12.2 | 48.8 | 25000 | 200000 | 2.7 | 8 | 15 |
| Q9UHY7 | Oncology II | | | | | | 9 | 18 |
| P21695 | Oncology II | 3125.0 | 3125.0 | 200000 | 800000 | 1.8 | 8 | 25 |
| O15213 | Oncology II | 200000.0 | 200000.0 | 6400000 | 12800000 | 1.5 | 9 | 9 |
| Q9P000 | Oncology II | 195.3 | 390.6 | 50000 | 200000 | 2.1 | 11 | 18 |
| Q14641 | Oncology II | 97.7 | 195.3 | 25000 | 100000 | 2.1 | 14 | 17 |
| O75460 | Oncology II | 195.3 | 390.6 | 100000 | 200000 | 2.4 | 9 | 13 |
| Q8IYS2 | Oncology II | 12500.0 | 12500.0 | 800000 | 800000 | 1.8 | | |
| P52848 | Oncology II | 195.3 | 390.6 | 50000 | 200000 | 2.1 | 9 | 14 |
| Q96IQ7 | Oncology II | 195.3 | 195.3 | 6250 | 50000 | 1.5 | 11 | 13 |
| Q6B8I1 | Oncology II | 781.3 | 781.3 | 50000 | 400000 | 1.8 | 10 | 29 |
| Q9GZZ8 | Oncology II | 6250.0 | 6250.0 | 400000 | 3200000 | 1.8 | 10 | 25 |
| Q9HC57 | Oncology II | | | | | | 11 | 12 |
| Q16891 | Oncology II | 781.3 | 781.3 | 50000 | 800000 | 1.8 | 13 | 11 |
| P56851 | Oncology II | 24.4 | 48.8 | 3125 | 12500 | 1.8 | 11 | 33 |
| P63146 | Oncology II | | | | | | 7 | 11 |
| Q5VX71 | Oncology II | 48.8 | 48.8 | 25000 | 100000 | 2.7 | 14 | 13 |

| | | | | | | | | |
|---|---|---|---|---|---|---|---|---|
| Q9BZW2 | Oncology II | | | | | | 9 | 12 |
| Q92485 | Oncology II | 12500.0 | 12500.0 | 800000 | 3200000 | 1.8 | 9 | 16 |
| P49354 | Oncology II | 24.4 | 48.8 | 12500 | 50000 | 2.4 | 9 | 16 |
| P05091 | Oncology II | 781.3 | 1562.5 | 100000 | 400000 | 1.8 | 5 | 27 |
| Q9UKM9 | Oncology II | 781.3 | 781.3 | 100000 | 800000 | 2.1 | 11 | 13 |
| O95433 | Oncology II | | | | | | 11 | 9 |
| Q13287 | Oncology II | 195.3 | 390.6 | 200000 | 800000 | 2.7 | 12 | 22 |
| Q06643 | Oncology II | 390.6 | 390.6 | 50000 | 200000 | 2.1 | 9 | 18 |
| Q6PKG0 | Oncology II | 781.3 | 1562.5 | 200000 | 800000 | 2.1 | 13 | 21 |
| Q9Y5E8 | Oncology II | 1562.5 | 3125.0 | 200000 | 800000 | 1.8 | 8 | 16 |
| P35249 | Oncology II | 781.3 | 1562.5 | 200000 | 800000 | 2.1 | 7 | 9 |
| Q8NEB7 | Oncology II | 195.3 | 390.6 | 25000 | 100000 | 1.8 | 12 | 12 |
| Q9BU02 | Oncology II | 195.3 | 390.6 | 25000 | 200000 | 1.8 | 7 | 25 |
| O60749 | Oncology II | | | | | | 8 | 17 |
| O75351 | Oncology II | 781.3 | 1562.5 | 100000 | 800000 | 1.8 | 7 | 32 |
| Q6UW88 | Oncology II | 195.3 | 390.6 | 25000 | 100000 | 1.8 | 2 | 8 |
| P20807 | Oncology II | 195.3 | 390.6 | 100000 | 800000 | 2.4 | 12 | 21 |
| Q8N5J2 | Oncology II | | | | | | | |
| Q96RE7 | Oncology II | 781.3 | 781.3 | 50000 | 200000 | 1.8 | 12 | 12 |
| Q86TE4 | Oncology II | 195.3 | 390.6 | 25000 | 800000 | 1.8 | 11 | 10 |
| Q8WUD1 | Oncology II | 3125.0 | 3125.0 | 200000 | 800000 | 1.8 | 12 | 21 |
| Q9NUW8 | Oncology II | 195.3 | 390.6 | 25000 | 200000 | 1.8 | 19 | 36 |
| A1KZ92 | Oncology II | 781.3 | 781.3 | 50000 | 400000 | 1.8 | 12 | 21 |
| Q49A26 | Oncology II | | | | | | 10 | 17 |
| Q6UW15 | Oncology II | | | | | | 18 | 13 |
| P01266 | Oncology II | 6.1 | 12.2 | 3125 | 12500 | 2.4 | 8 | 16 |
| Q9GZX6 | Oncology II | 12.2 | 24.4 | 6250 | 25000 | 2.4 | 13 | 15 |
| Q8N0Z9 | Oncology II | 195.3 | 390.6 | 100000 | 400000 | 2.4 | 11 | 14 |
| Q13410 | Oncology II | 3.1 | 6.1 | 3125 | 25000 | 2.7 | 13 | 14 |
| P31785 | Oncology II | 390.6 | 781.3 | 50000 | 800000 | 1.8 | 12 | 13 |
| Q8IY22 | Oncology II | 6250.0 | 12500.0 | 800000 | 3200000 | 1.8 | 8 | 16 |
| Q9BW85 | Oncology II | 781.3 | 781.3 | 50000 | 200000 | 1.8 | 12 | 13 |
| P36873 | Oncology II | 1562.5 | 3125.0 | 200000 | 800000 | 1.8 | 6 | 20 |

| | | | | | | | | |
|---|---|---|---|---|---|---|---|---|
| Q14160 | Oncology II | 195.3 | 390.6 | 50000 | 800000 | 2.1 | 6 | 20 |
| P29536 | Oncology II | | | | | | 14 | 24 |
| Q765P7 | Oncology II | | | | | | 14 | 32 |
| Q7Z692 | Oncology II | 48.8 | 97.7 | 6250 | 50000 | 1.8 | 17 | 23 |
| O95166 | Oncology II | 1562.5 | 1562.5 | 100000 | 400000 | 1.8 | 11 | 23 |
| P19525 | Oncology II | | | | | | 6 | 17 |
| Q14BN4 | Oncology II | 97.7 | 195.3 | 12500 | 50000 | 1.8 | 7 | 21 |
| A6NM11 | Oncology II | 781.3 | 1562.5 | 100000 | 400000 | 1.8 | 5 | 19 |
| Q9Y3L3 | Oncology II | 1562.5 | 3125.0 | 200000 | 800000 | 1.8 | 6 | 29 |
| O75528 | Oncology II | | | | | | 13 | 21 |
| Q9H8Y8 | Oncology II | 6250.0 | 6250.0 | 400000 | 1600000 | 1.8 | 7 | 25 |
| Q96T91 | Oncology II | 97.7 | 97.7 | 6250 | 50000 | 1.8 | 5 | 7 |
| P61812 | Oncology II | 12500.0 | 12500.0 | 400000 | 1600000 | 1.5 | 15 | 23 |
| P21912 | Oncology II | 781.3 | 1562.5 | 200000 | 800000 | 2.1 | 7 | 35 |
| Q96AQ6 | Oncology II | | | | | | 11 | 9 |
| Q8TBM8 | Oncology II | 48.8 | 97.7 | 25000 | 100000 | 2.4 | 8 | 13 |
| Q9HD43 | Oncology II | 12.2 | 48.8 | 25000 | 200000 | 2.7 | 7 | 16 |
| P84022 | Oncology II | 12500.0 | 12500.0 | 800000 | 800000 | 1.8 | 13 | 17 |
| P43632 | Oncology II | 48.8 | 97.7 | 12500 | 200000 | 2.1 | 7 | 17 |
| P43627 | Oncology II | 97.7 | 97.7 | 6250 | 50000 | 1.8 | 15 | 27 |
| Q86UX2 | Oncology II | 781.3 | 1562.5 | 100000 | 400000 | 1.8 | 17 | 37 |
| Q8NG06 | Oncology II | 390.6 | 390.6 | 25000 | 200000 | 1.8 | 7 | 11 |
| Q9UHP3 | Oncology II | 97.7 | 195.3 | 12500 | 50000 | 1.8 | 7 | 14 |
| P13051 | Oncology II | | | | | | 12 | 28 |
| Q9H2R5 | Oncology II | 97.7 | 97.7 | 12500 | 100000 | 2.1 | 13 | 20 |
| Q9BRQ6 | Oncology II | 781.3 | 781.3 | 100000 | 400000 | 2.1 | 9 | 10 |
| Q92783 | Oncology II | | | | | | 10 | 14 |
| P54652 | Oncology II | 195.3 | 781.3 | 50000 | 200000 | 1.8 | 11 | 13 |
| Q6WCQ1 | Oncology II | | | | | | 15 | 18 |
| Q86SQ7 | Oncology II | 6.1 | 24.4 | 12500 | 50000 | 2.7 | 10 | 19 |
| Q9H6E4 | Oncology II | 1562.5 | 3125.0 | 200000 | 800000 | 1.8 | 16 | 10 |
| Q9ULC4 | Oncology II | | | | | | 4 | 8 |
| P51808 | Oncology II | | | | | | 14 | 28 |

| | | | | | | | | |
|---|---|---|---|---|---|---|---|---|
| Q15599 | Oncology II | | | | | | 9 | 20 |
| Q05193 | Oncology II | 1562.5 | 3125.0 | 200000 | 800000 | 1.8 | 6 | 40 |
| P33316 | Oncology II | 1562.5 | 3125.0 | 400000 | 800000 | 2.1 | 13 | 29 |
| O60245 | Oncology II | 3125.0 | 3125.0 | 200000 | 800000 | 1.8 | 5 | 12 |
| Q16549 | Oncology II | 1562.5 | 3125.0 | 200000 | 800000 | 1.8 | 18 | 22 |
| P10415 | Oncology II | 195.3 | 195.3 | 50000 | 200000 | 2.4 | 11 | 19 |
| O43805 | Oncology II | | | | | | 6 | 18 |
| Q96K21 | Oncology II | 1562.5 | 3125.0 | 400000 | 800000 | 2.1 | 5 | 29 |
| Q6UW56 | Oncology II | 48.8 | 48.8 | 12500 | 50000 | 2.4 | 7 | 9 |
| O60232 | Oncology II | 1562.5 | 3125.0 | 200000 | 800000 | 1.8 | 6 | 13 |
| P11387 | Oncology II | | | | | | 6 | 21 |
| Q96A25 | Oncology II | 97.7 | 97.7 | 12500 | 50000 | 2.1 | 8 | 33 |
| Q9BQE9 | Oncology II | 390.6 | 781.3 | 50000 | 800000 | 1.8 | 14 | |
| Q9Y5Q6 | Oncology II | 97.7 | 97.7 | 12500 | 50000 | 2.1 | | |
| Q03169 | Oncology II | 48.8 | 48.8 | 12500 | 50000 | 2.4 | 5 | 7 |
| P53384 | Oncology II | | | | | | 5 | |
| Q7Z4H3 | Oncology II | 195.3 | 390.6 | 50000 | 800000 | 2.1 | 6 | 8 |
| P14902 | Oncology II | 195.3 | 390.6 | 100000 | 400000 | 2.4 | 7 | 14 |
| Q16819 | Oncology II | 195.3 | 195.3 | 50000 | 800000 | 2.4 | 5 | 17 |
| Q9P013 | Oncology II | 195.3 | 781.3 | 50000 | 200000 | 1.8 | 6 | 17 |
| O00203 | Oncology II | 195.3 | 781.3 | 50000 | 800000 | 1.8 | 7 | 26 |
| Q8IXQ3 | Oncology II | 195.3 | 390.6 | 25000 | 200000 | 1.8 | 5 | 19 |
| O75940 | Oncology II | | | | | | 8 | 14 |
| Q68D85 | Oncology II | 195.3 | 390.6 | 100000 | 400000 | 2.4 | 6 | 12 |
| Q9UH65 | Oncology II | 781.3 | 781.3 | 50000 | 800000 | 1.8 | 6 | 8 |
| Q14011 | Oncology II | | | | | | 7 | 37 |
| P17181 | Oncology II | 195.3 | 195.3 | 25000 | 800000 | 2.1 | 5 | 7 |
| Q676U5 | Oncology II | 781.3 | 1562.5 | 200000 | 800000 | 2.1 | 5 | 10 |
| Q96RF0 | Oncology II | 12500.0 | 12500.0 | 800000 | 1600000 | 1.8 | 6 | 11 |
| Q15172 | Oncology II | | | | | | | |
| Q86UU1 | Oncology II | | | | | | 21 | 22 |
| O43312 | Oncology II | | | | | | | |
| P20700 | Oncology II | | | | | | 17 | 16 |

| | | | | | | | | |
|---|---|---|---|---|---|---|---|---|
| Q02750 | Oncology II | 195.3 | 390.6 | 200000 | 800000 | 2.7 | 4 | 28 |
| Q99733 | Oncology II | 1562.5 | 1562.5 | 100000 | 800000 | 1.8 | 4 | 12 |
| O43653 | Oncology II | | | | | | 5 | 8 |
| Q6NUJ1 | Oncology II | 12.2 | 97.7 | 6250 | 25000 | 1.8 | 5 | 12 |
| P54577 | Oncology II | | | | | | | |
| Q2WEN9 | Oncology II | 12.2 | 24.4 | 6250 | 50000 | 2.4 | 5 | 8 |
| P42081 | Oncology II | 97.7 | 97.7 | 12500 | 200000 | 2.1 | 6 | 7 |
| Q8WXX5 | Oncology II | | | | | | 7 | 14 |
| Q8IV16 | Oncology II | | | | | | 23 | 25 |
| Q63HQ2 | Oncology II | 781.3 | 1562.5 | 200000 | 800000 | 2.1 | 7 | 6 |
| Q9UIM3 | Oncology II | 195.3 | 390.6 | 100000 | 400000 | 2.4 | 8 | 20 |
| P21128 | Oncology II | 97.7 | 195.3 | 25000 | 50000 | 2.1 | 7 | 11 |
| O75830 | Oncology II | 195.3 | 390.6 | 25000 | 200000 | 1.8 | 6 | 14 |
| Q14246 | Oncology II | 1562.5 | 1562.5 | 100000 | 800000 | 1.8 | 8 | 10 |
| Q96DE0 | Oncology II | | | | | | 5 | 31 |
| Q9Y5S2 | Oncology II | | | | | | 6 | 14 |
| O75071 | Oncology II | 25000.0 | 25000.0 | 1600000 | 3200000 | 1.8 | 7 | 6 |
| Q8TF64 | Oncology II | 195.3 | 390.6 | 25000 | 200000 | 1.8 | 7 | 36 |
| Q15262 | Oncology II | 3125.0 | 6250.0 | 800000 | 3200000 | 2.1 | 7 | 12 |
| Q14258 | Oncology II | 781.3 | 781.3 | 50000 | 200000 | 1.8 | 6 | 16 |
| P13284 | Oncology II | | | | | | 5 | 11 |
| Q674X7 | Oncology II | 781.3 | 1562.5 | 400000 | 800000 | 2.4 | 5 | 23 |
| Q92890 | Oncology II | 781.3 | 781.3 | 200000 | 400000 | 2.4 | | |
| Q6PL24 | Oncology II | 97.7 | 195.3 | 25000 | 200000 | 2.1 | 5 | 32 |
| Q7Z569 | Oncology II | 48.8 | 97.7 | 25000 | 800000 | 2.4 | 5 | 27 |
| Q8N6M0 | Oncology II | | | | | | 7 | 26 |
| P53990 | Oncology II | | | | | | | |
| O94988 | Oncology II | 97.7 | 390.6 | 25000 | 200000 | 1.8 | 6 | 25 |
| Q17RW2 | Oncology II | | | | | | 24 | 30 |
| P49223 | Oncology II | 24.4 | 24.4 | 6250 | 25000 | 2.4 | | |
| Q99447 | Oncology II | 781.3 | 1562.5 | 100000 | 400000 | 1.8 | 6 | 22 |
| Q96BQ1 | Oncology II | 48.8 | 97.7 | 12500 | 50000 | 2.1 | 4 | 18 |
| Q9H910 | Oncology II | | | | | | 8 | 39 |

| | | | | | | | | |
|---|---|---|---|---|---|---|---|---|
| P17568 | Oncology II | | | | | | 9 | 33 |
| Q9H2K0 | Oncology II | 781.3 | 3125.0 | 100000 | 800000 | 1.5 | | |
| Q9HC56 | Oncology II | 3125.0 | 6250.0 | 800000 | 800000 | 2.1 | 6 | 9 |
| Q9NPJ3 | Oncology II | 781.3 | 781.3 | 25000 | 200000 | 1.5 | 8 | 37 |
| Q6BCY4 | Oncology II | | | | | | 6 | 9 |
| P62330 | Oncology II | | | | | | | |
| Q8IWZ8 | Oncology II | 3125.0 | 6250.0 | 200000 | 800000 | 1.5 | 11 | 17 |
| P48060 | Oncology II | | | | | | 14 | 19 |
| Q96R05 | Oncology II | 781.3 | 1562.5 | 100000 | 200000 | 1.8 | 7 | 14 |
| O15182 | Oncology II | 781.3 | 781.3 | 50000 | 400000 | 1.8 | 4 | 18 |
